# Linking common and rare disease genetics through gene regulatory networks

**DOI:** 10.1101/2021.10.21.21265342

**Authors:** Olivier B. Bakker, Annique Claringbould, Harm-Jan Westra, Henry Wiersma, Floranne Boulogne, Urmo Võsa, Sophie Mulcahy Symmons, Iris H. Jonkers, Lude Franke, Patrick Deelen

## Abstract

Genetic variants identified through genome-wide association studies (GWAS) are typically non-coding and exert small regulatory effects on downstream genes, but which downstream genes are ultimately impacted and how they confer risk remains mostly unclear. Conversely, variants that cause rare Mendelian diseases are often coding and have a more direct impact on disease development. We demonstrate that common and rare genetic diseases can be linked by studying the gene regulatory networks impacted by common disease-associated variants. We implemented this in the ‘Downstreamer’ method and applied it to 44 GWAS traits and find that predicted downstream “key genes” are enriched with Mendelian disease genes, e.g. key genes for height are enriched for genes that cause skeletal abnormalities and Ehlers-Danlos syndromes. We find that 82% of these key genes are located outside of GWAS loci, suggesting that they result from complex *trans* regulation rather than being impacted by disease-associated variants in *cis*. Finally, we discuss the challenges in reconstructing gene regulatory networks and provide a roadmap to improve identification of these highly connected genes for common traits and diseases.

## Introduction

Genetic variation plays a major role in the development of both common and rare diseases, yet the genetic architectures of these disease types are usually considered quite different. Rare genetic disorders are thought to primarily be caused by a single, mostly protein-coding genetic variant that has a large effect on disease risk. As a consequence, the causal genes for a rare disorder can often be identified by sequencing individual patients or families. In contrast, the genetic risks for common diseases are modulated by a large number of mostly non-coding variants that individually exert small effects. These variants are typically identified through genome-wide association studies (GWASs). However, identification of the causal variants and genes affected by GWAS loci remains challenging, in part due to linkage disequilibrium (LD) and small effect-sizes ^1, 2^.

Despite the differences between rare and complex diseases, it has been shown that GWAS loci for multiple traits are enriched for genes that can cause related rare diseases when damaged ^3, 4^. For instance, common variants associated to PR interval, a measurement of heart function, have been found within the *MYH6* gene ^5^, which is known to harbour mutations in individuals with familial dilated cardiomyopathy ^6^. Moreover, eQTL studies have found examples of rare disease genes that are affected by distal common variants in *trans*, such as the immunodeficiency gene *ISG15,* which is affected by multiple systemic lupus erythematosus–associated variants ^7^. These results indicate that common and rare diseases can result from damage to or altered regulation of the same genes, suggesting that the same biological pathways underlie these conditions ^4^. However, it is not fully known to what extent specific genes and pathways are shared between rare and common diseases.

Over the years, many pathway-enrichment methods have been developed that can identify which biological pathways are enriched for common diseases ^8–10^ as well as highlighting their most likely cellular context(s) ^11, 12^. In addition, several methods can prioritize individual genes within GWAS susceptibility loci by studying how they are functionally related to genes in other susceptibility loci ^8, 13–16^. However, these methods confine themselves to genes in GWAS loci, potentially missing relevant *trans-*regulated up- or downstream effects. In blood, expression quantitative trait locus (eQTL) mapping has been successful in identifying the downstream *trans* regulatory consequences of GWAS-associated variants (i.e. *trans*-eQTLs and eQTSs, where polygenic scores are linked to expression levels) ^7^. Unfortunately, large eQTL sample sizes are required to detect such effects, and such datasets are not yet available for most tissues.

Here we build upon the ‘omnigenic model’ hypothesis, which states that the genes that are most important in complex diseases are those that are modulated by many different common variants through gene regulatory networks ^17, 18^. The omnigenic model postulates that a limited number of core genes exist that drive diseases, but that many peripheral genes, which contain associated variants, contribute indirectly to disease development by modulating the activity of the core genes. Since the omnigenic model predicts that many core genes map outside GWAS loci, these genes will be missed by methods that prioritize genes within GWAS loci. The omnigenic model hypothesis is supported by recent works assessing RNA levels of blood cells ^19^ and molecular traits ^20^ and a large-scale *in vitro* knockdown experiment ^21^. However, these studies were performed in blood, limiting their conclusions to GWAS studies on blood-related traits and immunological disorders.

To take this work further, we integrated (mRNA level) gene regulatory networks with GWAS summary statistics to prioritize key genes that we suspect are more likely to directly contribute to disease predisposition than genes in GWAS loci. We have implemented this strategy in a software package called ‘Downstreamer’ that uses GWAS summary statistics and gene co-regulation based on 31,499 multi-tissue RNA-seq samples in order to prioritize these key genes. We also provide pathway, rare disease phenotype (coded by HPO terms) and tissue enrichments to aid in the comprehensive interpretation of GWAS results.

We applied Downstreamer to 44 GWASs for a wide variety of traits (Table S1) and show that the identified key genes are enriched for intolerance to loss-of-function (LoF) and missense (MiS) mutations and for Mendelian disease genes that lead to similar phenotypic outcomes as the GWAS trait. Specifically, we find that key genes for height are strongly enriched for severe growth defects and skeletal abnormalities in humans and mice. Additionally, key genes for auto-immune diseases point to lymphocyte checkpoints and regulators and those for glomerular filtration rate (GFR; a measure of kidney function) are transporters for several metabolites.

Key genes that cause Mendelian disease can therefore highlight the molecular pathways driving the complex disease. Conversely, predicted key genes may aid in identifying new Mendelian disease genes.

## Results

To enable identification of GWAS key genes, we developed Downstreamer (Methods), a tool that integrates GWAS summary statistics with gene expression–based co-regulation networks. Downstreamer first converts individual variant associations to gene p-values by aggregating associations within a 25kb window around the gene body for all protein-coding genes while correcting for LD between variants ^9^ (Fig. 1A). These gene p-values are then converted to z-scores. We calculated gene z-scores for 44 GWAS summary statistics reflecting a wide variety of disorders and complex traits (Table S1).

**Fig. 1.**
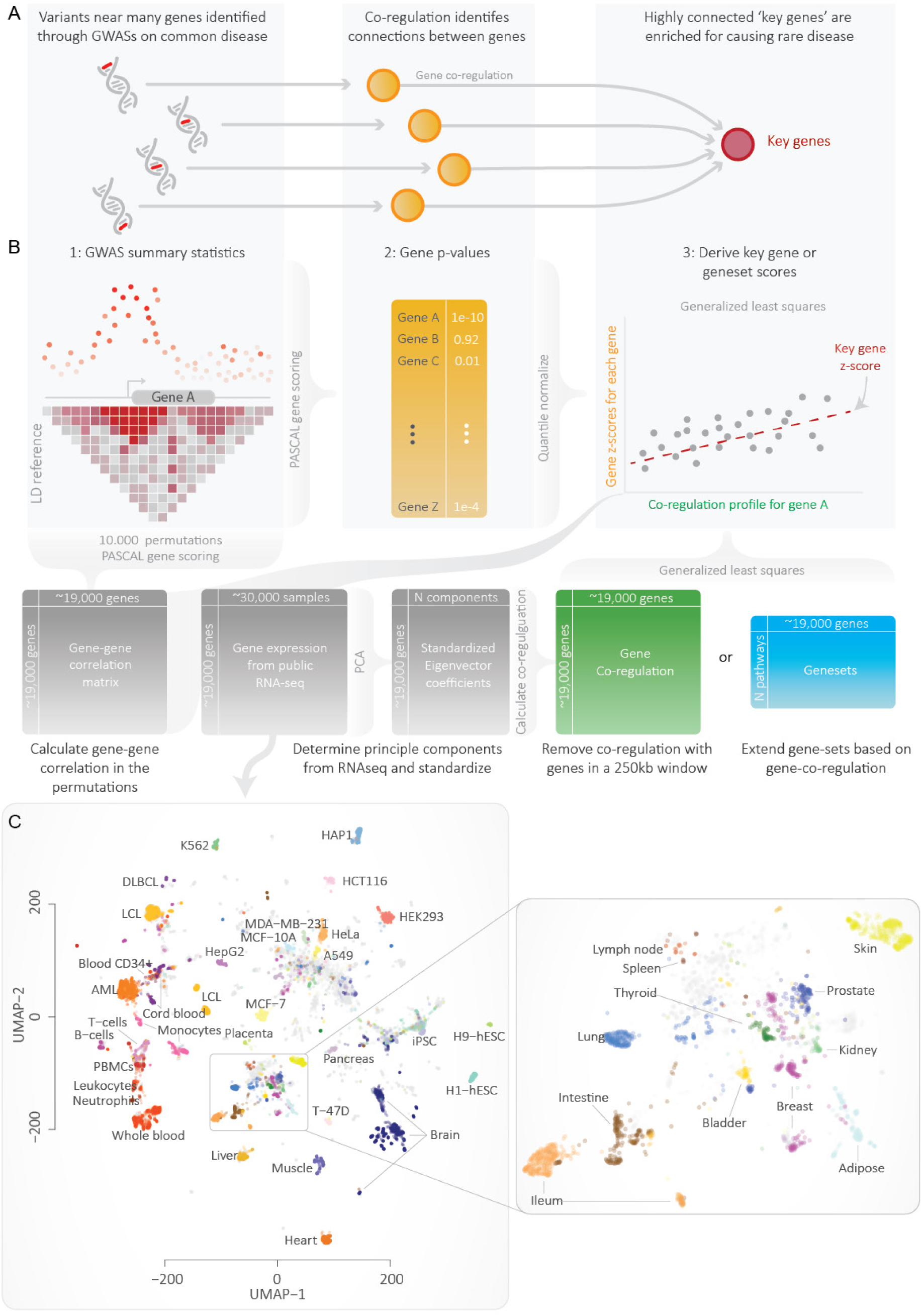
A) Downstreamer works on the idea that many genes identified through GWAS jointly affect a set of key genes that strongly impact disease development. B) Schematic overview of Downstreamer methodology. C) The 31,499 RNA-seq samples used for the study visualized using Uniform Manifold Approximation and Projection (UMAP). The zoom-in on the right shows a detailed view of the various primary tissues in the dataset.

### Association signals for polygenic traits cluster around transcription factors

We observed that the z-scores of individual GWASs were often weakly positively correlated (Fig. S1A), especially for traits for which many loci have been identified (Fig. S1 B). For instance, the gene-level z-scores for height correlated positively with the gene-level z-scores of all other traits. To investigate the source of this shared signal, we calculated the average gene-level significance across all 44 traits while correcting for bias that might be introduced for traits that are strongly correlated (see Methods).

We observed that 30% of the variation in this ‘average GWAS’ signal could be explained by both the extent of LD around a gene and by the local gene density (Fig. S2). The more extensive the LD around a gene, the higher the chance that genetic variants within the gene are associated, especially for highly polygenic traits ^22, 23^. Consequently, the gene-level z-score for these genes increases. Hence, when collapsing GWAS summary statistics into gene z-scores, some amount of correlation between well-powered GWAS studies is to be expected. However, this is unwanted when using gene z-scores in a pathway-enrichment analysis. We next evaluated if the remaining 70% of the average signal was enriched for any biological processes. After correcting for LD and gene density, we observed that 79 of the top 500 genes are transcription factors (OR: 2.22, p-value: 4.25×10^-9^). We also saw enrichment among the top 500 genes for pathways related to DNA binding and transcription, for example, transcription regulator activity (OR: 1.98, p-value: 2.24×10^-11^) (Table S2). Additionally, genes with a higher average gene z-score were enriched for intolerance to LoF (Fig. S3). These enrichments suggest that there is a set of genes, enriched for GWAS hits, that confer risk to many different types of traits. This is consistent with previous observations that broad functional categories tend to be enriched for many traits^17^.

However, as these often-associated genes obscure the specific pathways and key genes for a trait, we corrected the individual gene z-scores for the average gene z-score in order to get disease-specific gene-level significance scores that were as specific as possible. Downstreamer then correlates these corrected gene z-scores to gene expression patterns, pathway memberships and tissue expression through a generalized least squares (GLS) model that accounts for gene–gene correlations resulting from the relationship between LD and sharing of biological functionality (Fig. 1B, Methods). This results in a z-score that represents the significance of the association of a gene, pathway or tissue

### Identification of key genes using gene co-expression

To identify key genes, we searched for genes that are co-regulated with the genes within a given trait’s GWAS loci. We used a gene expression database containing 31,499 tissue and cell-line RNA-seq samples ^24^ to calculate gene co-regulation (a measure of the similarity of expression) for each pair of protein-coding genes (Fig. 1, Methods). Co-regulation is defined as the correlation between standardized eigenvector coefficients derived from the expression data (Fig. 1B). Since each component is given equal weight, co-regulation is less sensitive to the major tissue effects that can confound co-expression correlations calculated using expression data from a set of heterogeneous tissue samples. To ensure that no bias was introduced by GWAS loci located in highly co-regulated gene clusters, co-regulation relationships between genes within 250kb were removed to further compensate for these gene clusters resulting from genomic organisation. We then associated the gene z-scores to gene co-regulation using a GLS model. We use permutations to determine the significance of the association. The resulting association z-score reflects the overall connectivity of that gene to the GWAS genes in the network (Fig. 1B, Methods). We call this z-score the ‘key gene score’ throughout the manuscript, and we call the genes that pass Bonferroni significance and have a positive association ‘key genes’. Besides detecting key genes, Downstreamer is also able to identify pathway and tissue enrichments for GWAS traits, using reconstituted gene sets that provide increased statistical power to identify significant pathway enrichment (Note S1, Note S2). Pathway and tissue enrichments results yielded plausible results consistent with previous findings, indicating that correction for the average GWAS signal is a useful addition.

In total, we identified 3,648 key genes over the 44 tested traits, with most key genes arising from GWASs for white blood cell composition and other haematological factors (Fig. 2A). The number of samples and independent loci for a GWAS is positively correlated to the number of detected key genes (Pearson R: 0.38 and 0.33, p-values: 1×10^-2^ and 2.8×10^-2^, respectively; Fig. S4), which is to be expected as larger GWASs typically contain more signal. To determine how similar the key gene predictions are, we correlated the key gene scores of the 44 different traits to each other and observed that traits of the same class cluster together (Fig. 2B). For example, the immune diseases (inflammatory bowel disease (IBD), coeliac disease (CeD), type 1 diabetes (T1D), rheumatoid arthritis (RA), asthma and multiple sclerosis (MS)) clustered together, neighboured by traits representing white blood cell composition. Other distinct clusters were found for psychological traits (educational attainment, schizophrenia, major depressive disorder, body mass index (BMI)) and cardiovascular traits (pulse pressure, diastolic and systolic blood pressure, coronary artery disease), further showing that the gene regulatory networks downstream of GWAS signals are partially shared between related traits. To some extent, this sharing is expected, given known co-morbidities between, for example, CeD and T1D ^25^ and the widespread genetic correlations of related complex traits ^26^.

**Fig. 2.**
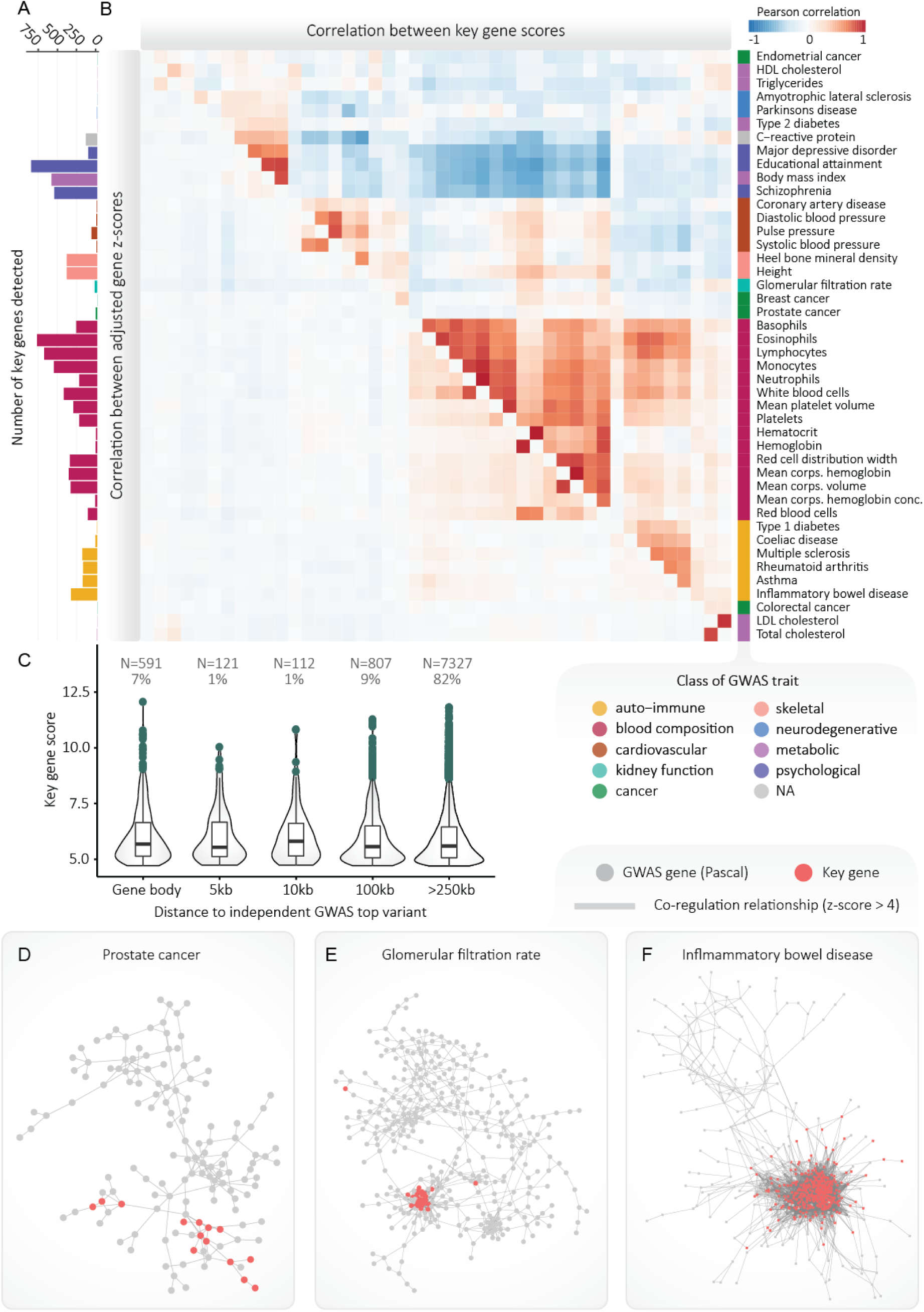
A) Number of key genes detected for each GWAS tested. B) Pearson correlations between the gene z-scores after correction for the mean signal (lower triangle) and the key gene scores (upper triangle). Correlations were calculated using all protein-coding genes (including non-significant ones). C) Boxplots showing key gene scores in relation to the independent significant top SNPs from the GWAS. D) The gene regulatory network for prostate cancer. The network shows how prostate cancer GWAS genes and key genes are interconnected. Grey nodes represent GWAS genes. Red nodes indicate key genes. A key gene may also be located in a GWAS locus. Only positive co-regulation relationships with a z-score >4 are drawn as an edge in the network. E) As D, but showing the network for the GWAS and key genes for glomerular filtration rate. F) As D, but showing the network for the GWAS and key genes for inflammatory bowel disease.

On average, 82% of predicted key genes are located outside GWAS loci (≥250kb from the lead variant) (Fig. 2C). This indicates that the key genes may be under *trans* regulation by the genes within the GWAS loci, rather than being impacted by a GWAS variants directly in *cis,* as is the case for most genes in GWAS loci. The other 18% of key genes are located within GWAS loci, suggesting that there is both a *cis* effect by a genetic variant that directly perturbs the function of these genes and a *trans* effect where the other GWAS loci modulate these genes.

Of note, out of the 3,648 key genes detected, 2,036 (55%) were detected in multiple GWAS traits. However, this number is largely driven by the genes we detected for highly correlated traits such as blood cell composition (Fig. 2A, Fig. S5). To better determine if key genes are trait-specific or shared among different diseases, we assigned each of the 44 traits to 10 broader classes such as auto-immune disease or blood cell composition. We then observed that 1,032 (28%) of the key genes are shared between at least two different classes (Fig. S5A). This is largely driven by the overlapping key genes of blood cell composition and auto-immune disease, which account for 413 of the 1,032 overlapping key genes.

Below we highlight key genes for a few traits. For prostate cancer, we prioritized 14 key genes, 10 of these are outside the GWAS loci (Fig. 2D). The most notable are *KLK3,* which encodes for PSA (prostate-specific antigen), the marker that is used to screen for prostate cancer ^27^, and *KLK2,* which is known to activate *KLK3*^28^. Additionally, many other key genes we identified have either been implicated in prostate cancer^29–32^ (*TMC5*, *MLPH, OVOL2* and *CHD1*) or in other types of cancer (*TFAP2C*, *BAIAP2L1* and *PLEKHN1)*^33–35^.

The GWAS for GFR, a measurement of kidney function, revealed 32 key genes (Fig. 2E), of which 6 genes are solute carriers (a group of membrane transporters). Two of these, *SLC22A12 and SLC17A1*, are known to be urate transporters, fitting the known relationship between urate levels and GFR ^36^. Other notable GFR key genes include four glucuronosyltransferases (*UGT1A9*, *UGT2B7*, *UGT2A3* and *UGT1A6)* that are important in drug metabolism and clearance of drugs by the kidneys ^37–39^ and four genes (*UMOD, SLC22A12, SLC36A2* and *NPHS2)* known to cause rare forms of kidney disease ^40^.

The auto-immune diseases shared several key genes, such as *IL-2RA*, *ICOS* and *CD48*, indicating an adaptive immune signature. Recently, a large-scale CRISPR assay assessing the regulators of *IL-2*, *IL-2RA* and *CTLA4* systematically knocked down thousands of immune genes in primary CD4+ T cells in order to assess how these genes are co-regulated ^21^. The genes regulating *IL-2RA* formed a highly inter-connected network, with the members of this network being significantly enriched for harbouring GWAS signals for MS. We prioritize *IL-2RA* as one of the most significant key genes for MS, but *IL-2RA* is also a key gene for IBD, asthma, CeD, RA and white blood cells, consistent with the role of T cells in these diseases. For IBD, low-dose IL-2 has been shown to alleviate symptoms of DSS-induced colitis in mice, highlighting this pathway as a potentially viable therapeutic target ^41^. Additionally, a duplication found in the *IL-2RA* locus, causing excessive IL-2 signalling, may predispose carriers to early-onset colitis ^42^.

Among the key genes identified for IBD (Fig. 2F), there are several known drug targets. Some targets were already identified through GWAS (e.g. *TNF*, *JAK2* and *PRKCB)*. Others are located outside the GWAS loci, including *ITGB7,* which is one of the targets of Vedolizumab ^43^; *JAK3,* which is targeted by JAK inhibitors ^44^ and *S1PR4* which is targeted by Amiselimod ^45^. Additionally, *RGS1* has been proposed to be a druggable target that protects against colitis when downregulated ^46–48^. While *RGS1* has been associated to CeD ^49^ and MS ^50^, its loci were not identified by the IBD GWAS.

Similarly, for schizophrenia, we identify key genes within GWAS loci that are targeted by schizophrenia drugs (*RM3*, *GRIA1* and *GRIN2A),* as well as key genes located outside of the GWAS loci that are established drug targets or being tested as drug targets. These include *HTR1A,* which is target of aripiprazole ^51^; *HTR5A* and *HTR1E,* which are both targeted by amisulpride ^52^ and *GRIA2* and *GRIA3*, which are both targets of topiramate ^53^

### Key genes can be depleted or enriched for cis-eQTL effects

It has been observed in blood that genes without a detectable *cis*-eQTL effect are more intolerant to loss-of-function mutations ^7^. This is possibly explained by more extensive buffering of regulatory effects on these important genes ^54^. This has implications for the use of *cis-*eQTLs to identify disease-relevant genes. Here, we assessed whether key genes have fewer *cis*-eQTL effects than expected by chance. We did not observe consistent depletion of *cis-*eQTL. When testing 28 traits for which Downstreamer predicted at least 10 key genes, using blood-based *cis-*eQTLs from the eQTLgen consortium (Fig. 3A), we found Bonferroni significant enrichments for five traits (IBD and four different white blood cell count measurements, ORs: 2.36 – 2.93, p-values: 1.31×10^-3^ – 2.45×10^-7^). Two traits were significantly depleted for blood *cis*-eQTLs: C-reactive protein levels (OR: 0.19, p-value: 4.68×10^-4^) and BMI (OR: 0.47, p-value: 1.19×10^-3^) (Fig. 3A). When using brain-based *cis*- QTLs from the MetaBrain project ^55^, we found three traits for which the key genes are significantly depleted for being *cis-*eQTLs: educational attainment (OR: 0.69, p-value 7.40×10^-7^), BMI (OR: 0.63, p-value 1.17×10^-7^), and schizophrenia (OR: 0.61, p-value 4.54×10^-8^) (Fig. 3B), and observed no significant enrichment.

**Fig. 3.**
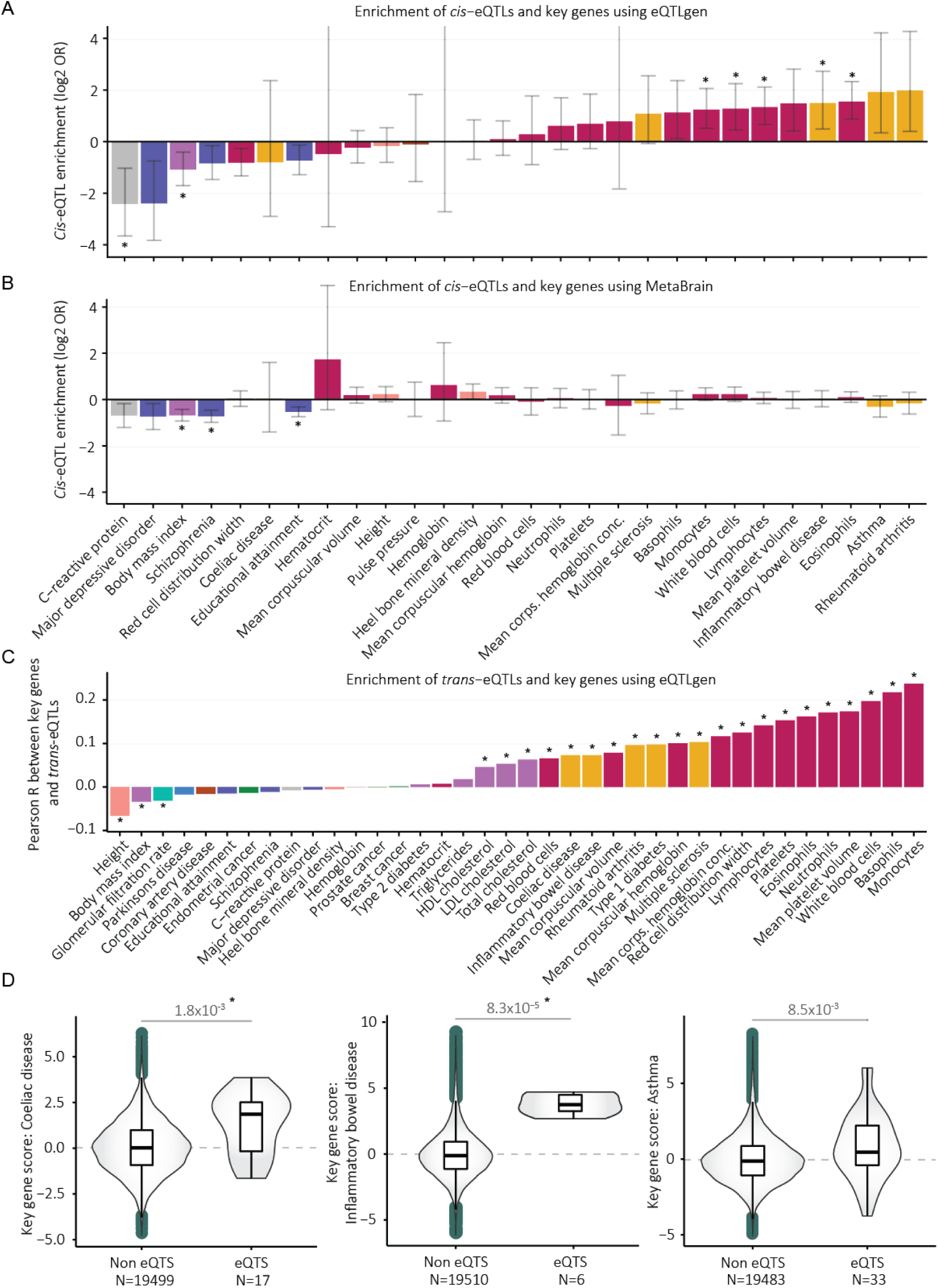
A) Enrichment of cis-eQTL genes and key genes. Cis eQTL genes are genes that have a significant cis eQTL effect in eQTLgen. * Indicates Bonferroni-adjusted p-values < 5e-2 corrected for 28 tests. B) As A, but using MetaBrain cis eQTLs. C) Pearson correlation coefficients between key gene scores and the sum chi square statistics (see Methods) of the trans eQTL effects from significant independent GWAS SNPs. * Indicates Bonferroni-adjusted p-values < 5e-2 corrected for 44 tests. D) Key gene scores for genes found to be in eQTS in the eQTLgen consortium for CeD, IBD and asthma. Nominal p-values of a t-test between the eQTS and non-eQTS groups are indicated. * Indicates Bonferroni-adjusted p-values < 5e-2 corrected for 11 tests.

### Overlap between key genes and trans regulatory targets

To investigate if the key genes result from *trans* regulation originating in the GWAS loci, we assessed if there was a correlation between the key gene scores and *trans*-eQTL effects from the eQTLgen consortium ^7^. To do so, for each gene, we summed the squared z-scores of *trans*-eQTL effects from the independent top hits for each GWAS. This results in a chi-square score for each gene that depicts to what extent the top GWAS variants of a trait affect the expression of those genes. We then correlated these scores with key gene scores. We found a significant association between the Downstreamer key gene scores and chi-square statistics of the eQTL effects for 24 of the 44 traits (Bonferroni-adjusted P≤0.05, Fig. 3C). Not surprisingly, the strongest associations were for the GWASs representing blood cell traits and auto-immune diseases. Interestingly, three non-blood traits – height, BMI and GFR – displayed significant negative correlations, which suggests the unsuitability of blood *trans*- eQTLs for interpreting non-blood traits, likely due to low expression of the relevant genes in blood ^7^.

Since *trans*-eQTL effects are typically small and current datasets are only powered to detect a fraction of these effects ^7^, the eQTLGen consortium correlated the polygenic scores for a diverse set of traits to gene expression levels. The genes with significantly lower or higher expression depending on the polygenic scores of the individuals (so-called eQTS genes) were prioritized as relevant for the trait. We observed that eQTS genes had higher key gene scores for three traits (T-test p-values, IBD: 1.8×10^-3^, CeD: 8.3×10^-5^ and asthma: 8.5×10^-3^), suggesting that key genes are more likely to be influenced by converging trans-eQTL effects (Fig. 3D).

We identified two genes that were both key genes and eQTS genes for asthma: *RELB* and *CST7*. *RELB* is a member of the NF-κB family of transcription factors that activate the non-canonical NF-κB pathway ^56^ and has been linked defects in T and B cell maturation, leading to combined immunodeficiency and auto-immune responses ^57^. *CST7* has been described as a critical factor in maintaining eosinophile function ^58^, and eosinophiles are known to be one of the key cell types in asthma ^59^.

### Key genes tend to be highly expressed in relevant tissues and cell types

As GWASs in the same class tended to show enrichment in the same cell types (Data S1, Fig. S7) and shared key genes, we next tested if key genes were highly expressed in the cell types relevant for the corresponding trait. To determine the tissue specificity of each gene for a given tissue, we calculated a statistic for how highly a gene is expressed in that tissue by subtracting the mean expression of the samples of that tissue from the mean of all other samples in our dataset. This revealed significant association between the key gene scores and the expression level of genes in seemingly relevant tissues (Fig. 4, Fig. S8, Fig. S9, Fig. S10), highlighting that the key genes tend to be highly expressed in the cell types where the GWAS is most enriched.

**Fig. 4.**
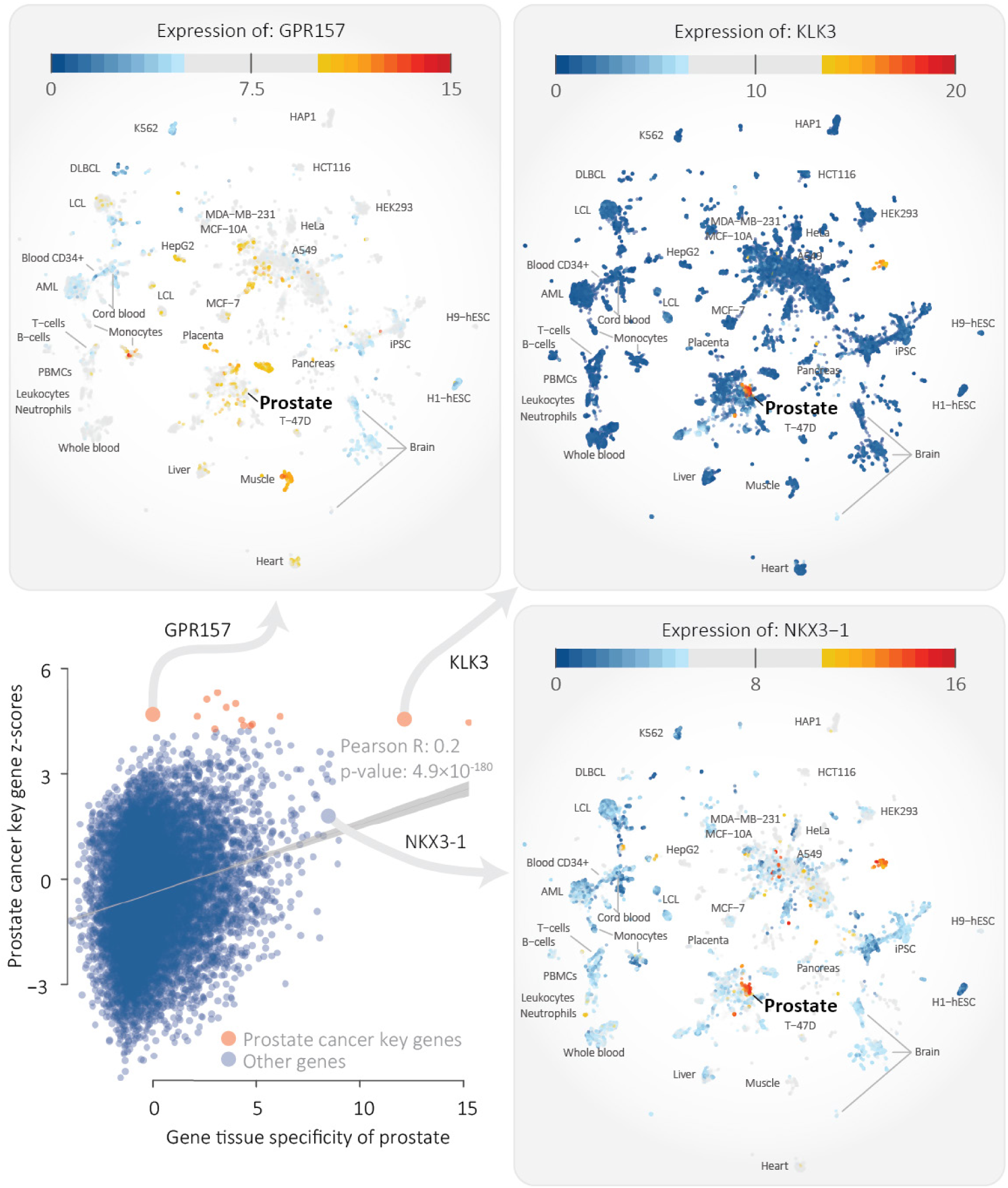
Scatterplot showing the specificity of expression of a gene in prostate (x-axis) versus the key gene z-score of prostate cancer (y-axis). The Pearson correlation is 0.2 (p-value: 4.9×10^-180^). Specificity of expression was determined by taking the mean in prostate samples and subtracting the mean from all other annotated samples in the dataset. The three panels show the expression of the highlighted genes, revealing that some key genes are specifically expressed in prostate but there are also key genes that are not prostate-specific and, vice-versa, there are prostate-specific genes that are not predicted to be key genes.

For example, several key genes for prostate cancer, such as *KLK3* (coding for the prostate-specific antigen), are highly expressed in prostate (Fig. 4). However, we also identified key genes for prostate cancer, such as *GPRS158,* that showed average expression in prostate but were much more highly expressed in other tissues such as muscle. Additionally, we also observed genes that were highly expressed in the prostate that were not key genes, such as *NKX3-1*. We saw similar examples for GFR and IBD (Fig. S9, Fig. S10). This indicates that the key gene prioritizations are in all likelihood not purely driven by tissue specific expression.

### Key genes are more often constrained

We next reasoned that if key genes are essential to fundamental biological processes, they would be more likely to be more evolutionarily constrained. We therefore compared the key gene scores to the MiS and LoF z-scores from gnomAD ^60^. These z-scores describe the tolerance level of a gene to MiS or LoF variants. The higher the score, the less tolerant the gene is to these types of variants. We observed significant association between the key gene scores and the MiS (Pearson R: 0.12, p-value: 3.7×10^-57^) and LoF (Pearson R: 0.13, p-value: 1.4×10^-7^) z-scores (Fig. S11). Compared to the key genes, genes that map within GWAS loci had a lower LoF association (Pearson R: 0.07, p-value: 1.2×10^-05^), but similar association with MiS (Pearson R: 0.12, p-value: 1.3×10^-12^). Next, we evaluated if this association was driven by genes that have more connections in the gene network (i.e. whether a gene is a ‘hub’ gene or not), as we observed that the number of connections a gene has in the network is associated to the key gene score (Fig. S13). After correcting for the number of connections a gene has, the associations for MiS and LoF remained, but were reduced (Pearson R: 0.07, 0.08, p-value 6.31×10^-21^ and 3.9×10^-24^, respectively). Together, these results suggest that key genes tend to be evolutionarily constrained and are especially intolerant to LoF variants compared to the PASCAL gene p-values.

### Key genes are enriched for Mendelian genes for related phenotypes

Genes in GWAS loci are known to be enriched for causing Mendelian diseases ^3^. We observe that these enrichments of Mendelian disease genes are even stronger for the key genes that we prioritize. For example, we identified 398 Bonferroni-significant key genes for height and 90 (22.6%) of those are Mendelian disease genes causing “Abnormality of the skeletal system” (p-value: 5.18×10^-9^, OR: 2.13, Data S4). This enrichment is stronger than for genes in the GWAS loci, where 319 of the 1,951 (16.4%) genes are annotated to cause “Abnormality of the skeletal system” (p-value: 7.86×10^-9^, OR: 1.47, Data S2). Even when only considering the closest gene near the lead height GWAS hits, the enrichment of key-genes remains stronger (p-value: 7.02×10^-12^, OR: 1.85, Data S3)

The most significant enrichment for height key genes is for “Abnormal lower limb bone morphology”: 43 key genes are annotated to this HPO term (p-value: 6.86×10^-15^, OR: 4.71). When also considering phenotypes based on mouse orthologs, we found 78 that are associated to growth and 128 that are pre- or post-natal lethal. In total, we can hereby explain 171 of the 398 (43%) associated height genes (Fig. 5A). These key genes are enriched for various pathways (Fig. 5B) including ‘Collagen fibril organisation’, ‘Embryonic digit morphogenesis’ and ‘Extracellular matrix organisation’.

**Fig. 5.**
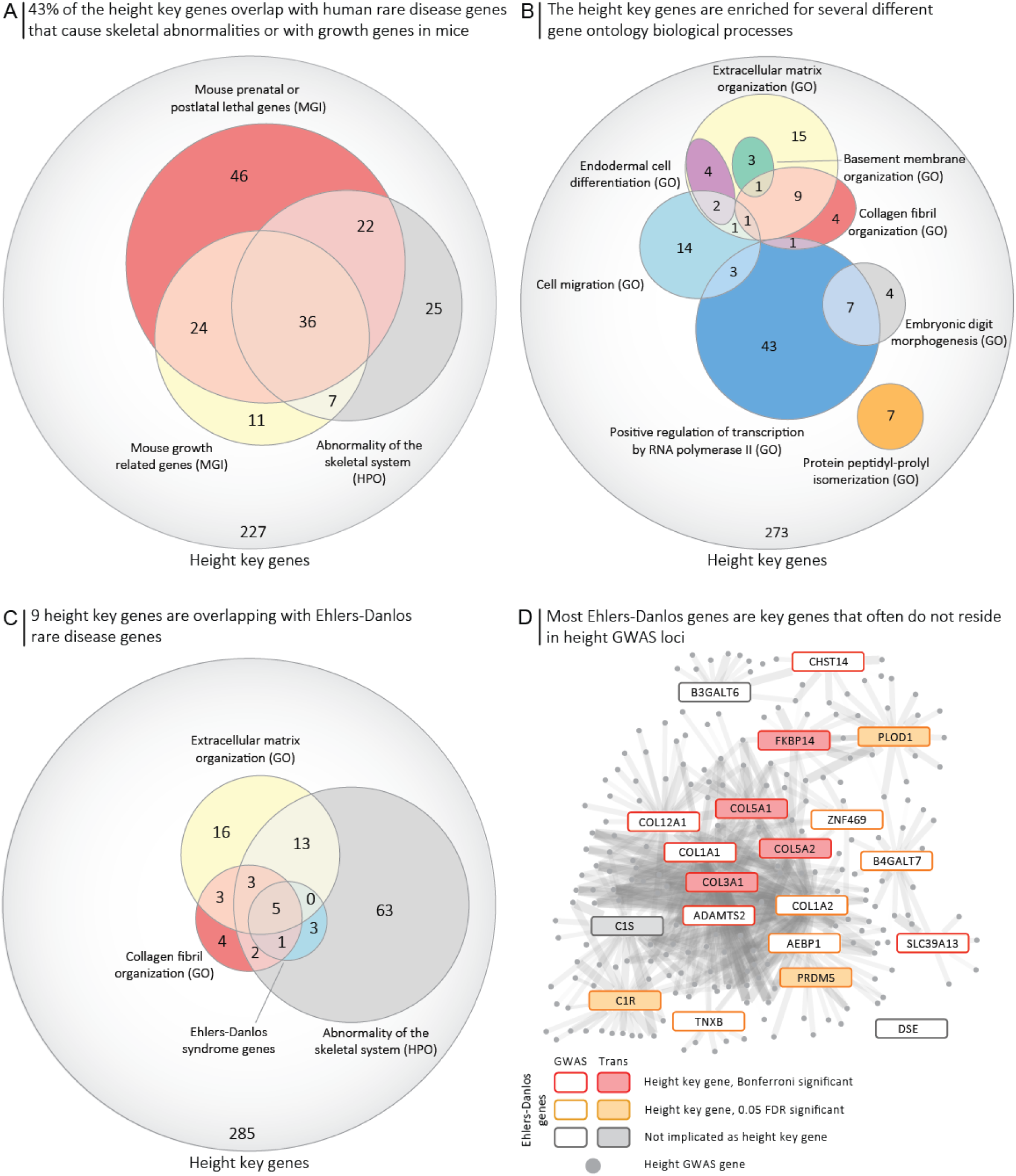
A) 43% of height key genes are known to result in growth abnormalities in either humans or mice, indicating that these key genes are important genes for height. B) The height key genes are enriched with different, only partly overlapping, pathways, indicating that the key genes are part of multiple biological processes. C) Nine of the height key genes are known to cause Ehlers-Danlos syndromes, which involve abnormalities of the skeletal system. Most of these are annotated to the GO pathways for “Extracellular matrix organization” and “Collagen fibril organization”. It may be that the key genes that we now link to height and that are part of the collagen or extracellular matrix pathways also contribute to Ehlers-Danlos syndromes. D) The Ehlers-Danlos genes are co-expressed with many height GWAS loci genes.

Among the Bonferroni-significant key genes for height are 9 of the 21 known Ehlers-Danlos genes (Fig. 5C, D). When using a less stringent FDR of 5%, we predict 17 out of the 21 Ehlers-Danlos genes to be key genes for height. Ehlers-Danlos syndromes are disorders of the connective tissues that often result in skeletal malformities ^61^. These syndromes are caused by defects in, or related to, the collagen formation needed for the extracellular matrix. This is in line with the pathway enrichments of the height key genes and earlier findings ^62^.

Three genes with a Bonferroni significant gene p-value for IBD (*SKIV2L*, *NOD2*, *RTEL1*) overlapped with the 36 HPO-annotated colitis genes (OR: 4.87, p-value: 2.8×10^-2^). The enrichment for colitis genes improved when assessing the key genes, which increased the overlap to six genes (*IL10RA*, *RASGRP1*, *NCF4*, *TNFAIP3*, *FASLG*, *ZAP70;* OR: 11.46, p-value: 3.3×10^-5^). *IL10RA*, *RASGRP1*, *NCF4* and *ZAP70* were all located further than 250kb from an independent GWAS hit in the GWAS used, meaning that these genes would not have been identified by overlapping IBD GWAS loci with known Mendelian genes. Other phenotypes that were significantly enriched among the key genes for IBD included those related to recurrent (fungal) infections and various phenotypes relating to immune function. Enriched mouse phenotypes included many related to T and B cell function and abundance (Data S1).

We matched each of the 44 GWAS traits to a best-fitting HPO term based on the phenotypic descriptions (Table S3). We observed that 22% of the identified key genes are linked to rare diseases that cause related phenotypes (Fig. 6A, Table S4). We found that the key genes for 18 of the 44 traits are significantly enriched (adjusted for 44 tests) for related rare disease genes (Fig. 6B; p-values: 2.98×10^-4^ to 4.91×10^-29^, OR: 1.79 to 71.83). Another eight traits showed nominally significant overlap between key genes and related rare disease genes. For the 18 traits without significant overlap, we found between 0 and 20 key genes, indicating that our power for these traits was limited (Fig. 6D). The only exception to this was the GWAS for c-reactive protein levels, for which we found 148 key genes but only 6 genes were linked to its HPO term. Despite the limited power for these 18 traits, 9 traits had significantly larger key gene scores for the HPO-associated genes (U-test p-values: 8.73×10^-12^ to 8.28×10^-4^), indicating that the key gene scores still have some predictive power for detecting rare disease genes (Fig. 6C).

**Fig. 6.**
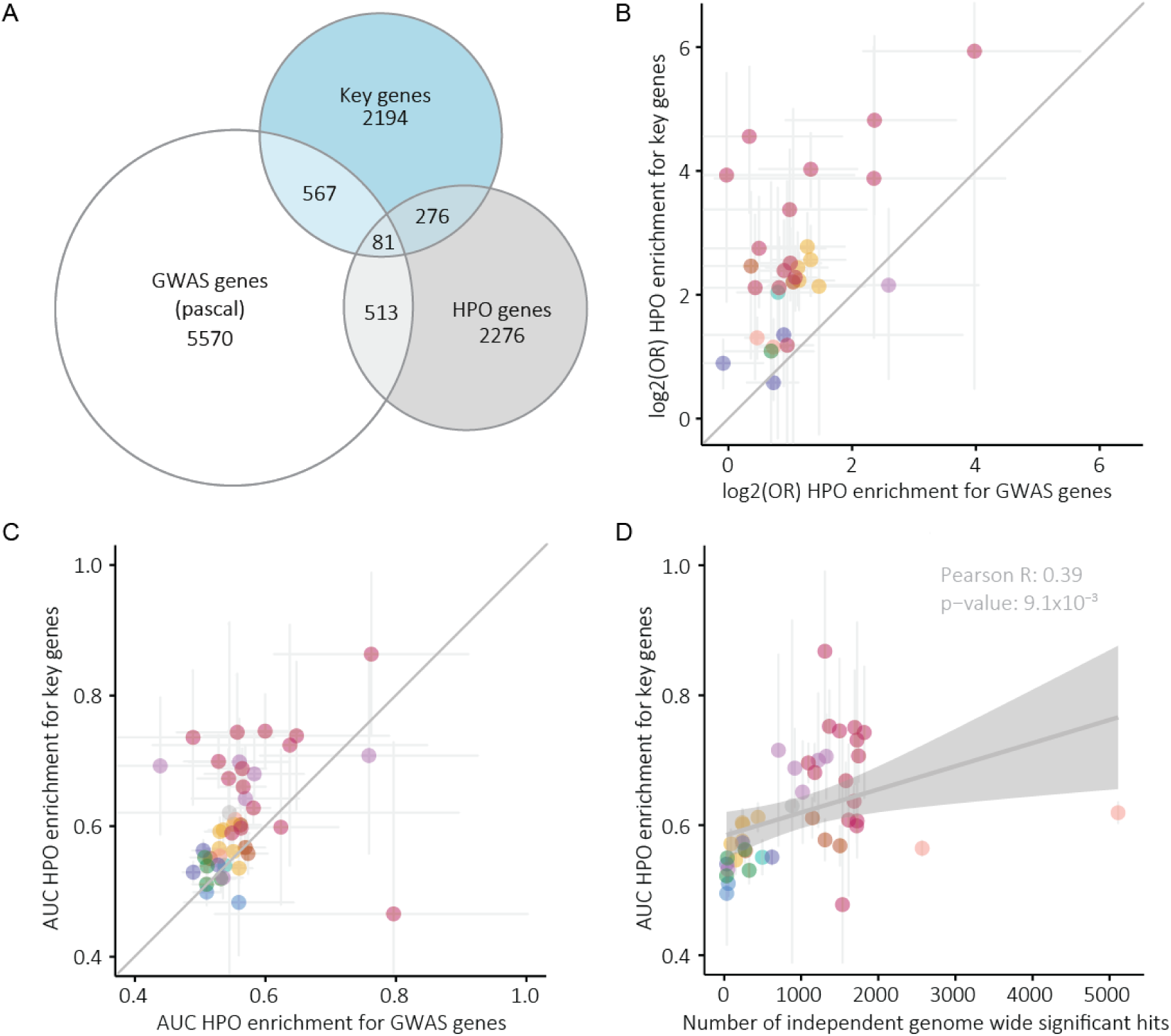
A) Overview of the overlap between key genes, the genes for the 44 HPO terms we matched to their respective GWAS and the Bonferroni-significant genes identified by PASCAL. B) Comparison between the odds ratios of the HPO enrichment done using key genes and Bonferroni-significant GWAS genes identified by PASCAL. Each dot represents the HPO term matched to that GWAS. 95% confidence intervals of the odds ratios are represented. C) As in B but showing the AUC values calculated using the entire key gene z-score or GWAS gene p-value vector for all protein-coding genes. D) Association between the AUC values and the number of genome-wide significant hits for each GWAS.

## Discussion

In this work, we present Downstreamer, a method that integrates gene co-regulation with GWAS summary statistics to prioritize genes central in the respective trait’s network. We applied Downstreamer to 44 GWAS studies and prioritized genes that are not directly implicated by GWAS, yet are good candidates based on pathway annotation and their involvement in Mendelian diseases. Some of the genes showed evidence of being directly regulated by *trans*-acting genetic factors. The key genes are enriched for being evolutionarily constrained, indicating they more often have crucial biological functions. These findings suggest that the small effects of GWAS-associated variants ultimately converge on key disease genes.

We observed that the gene prioritization scores of related traits are often correlated (Fig. 2B). To some extent this is expected given the known shared genetic signature of, for instance, auto-immune disorders. It could also potentially indicate that the gene prioritizations are confounded by cell type–specific expression levels. Indeed, we found the genes prioritized for a trait to be more abundantly expressed in the samples that best match that trait (Fig. 4, Fig. S8). However, this was not the sole driver of our key gene prioritization. We also found several examples of key genes that are not specifically expressed in the relevant tissues, as well as genes with very low key genes scores that show similar tissue-specific expression to the key genes. For instance*, GPR157* is predicted to be a key gene for prostate cancer, but it is highly expressed in many tissues (Fig. 4, Fig. S9, Fig. S10). Additionally, high expression of key genes in the disease-relevant tissue is to be expected because rare disease genes are also highly expressed in the tissue relevant to those diseases ^63^.

We recently also applied a pre-release version of Downstreamer to several neurodegenerative diseases, while using a comprehensive brain-specific gene co-regulation network of the MetaBrain project. This revealed that the signal of underrepresented cell types and tissues can be overshadowed by more abundant tissues in our expression data ^55^. This might be especially relevant for diseases in which uncommon or rare cell types are instrumental to disease pathophysiology, such as gluten-specific T cells in CeD ^64^. We therefore expect that key gene prioritizations can benefit from creating tissue-or even cell type–specific gene regulatory networks, should enough samples be available for the relevant tissue or cell type to accurately calculate co-expression. The future generation and inclusion of single-cell RNA-seq data should also be able to solve the issues regarding confounding by cell-type composition.

We observe that genes with *cis-*eQTL effects in blood are enriched for being key genes for blood traits. For instance, for IBD the key genes are more likely to be blood eQTLs than is expected by chance (Fig. 3A). A similar enrichment is seen at nominal significance levels for rheumatoid arthritis and asthma. Different results were found when using brain-derived *cis-*eQTLs. Using MetaBrain eQTLs, we found a depletion of *cis-*eQTL genes among the key genes of several brain related traits (Fig. 3B). This might indicate that genes that are important for the brain are more tightly controlled and are therefore not as easily affected by eQTL effects compared to important immune genes.

When comparing our prioritized genes to the results of a large blood-based *trans*-eQTL and eQTS analysis, we found an overlap in identified genes (Fig. 3C). As expected, this primarily holds for traits manifesting in blood, such as immune disorders and blood cell proportions. This confirms that a portion of the genes identified by Downstreamer are modulated by disease-associated variants in *trans*. We suspect four main causes for why there are key genes for which we cannot confirm *trans* regulation using blood-based *trans*- eQTLs or eQTSs: 1) the gene is not expressed in blood or the regulation is not present in blood, 2) the effects of genetic variants that only act in rarer blood cell types that are diluted by the expression levels in the more common cell types ^65^, 3) some trait-associated eQTLs depend on specific environmental stimuli ^66^, which can hinder the ability of population cohort studies to identify the regulatory consequences of disease-associated variants, and 4) Downstreamer works by integrating the many small effects originating from many different loci. Individually these effects might be too weak to currently be detected as *trans*-eQTL effects. We therefore conclude that co-expression-based methods such as Downstreamer are complementary to existing studies that link disease-associated variants to gene profiles.

One assumption we make when calculating the gene p-values is that the genes within 25kb of a GWAS signal are affected by the GWAS variants, but this might not the case for all genes. However, recent work has suggested that except for integrating epigenetic and HiC contact data, the next best predictor of causality is the closest gene to the top of the GWAS signal and this approach outperforms eQTL-based approaches ^67^. We therefore decided not to integrate any prior eQTL information when calculating gene p-values, as this would often lead to incorrect prioritizations. In addition, the genes affected by GWAS variants are also likely to be tissue-specific, further complicating the prioritizations, and we would need extensive prior information to select the correct eQTLs or epigenetic information. This presents an area where major improvements could be made in future, when more accurate and systematic predictions can be made about which genes are regulated by GWAS variants in *cis*.

Our findings are in line with the infinitesimal model ^68^ that postulates that a quantitative trait or complex genetic disease can result from an infinite number of variants, each exerting an infinitely small effect size. An extension of the infinitesimal model is the omnigenic model ^17^, which predicts that all genes that are expressed in the relevant tissue or cell type will have a non-zero effect on disease outcome. The omnigenic model also postulates the existence of core genes that are pivotal in the development of a disease or trait. These core genes are expected to be enriched for genes that are involved in rare Mendelian diseases. The fact that key genes tend to be highly expressed in the relevant tissues for a trait, together with the enrichments of rare disease genes among the key genes, fits the regulatory pattern hypothesized in the omnigenic model. Hence, (some proportion of) the key genes we predict using Downstreamer could be the core genes described in the omnigenic model.

There is an important implication of the enrichment of key genes among known Mendelian disease genes for rare disease diagnostics. On average, a genetic cause is currently identified for only 30% of the patients with a suspected rare disease ^69^. One of the reasons for this low diagnostic yield is that if a rare variant is found in a gene with an unknown function, it is difficult to determine if this variant could be causative for a patient’s phenotype. We expect that in the future approaches like ours could be used to leverage the key genes of common diseases and traits to prioritize candidate rare disease genes in a manner similar to what we did previously using GADO ^24^.

In summary, we present Downstreamer, a method that integrates multi-tissue gene regulatory networks with GWAS summary statistics to prioritize key genes central in the gene network. These key genes were enriched for Mendelian variants that cause related phenotypes, highlighting that GWAS signals partially converge on Mendelian disease genes. While gaps remain in our understanding of the *trans* regulatory architecture of GWAS traits and diseases, assessing the genes most central in their respective regulatory network presents a promising way forward for interpreting both complex and rare disease genetics.

## Methods

### GWAS summary statistics

We downloaded the publicly available summary statistics from either the GWAS catalogue ^70^ or supplementary data files. A full list of the summary statistics used is available as Table S1. Downstreamer requires rs identifiers (RsId) of the variants as well as the p-values. These were extracted from the summary statistic files, and any duplicate variants or variants without a RsId were removed. Where needed, summary statistics were lifted to build 37 and the RsIds matched on position and allele to 1000 Genomes phase 3 EUR for all variants with a minor allele frequency (MAF) > 0.05 ^71^.

### Pathway databases

We used the following pathway and gene-set databases: Reactome^72^, KEGG^73^ and GO^74^ (downloaded July 18, 2020), HPO^75^ (filtered version as in ^76^) and MGI (downloaded October 20, 2020) ^77^.

For the pathway enrichments below in step 2.2, we first expanded the known pathway annotations using the pathway predictions algorithm described in ^76^. We expanded the pathway annotations with all genes with a Bonferroni-significant prediction of a pathway. Using the DEPICT algorithm, we have already shown that using predicted pathway annotations improves pathway enrichments ^8^. Therefore, we used these expanded pathways when associating pathways to traits using Downstreamer.

### Overview of Downstreamer methodology

In short, Downstreamer associates a gene-level prioritization score (GWAS gene z-scores) to a gene–gene co-regulation matrix to find genes that have many connections (at the expression level) to genes inside GWAS loci (core genes). In addition, Downstreamer can identify pathway enrichments by switching the co-regulation matrix for pathway annotations. Downstreamer implements a strategy that can perform these associations while accounting for LD structure and chromosomal organization. Downstreamer operates in two steps. In the first step, the GWAS gene z-scores are calculated for the GWAS trait and a null distribution. In the second step, the GWAS gene z-scores are associated with either the co-regulation matrix or the pathway annotations. Details of these steps are outlined in the sections below.

#### Downstreamer step 1.1. Calculation of GWAS gene z-scores

The first step in Downstreamer is to convert GWAS summary statistics from p-values per variant to an aggregate p-value per gene while accounting for local LD structure (1000 Genomes phase 3 EUR). This p-value is then converted to a gene z-score. This aggregate gene-level z-score represents the GWAS signal potentially attributable to that gene.

This was done as follows. First, we applied genomic control to correct for inflation in the GWAS signal. We then integrated the procedure from the PASCAL method into Downstreamer so that we can aggregate variant p-values into a gene p-value while accounting for the LD structure ^9^. We aggregated all variants within a 25kb window around the start and end of a gene using the non-Finnish European samples of the 1000 Genomes (1000G) project, Phase 3 to calculate LD ^71^. We calculated GWAS gene p-values for all 20,327 protein-coding genes (Ensembl release v75).

#### Downstreamer step 1.2. Null GWAS to account for chromosomal organization of genes and empirical p-value calculations

To account for the longer-range effects of haplotype structure, which result in genes having a similar GWAS gene z-score, we use a GLS regression model for all regressions done in Downstreamer. The GLS model takes a correlation matrix that models this gene– gene correlation.

To calculate this correlation matrix, we first simulated 10,000 random phenotypes by drawing phenotypes from a normal distribution and then associating them to the genotypes of the 1000G Phase 3 non-Finnish European samples. Here, we only used the overlapping variants between the real traits and the permuted GWASs to avoid biases introduced by genotyping platforms or imputation. We then calculated the GWAS gene z-scores for each of the 10,000 simulated GWAS signals, as described above. Next, we calculated the Pearson correlations between the GWAS gene z-scores. As simulated GWAS signals are random and independent of each other, any remaining correlation between GWAS gene z-scores reflects the underlying LD patterns and chromosomal organization of genes.

We simulated an additional 10,000 GWASs as described above to empirically determine enrichment p-values. Finally, we used an additional 100 simulations to estimate the false discovery rate (FDR) of Downstreamer associations.

#### Downstreamer step 1.3. Correction for additional variables and mean gene p-value calculation

To facilitate the correction of additional parameters, variables can be provided which are used to correct the GWAS gene p-values before fitting the GLS (step 2.2). These are fit using a (multivariate) OLS model of which the residuals are taken and used as input for the subsequent steps. We used this option to additionally correct for gene length as well as the mean gene p-value over the 44 traits. The mean gene p-value was calculated by first calculating the mean of the traits in each of the 10 classes of GWAS traits for each gene. Then, for each gene, the mean over these 10 means was calculated in order to avoid having the overrepresented classes (blood cell composition) overshadow the calculation of the means.

#### Downstreamer step 2.1. Pre-processing GWAS gene z-scores and pruning highly correlated genes

For each GWAS, both real and simulated, we force-normalized the GWAS z-scores into a normal distribution to ensure that outliers will not have disproportionate weights. Due to limitations in the PASCAL methodology that result in ties at a minimum significance level of 1×10^-12^ for highly significant genes, we use the minimum SNP p-value from the GWAS to identify the most significant gene and resolve the tie. We then use the linear model (step 1.3) to correct for gene length, as longer genes will typically harbour more SNPs.

Sometimes, two (or more) genes will be so close to one another that their GWAS gene z-scores are highly correlated, violating the assumptions of the linear model. Thus, genes with a Pearson correlation r ≥ 0.8 in the 10,000 GWAS permutations were collapsed into ‘meta-genes’ and treated as one gene. Meta-gene z-scores were averaged across the input z-scores. Lastly, the GWAS z-scores of the meta genes were scaled (mean = 0, standard deviation = 1).

#### Downstreamer step 2.2. GLS to calculate key gene scores and pathway enrichments

We used a GLS regression to associate the GWAS gene z-scores with the gene co-regulation z-scores or with the expanded pathway annotations. These two analyses result in the key gene prioritizations and pathway enrichments, respectively. We used the gene–gene correlation matrix derived from the 10,000 permutations as a measure of the conditional covariance of the error term (Ω) in the GLS to account for the relationships between genes due to LD and proximity. The pseudo-inverse of Ω is used as a substitute for Ω^-1^

The formula of the GLS is as follows:

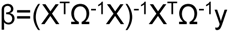

Where β is the estimated effect size of the pathway, term or gene from the co-regulation matrix; Ω is the gene-gene correlation matrix; X is the design matrix of real GWAS z-scores and y is the vector of gene z-scores per pathway, term or gene from the co-regulation matrix. As we standardized the predictors, we did not include an intercept in the design matrix and X only contains one column with the real GWAS gene z-scores. We estimated the betas for the 10,000 random GWASs in the same way and subsequently used them to estimate the empirical p-value for β.

#### Downstreamer step 2.3. Pathway and gene set gene enrichments

To identify pathway and disease enrichments, we used the following databases: HPO, KEGG, Reactome, MGI and GO Biological Process, Cellular Component and Molecular Function. We have previously predicted how much each gene contributes to these gene sets, resulting in a z-score per pathway or term per gene ^24^. We then used Bonferroni correction to determine if the gene should be added to the extended pathway membership.

Next, we collapsed genes into meta-genes, in parallel with the GWAS step, to ensure compatibility with the GWAS gene z-scores, following the same procedure as in the GWAS pre-processing. The pathway memberships for a meta-gene were calculated as the sum of the membership divided by the square root of the number of genes. So, a meta-gene containing five genes, of which two are in a pathway, would get a value of 2 / √5 = 0.89 for that pathway. Finally, the pathway memberships of the meta-genes were scaled and centred (mean = 0, standard deviation = 1).

#### Downstreamer step 2.4: Co-regulation matrix

To calculate key gene scores, we used a previously generated co-regulation matrix based on a large multi-tissue gene network ^24^. In short, publicly available RNA-seq samples were downloaded from the European Nucleotide Archive (https://www.ebi.ac.uk/ena). After quality control, 56,435 genes and 31,499 samples covering a wide range of human cell-types and tissues remained. We performed a principal component analysis on this dataset and selected the 165 principal components representing 50% of the variation that offered the best prediction of gene function ^76^. We then selected the protein-coding genes and centred and scaled the eigenvectors for these 165 components (mean = 0, standard deviation = 1) such that each component was given equal weight. The first components mostly describe tissue differences ^24^, so this normalization ensures that tissue-specific patterns do not disproportionately drive the co-regulation matrix. The co-regulation matrix is defined as the Pearson correlation between the genes from the scaled eigenvector matrix. The diagonal of the co-regulation matrix was set to zero to avoid correlation with itself having a disproportionate effect on the association to the GWAS gene z-scores. Finally, we converted the Pearson r to z-scores. To associate the co-regulation to the gene z-scores, the same meta-gene procedure was applied as outlined for the pathway enrichments.

### Enrichment of key genes

Enrichments of key genes among HPO/MGI/GO terms and KEGG gene sets was done by Fisher’s exact test, taking all key genes at Bonferroni or FDR significance and comparing their overlap to all other genes. AUCs were calculated by dividing the Mann-Whitney U statistic of the key gene z-scores and gene set membership by the product of sample sizes. The gene-pathway/term definitions we used were those provided by the respective databases, thus they were not the extended versions used for the GWAS gene set enrichments. This is implemented in Downstreamer using –T PRIO_GENE_ENRICH.

### Enrichment of average gene z-scores and association with LD and gene density

Enrichments of the top 500 average gene z-scores were done by first correcting the mean gene z-score vector (see step 1.3 for details on calculating this) for the extent of the LD around a gene as well as the gene density. To quantify the extent of the LD block, we took the mean of the LD scores of all SNPs in a 25kb window around the gene. Pre-computed European LD scores were downloaded from https://github.com/bulik/ldsc. Gene density was calculated by counting the number of genes in a 500kb window around the start end of the gene. Both these factors were then fit in a linear model with the mean gene z-score as the outcome. The residuals were taken and ranked to arrive at the top 500 genes. We then carried out overrepresentation analysis using https://toppgene.cchmc.org/enrichment.jsp with the default background set.

### Association with LoF and MiS intolerance

MiS and LoF intolerance z-scores were downloaded from the gnomAD consortium (https://gnomad.broadinstitute.org/downloads > pLoF Metrics by Gene TSV v2.1.1). As an overall measure of the “keyness” of a gene, we calculated the maximum key gene z-score observed over the 44 traits for each gene. We then associated this to the MiS and LoF z-scores from the gnomAD consortium by Pearson correlation.

### Enrichment of cis-eQTL and key genes

Enrichments of cis-eQTL and key genes were calculated by fisher exact test, taking all the genes tested in eQTLgen or MetaBrian respectively as the background set. A gene was considered to be a *cis*-eQTL gene if it had a significant association in eQTLgen or MetaBrain analyses respectively.

### Overlap with trans-eQTLs and eQTS genes

To investigate their overlap with the key genes identified by Downstreamer, we downloaded the *trans*-eQTL and eQTS results from the eQTLGen Consortium (www.eqtlgen.org). For each GWAS, we selected all *trans*-eQTLs that emanate from independent top SNPs (1000 Genomes phase 3 EUR, r^2^ 0.2, 500kb window) and calculated the sum of *trans*-eQTL squared z-scores for each gene. We then log-transformed this and associated it to the key gene z-score for the GWAS using Pearson correlation.

For the overlap with eQTS genes, we selected eQTSs for which we had overlapping GWAS traits. We then evaluated if the eQTS genes had a higher key gene z-score compared to all other genes using a Student’s T-test.

## Data Availability

Software and scripts are available for download at: https://github.com/molgenis/systemsgenetics/tree/master/Downstreamer
A manual for Downstreamer is available at: https://github.com/molgenis/systemsgenetics/wiki/Downstreamer
All RNA-seq data used in the main analysis are publicly available in the European Nucleotide Archive.

## Code and data availability

Software and scripts are available for download at: https://github.com/molgenis/systemsgenetics/tree/master/Downstreamer

A manual for Downstreamer is available at: https://github.com/molgenis/systemsgenetics/wiki/Downstreamer

All RNA-seq data used in the main analysis are publicly available in the European Nucleotide Archive, for details please see ^24^.

## Acknowledgements

We thank Kate McIntyre for the editorial assistance.

We thank the UMCG Genomics Coordination Center, the UG Center for Information Technology and their sponsors BBMRI-NL & TarGet for storage and compute infrastructure.

## Author contributions

• Conceptualization: O.B.B., A.C., L.F., P.D.

• Data curation: O.B.B., A.C., P.D.

• Formal Analysis: O.B.B., A.C., P.D.

• Funding acquisition: L.F., P.D.

• Investigation: A.C., O.B.B., H.J.W., H.W., F.B., U.V., S.M.S.

• Methodology: O.B.B., L.F., P.D.

• Software: O.B.B., H.W., P.D.

• Supervision: O.B.B., I.H.J., L.F.

• Visualization: O.B.B., A.C., L.F., P.D.

• Writing – original draft: O.B.B., A.C., L.F., P.D.

• Writing – review & editing: I.H.J, H.J.W., U.V.,

Roles as defined by: CRediT (Contributor Roles Taxonomy)

## Materials & Correspondence

Lude Franke, Patrick Deelen;

## Supplementary data

**Fig. S1.**
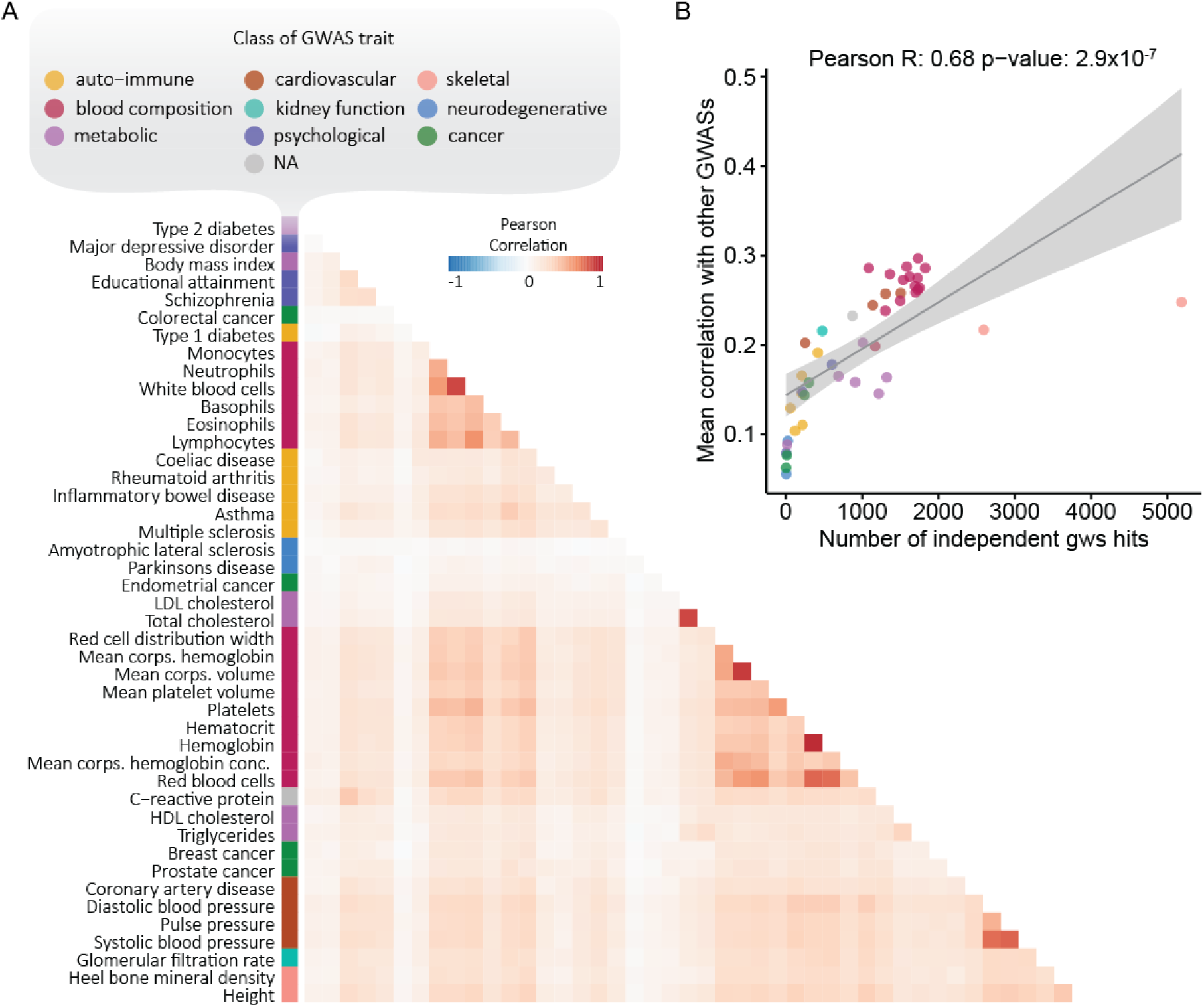
Association between gene z-score profiles of different traits. A) Pearson correlations between gene z-scores reveal that most pairwise correlations between traits are positive. B) Mean correlation in gene z-scores with all other GWASs (y-axis) versus the number of independent genome-wide significant hits for the respective GWAS.

**Fig. S2.**
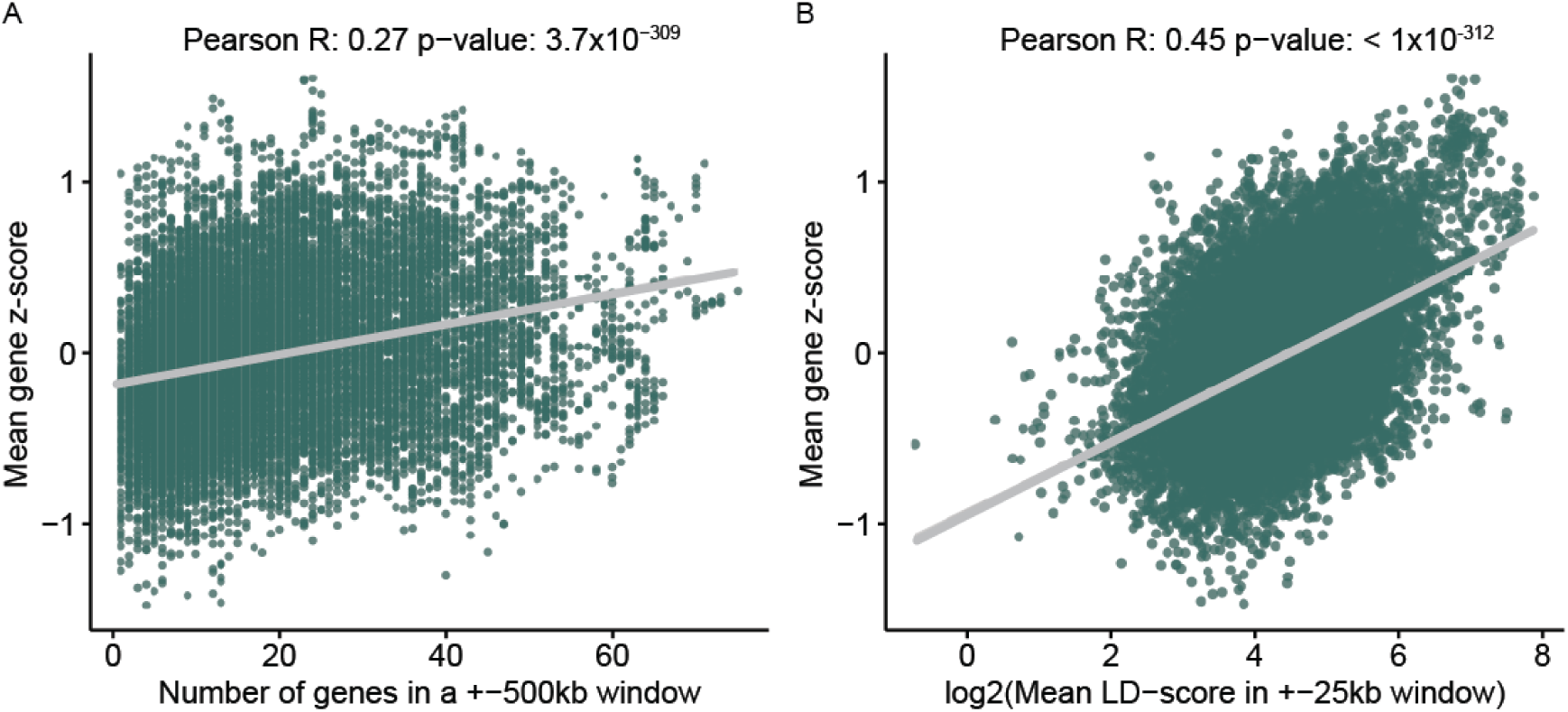
Association between average gene z-score and LD and gene density. A) Association between average gene z-score (y-axis) and the number of genes within a ±500kb window (x-axis). B) As in (A), but x-axis indicates the log2 of the average LD score of SNPs located ±25kb around the start and end of a gene. The adjusted R2 of the model associating the average gene z-score and these two parameters as the independent variables is 0.298 p-value <1e-16.

**Fig. S3.**
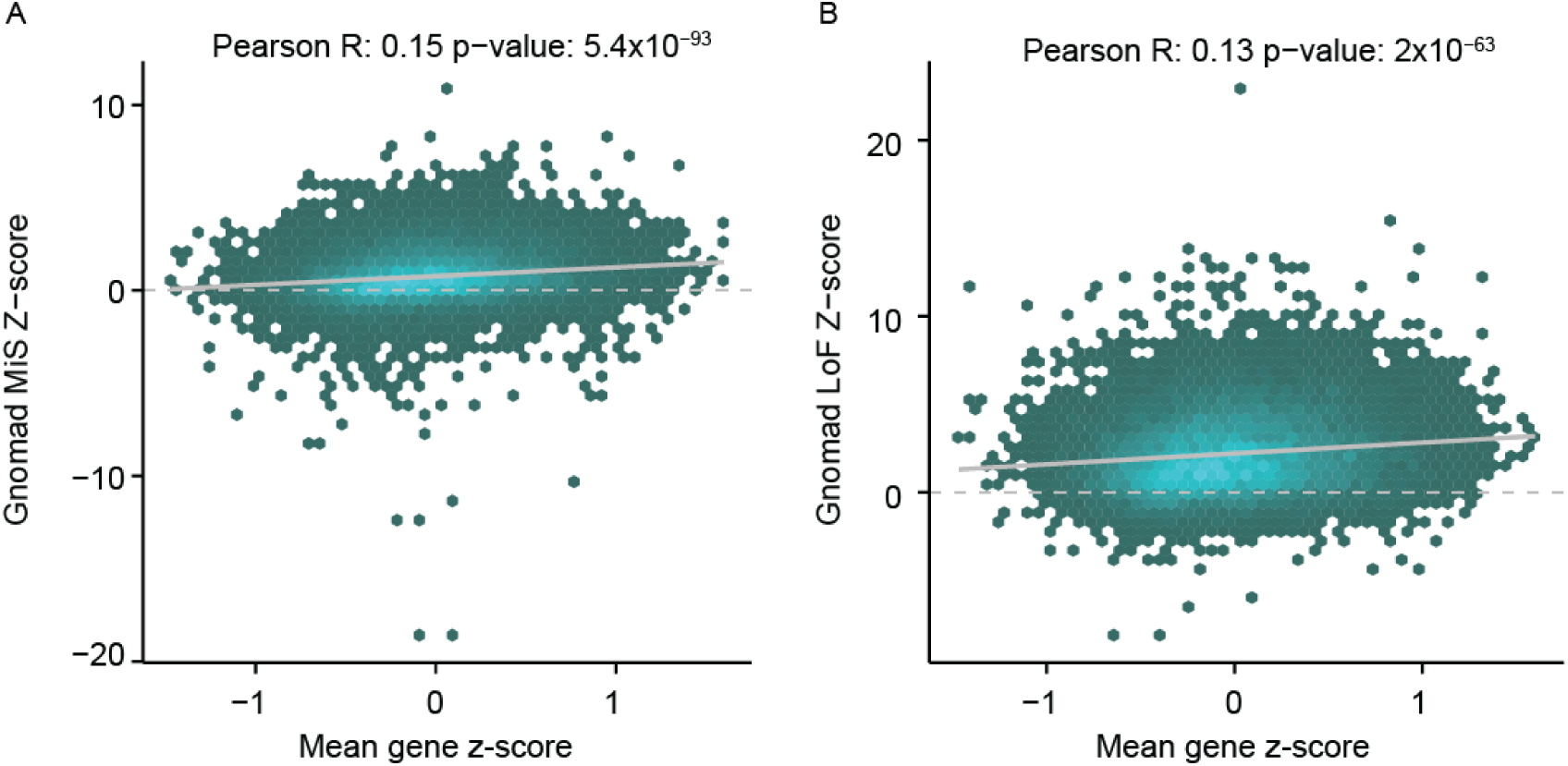
Enrichment of missense and loss of function intolerance in the average gene z-score. A) Association between average gene z-score (x-axis) and the missense intolerance z-scores from the gnomAD consortium (y-axis). B) As in (A), but the y-axis indicated the z-score for loss of function intolerance. Pearson correlation coefficients indicated.

**Fig. S4.**
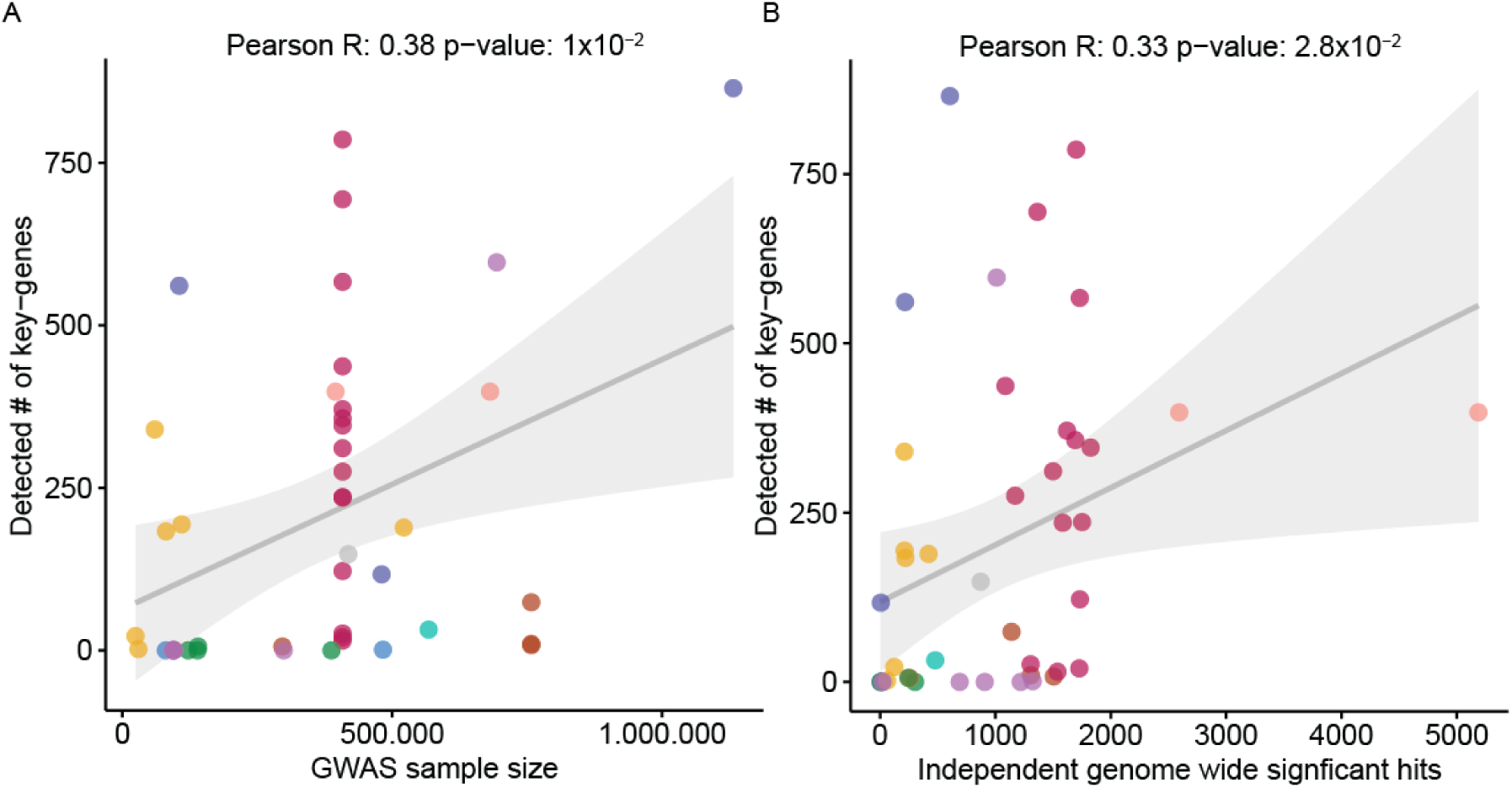
Relationship between number of samples used for a GWAS and the power to detect key genes. A) Association between the detected number of key genes for each GWAS (y-axis) and the sample size of that GWAS (x-axis). B) As in (A), but the x-axis shows the number of independent genome-wide significant hits as determined by clumping using a LD R^2^ threshold of 0.2 in a 250kb window.

**Fig. S5.**
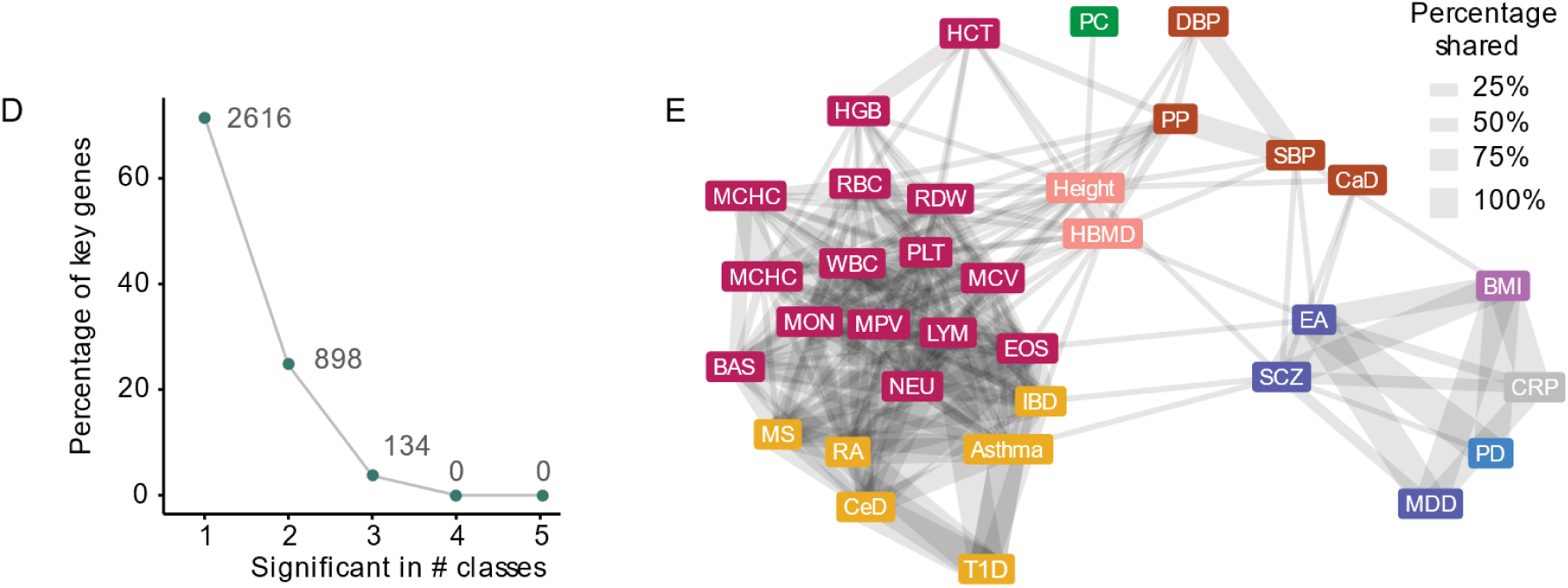
Sharing among the key genes. A) Percentage of all detected key genes (y-axis) versus the number of classes in which the key gene was detected. B) Network plot showing the sharing of key genes (Bonferroni significant). Edges represent the percentage of sharing calculated on the smaller of the pair. Nodes represent a GWAS trait. Traits without predicted key genes or traits that show no overlap are omitted in this panel.

**Fig. S6.**
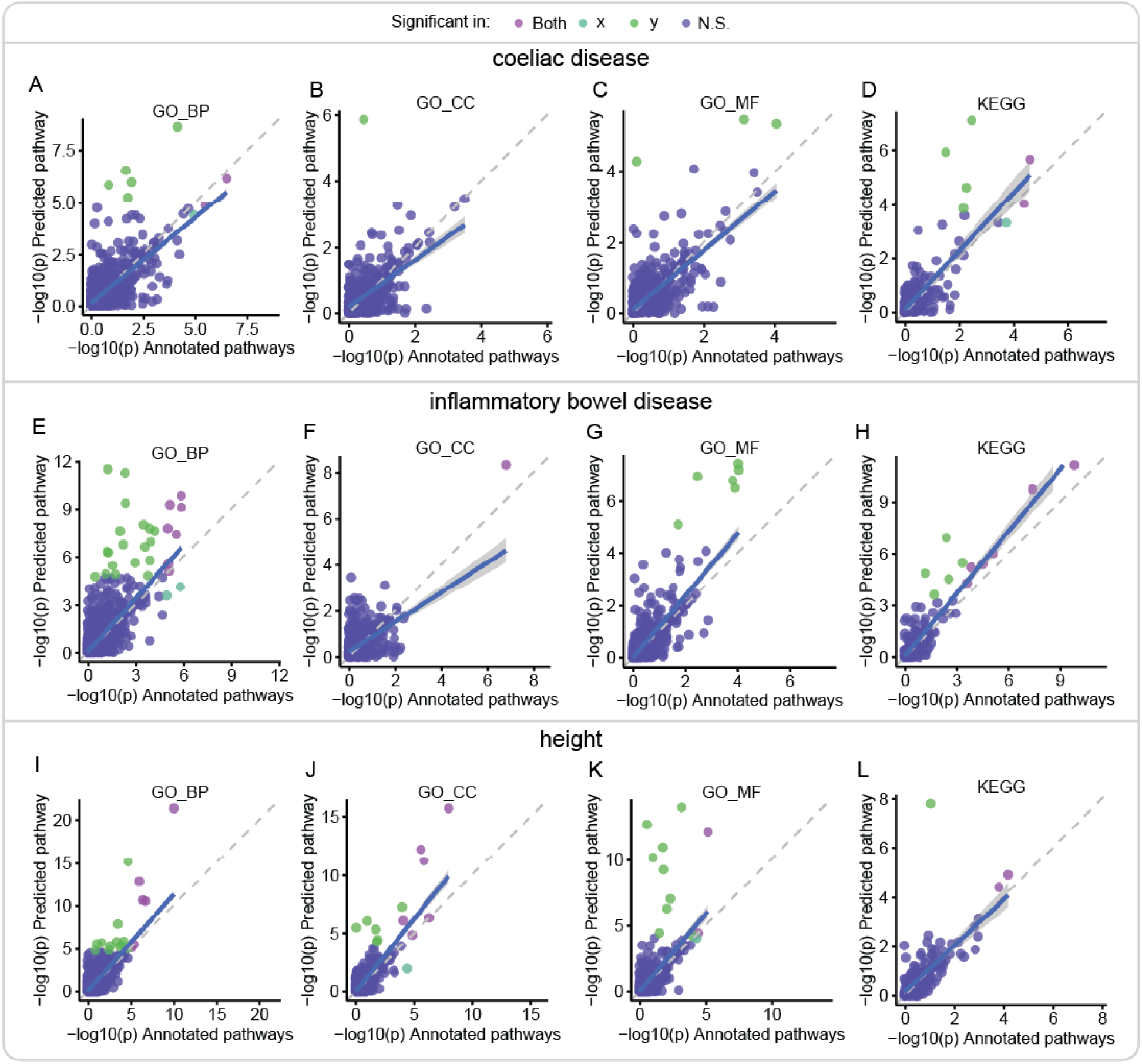
Comparison of pathway enrichment results before and after extending pathway definitions using a gene regulatory network. Comparisons of Downstreamer association -log10(p-values) for the pathway databases GO BP, GO MF, GO CC and KEGG before extending the pathway memberships (x-axis) and after extending the pathway memberships. A–D) indicate the comparison for CeD. E–H) indicate the comparison for inflammatory bowel disease. I–L) indicate the results for height. Colours indicate significance: bright green significant in extended set only, blue green significant in annotated set only, purple significant in both, blue significant in neither.

**Fig. S7. Sample enrichment per plots per traits**

Provided separately

The first plot is an UMAP coloured using the annotations of the 31,499 RNA-seq samples used. The right panel is a zoom-in of the marked region to better reveal the differences of the primary tissues. The plots on the other pages show the enrichment scores for each sample per GWAS. Only samples that pass the FDR 5% threshold are given a colour indicating enrichment or depletion.

**Fig. S8.**
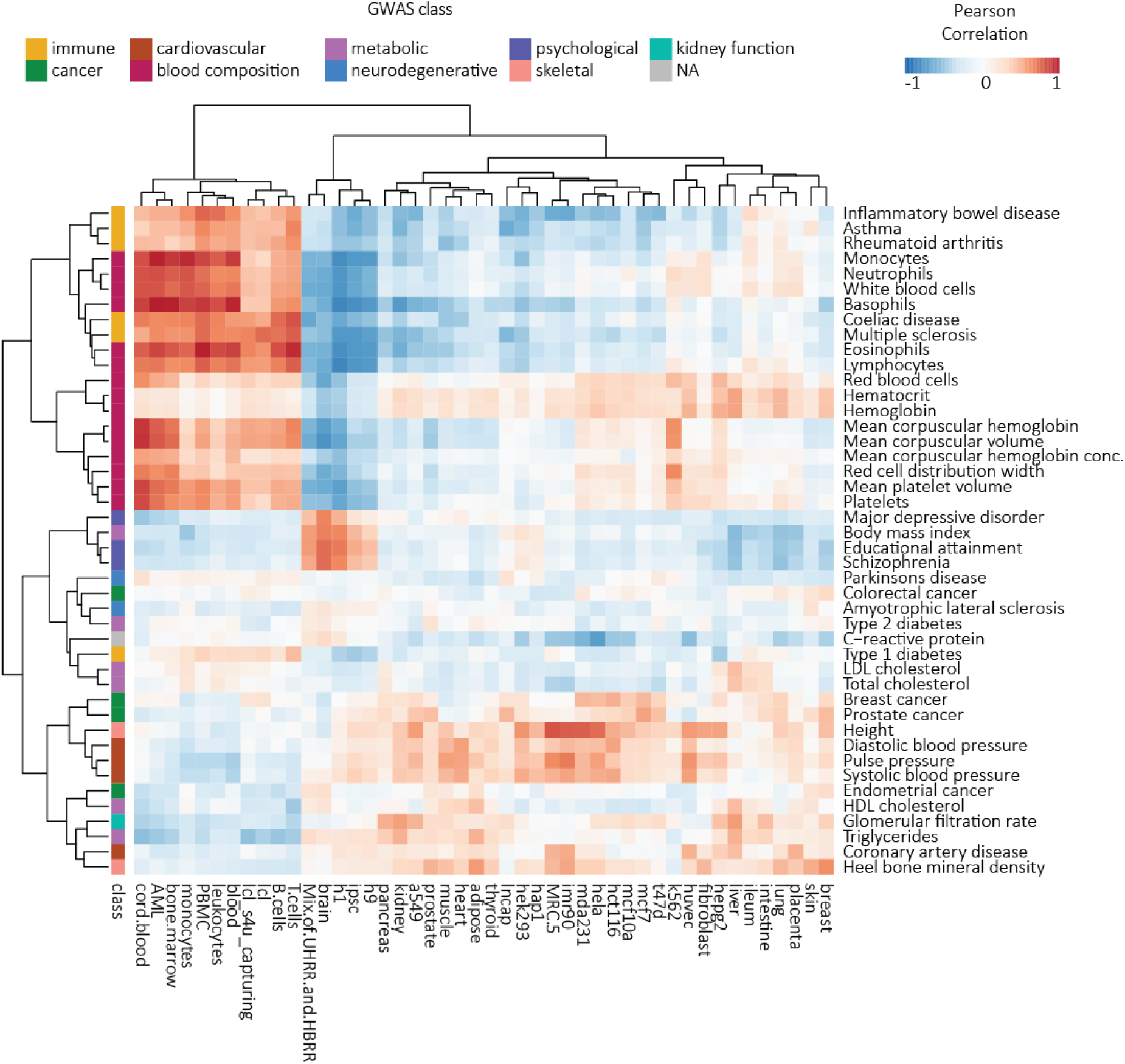
Correlations between tissue expression and key gene z-scores. Correlation heatmap showing Pearson correlation coefficients between the key gene z-scores calculated by Downstreamer and the specificity of expression of a gene. Specificity of expression was determined by taking the mean in the query samples and subtracting the mean from all other annotated samples in the dataset.

**Fig. S9.**
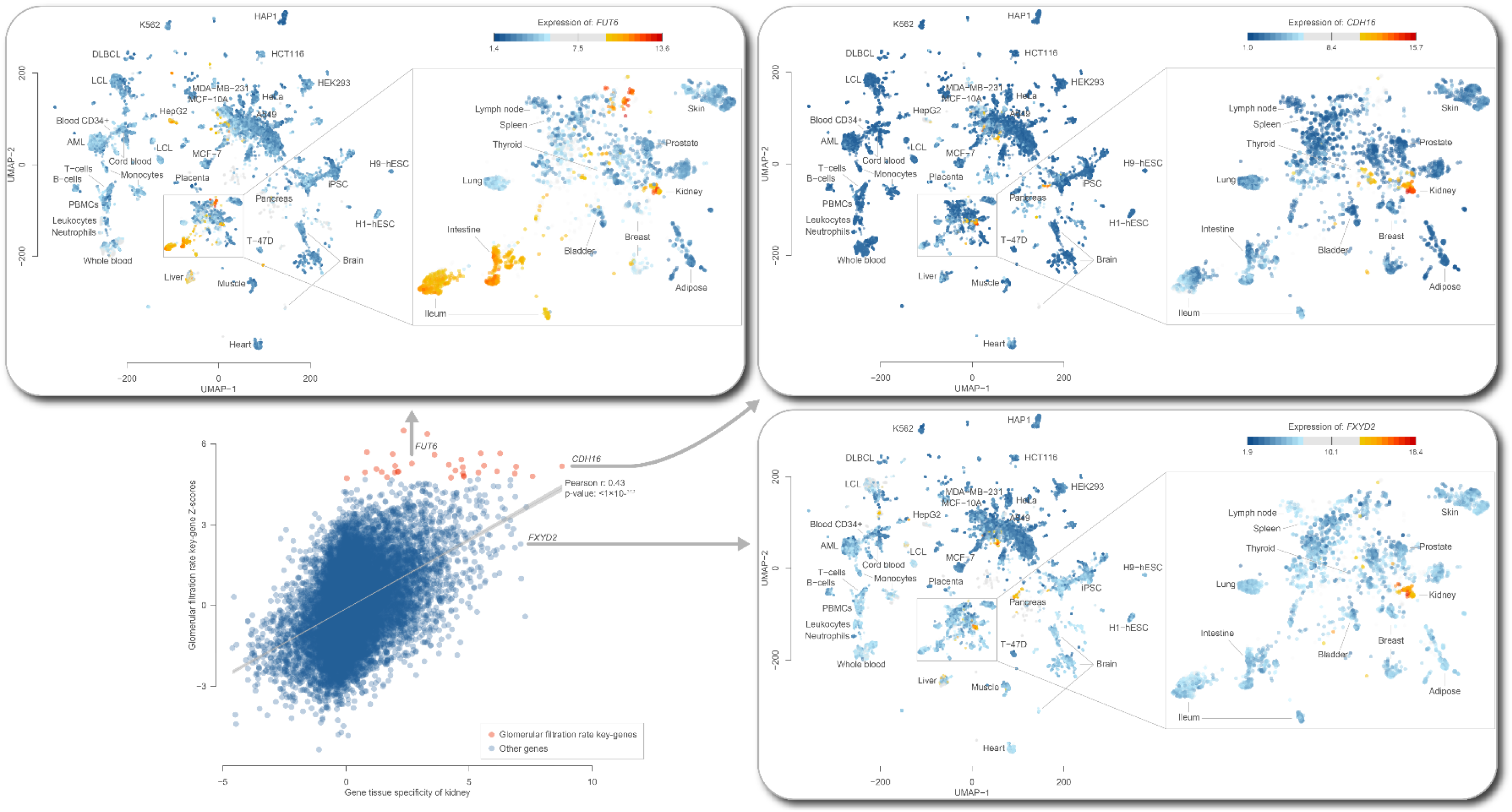
Relation between glomerular filtration rate key gene scores and expression levels. Expression specificity of genes in kidney vs key gene scores of glomerular filtration rate. We highlight three genes to show that not all key genes are kidney-specific genes and that not all kidney-specific genes are predicted to be key genes.

**Fig. S10.**
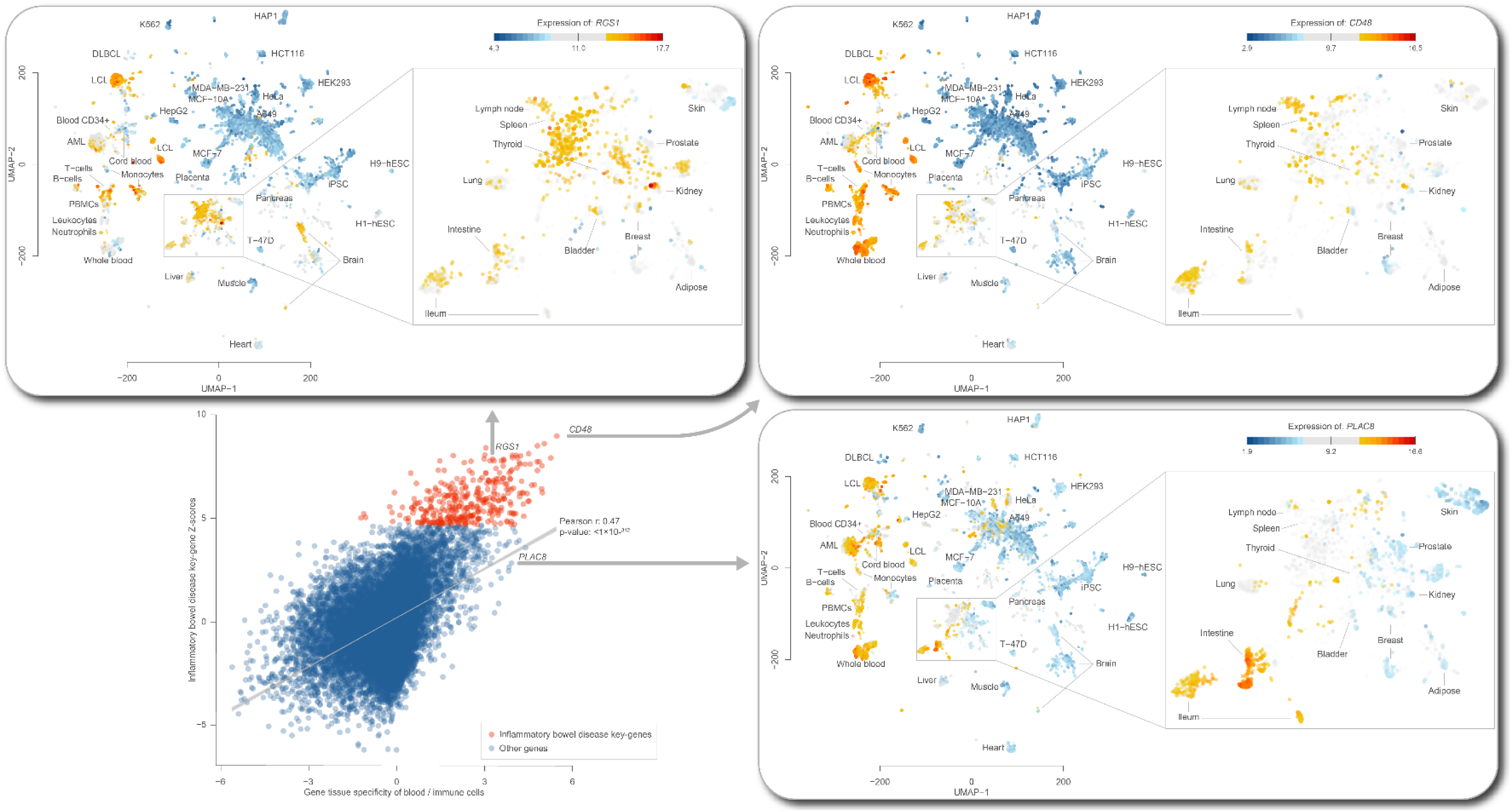
Relation between inflammatory bowel disease key gene scores and expression levels. Expression specificity of genes in blood and immune cells vs key gene scores of inflammatory bowel disease. We highlight three genes to show that not all key genes are blood/immune specific genes and that not all blood and immune genes are predicted to be key genes.

**Fig. S11.**
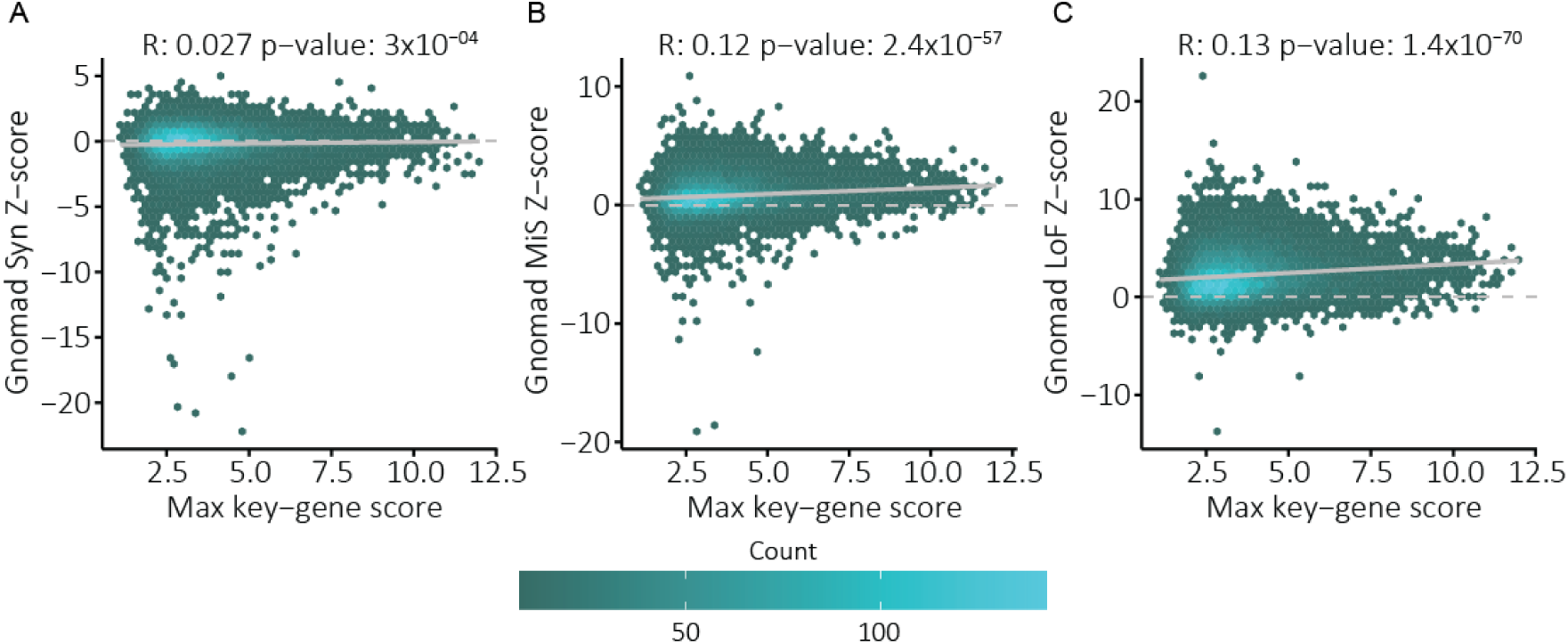
Association between LoF and MiS scores and key gene z-scores. Association between the maximum key gene score for a gene (x-axis) and the constraint metrics from the gnomAD consortium (y-axis). Pearson correlations are indicated above each panel. A) Association with the synonymous variant z-score. B) Association with the missense variant z-score. C) Association with the loss of function variant z-score.

**Fig. S12.**
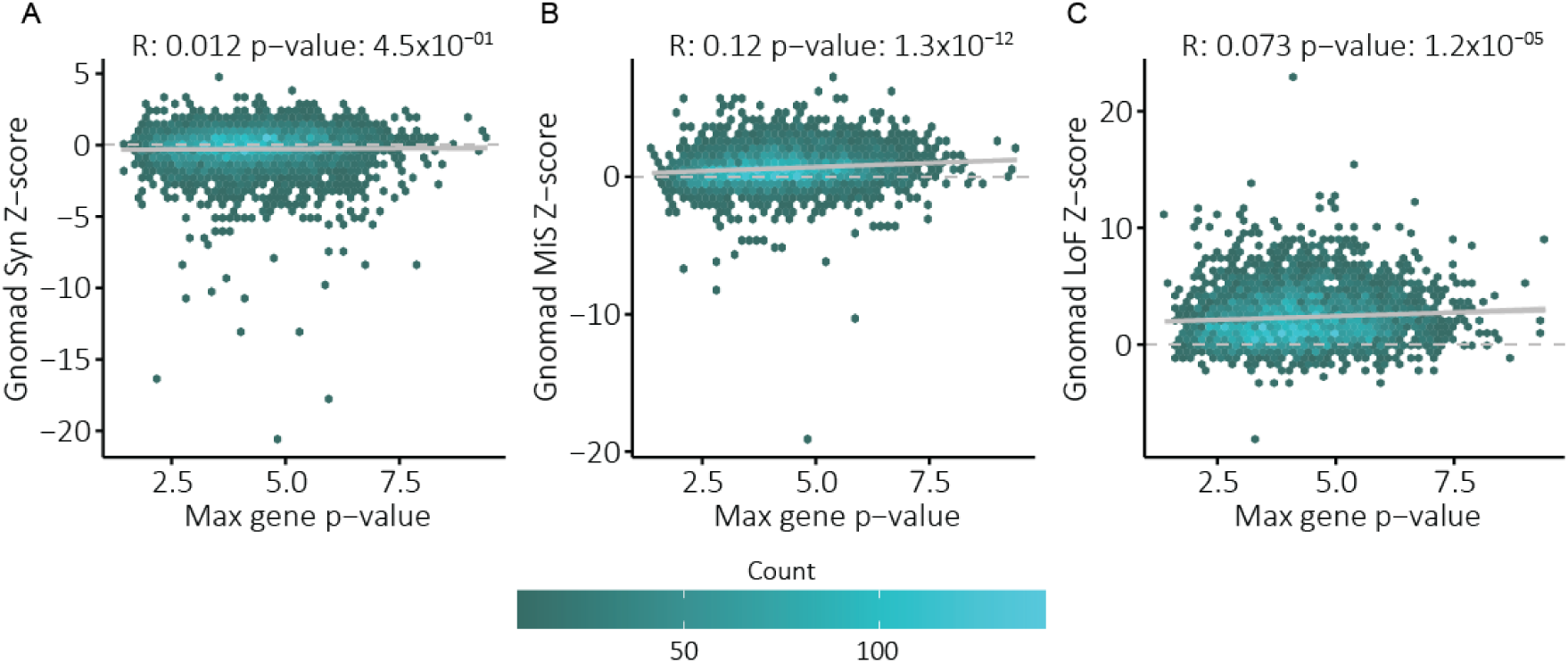
Association between LoF and MiS scores and GWAS gene p-value. Association between the maximum GWAS gene p-value gene calculated by Pascal (x-axis) and the constraint metrics from the gnomAD consortium (y-axis). Pearson correlations are indicated above each panel. A) Association with the synonymous variant z-score. B) Association with the missense variant z-score. C) Association with the loss of function variant z-score.

**Fig. S13.**
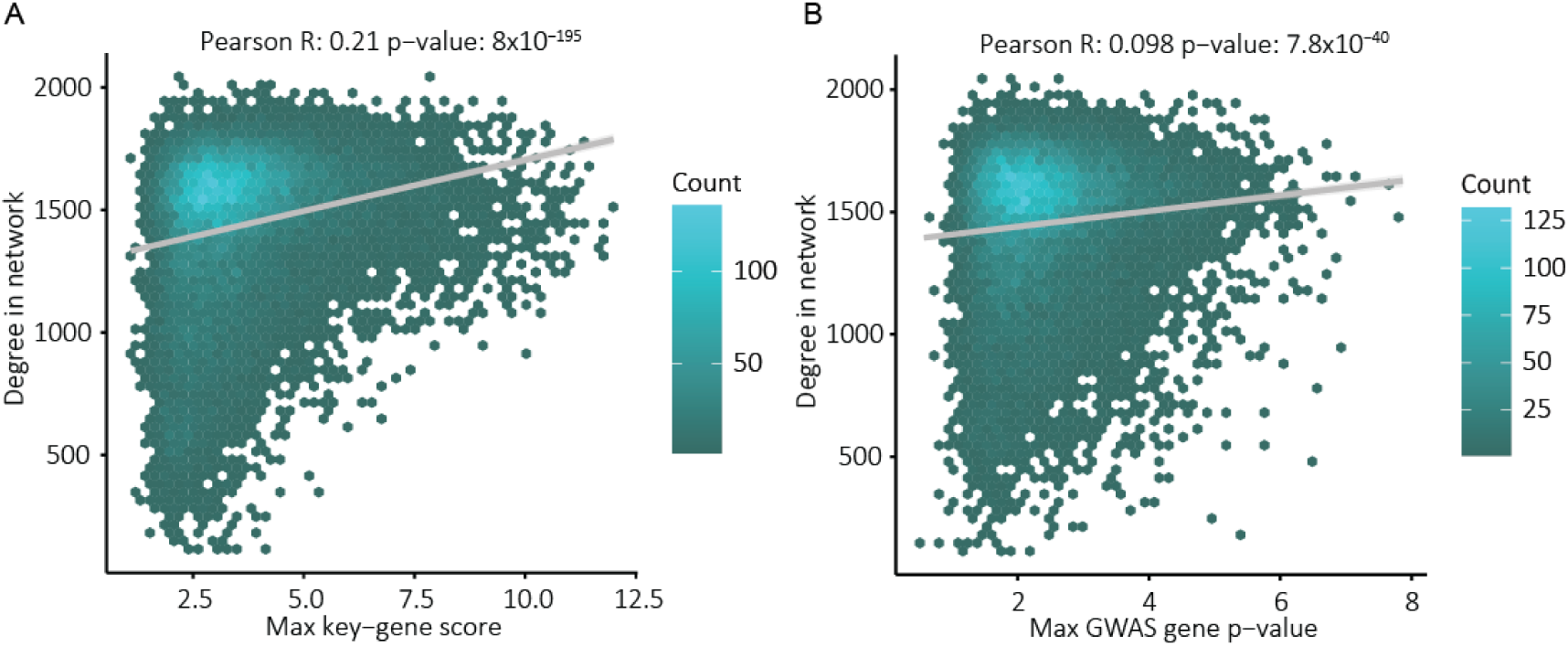
Association between degree and key gene z-scores. A) Association between the degree of a gene in the gene network (y-axis; degree indicates the number of connections at a z-score threshold of 1.6) and the maximum key gene score a gene obtained (x-axis). B) As in (A), but x-axis indicates the maximum GWAS gene p-value calculated by Pascal. Pearson correlations are indicated above each panel.

**Fig. S14.**
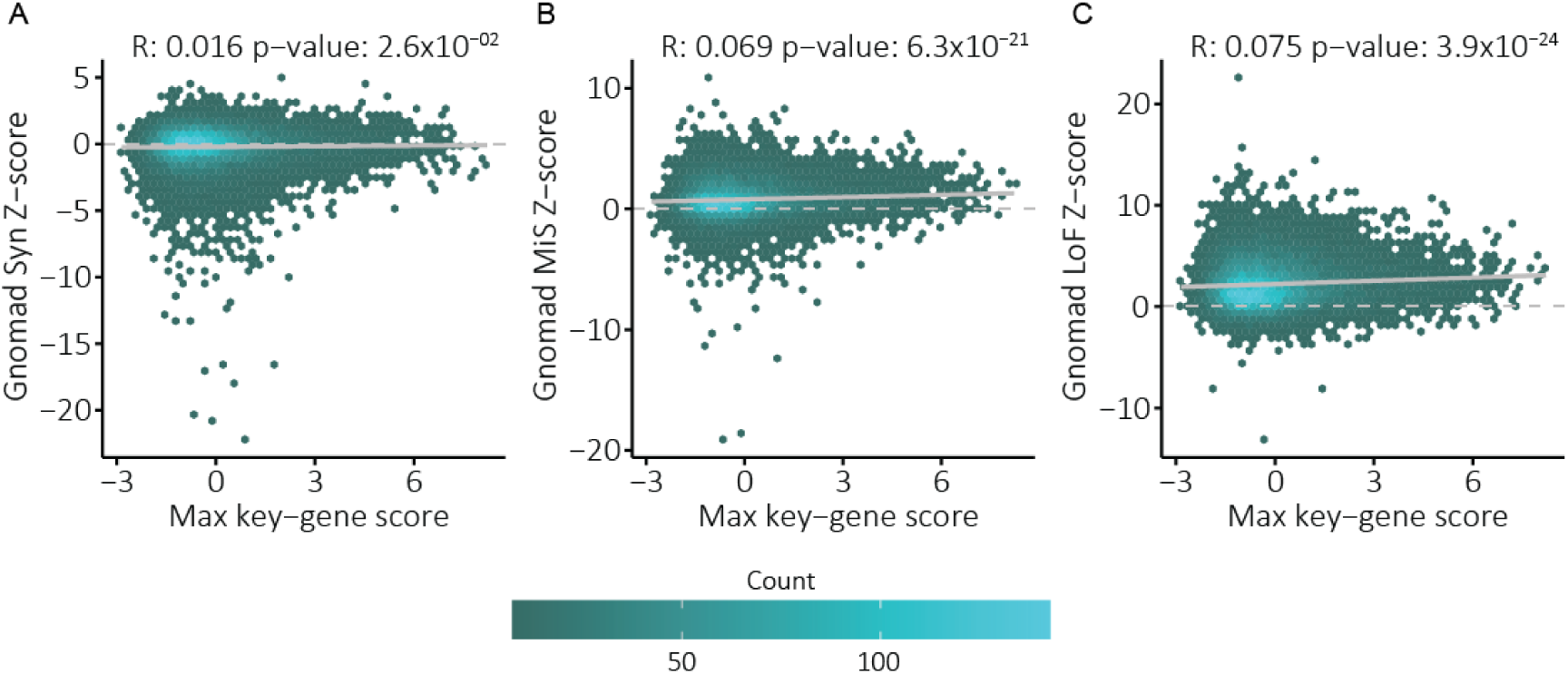
Association between LoF and MiS scores and key gene z-scores after correction for the degree. Association between the maximum key gene score a gene obtained after linearly correcting the key gene scores for the degree in the gene network (x-axis; degree indicates the number of connections at a z-score threshold of 1.6) and the constraint metrics from the gnomAD consortium (y-axis). Pearson correlations are indicated above each panel. A) Association with the synonymous variant z-score. B) Association with the missense variant z-score. C) Association with the loss of function variant z-score.

**Fig. S15.**
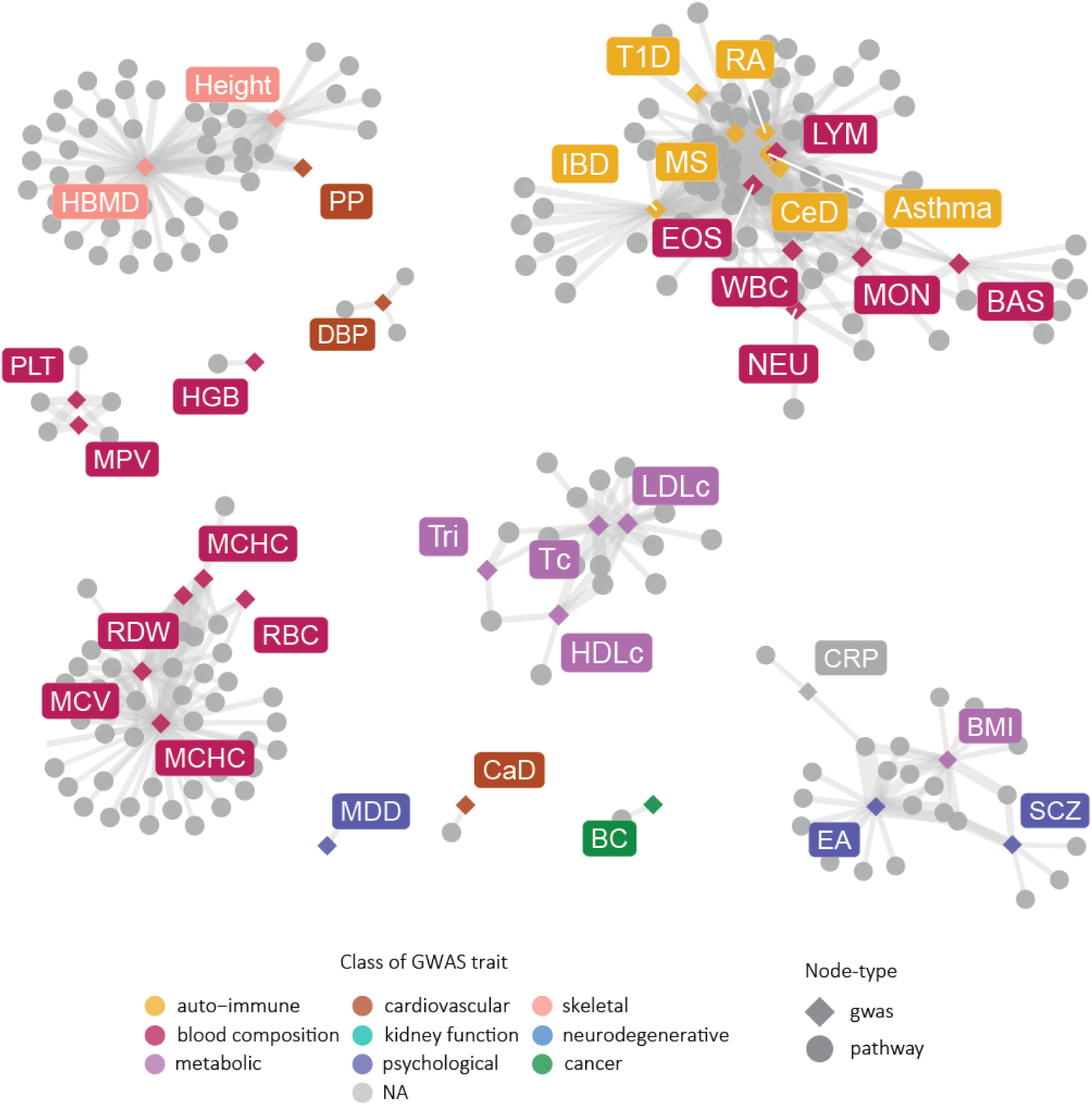
Sharing between GWASs among pathway enrichments for GO biological process. Network plot showing Bonferroni-significant pathways (grey dots). Connections with the respective GWAS are indicated by an edge. Colours indicate the class of GWAS trait. Clusters formed with white blood cells and immune diseases, hematopoietic factors, lipid levels and psychological traits

**Table S1.**
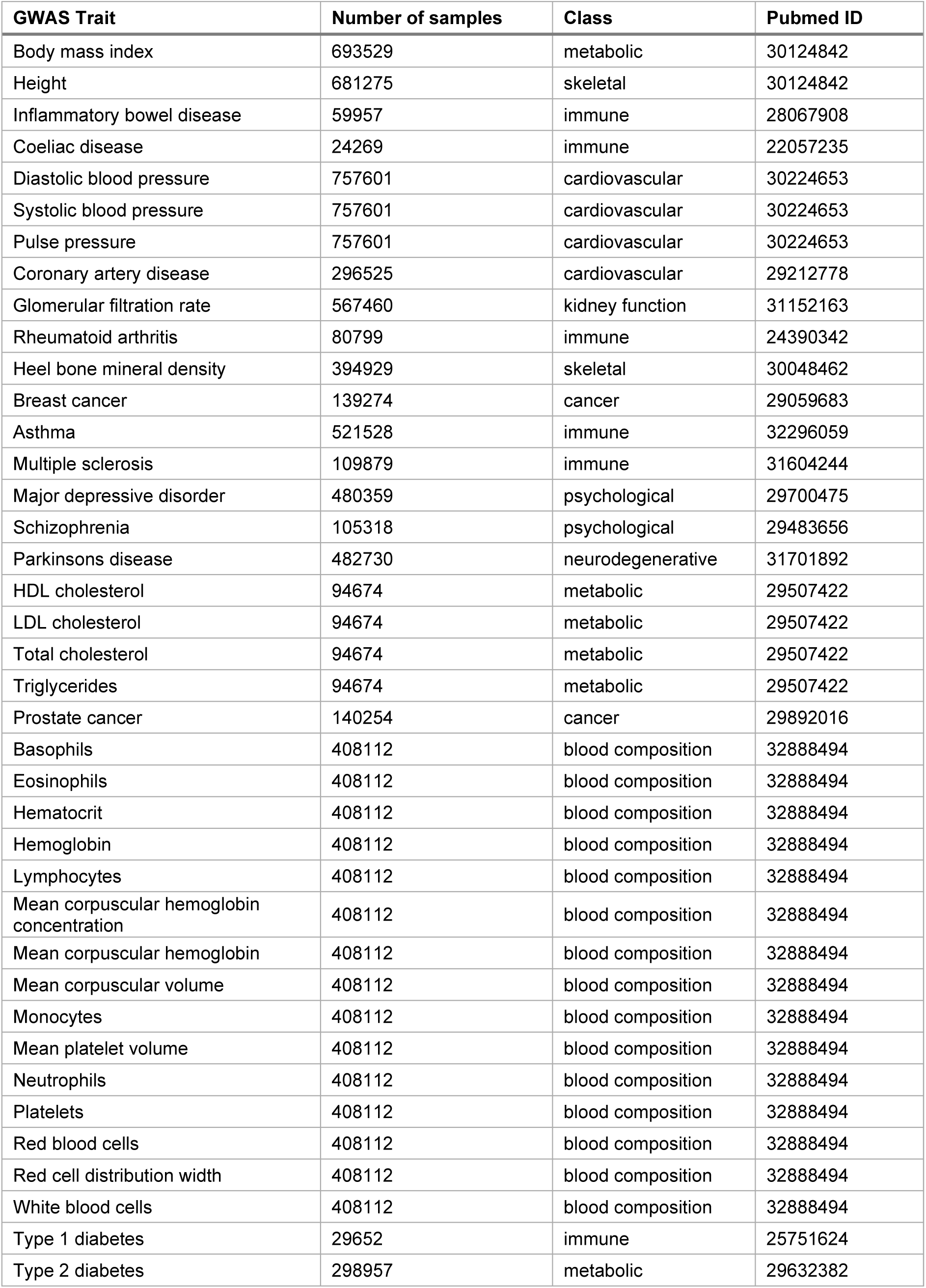
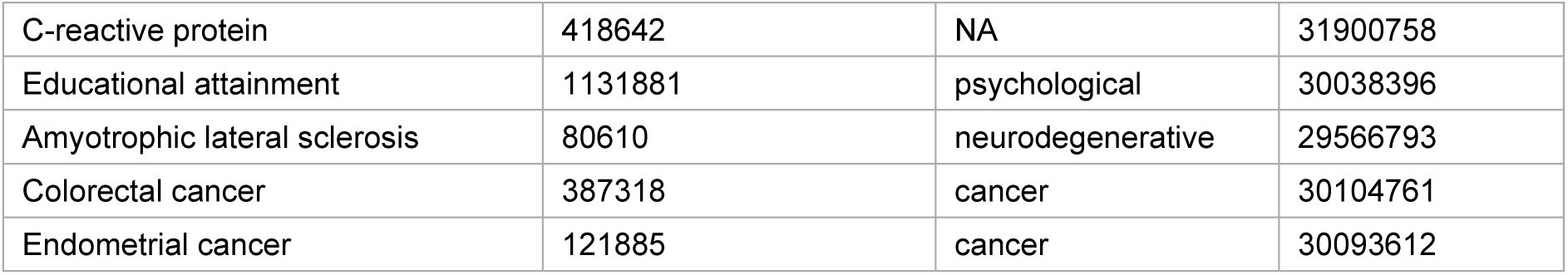
List of the 44 complex traits and diseases to which we applied Downstreamer.

| GWAS Trait | Number of samples | Class | Pubmed ID |
| --- | --- | --- | --- |
| Body mass index | 693529 | metabolic | 30124842 |
| Height | 681275 | skeletal | 30124842 |
| Inflammatory bowel disease | 59957 | immune | 28067908 |
| Coeliac disease | 24269 | immune | 22057235 |
| Diastolic blood pressure | 757601 | cardiovascular | 30224653 |
| Systolic blood pressure | 757601 | cardiovascular | 30224653 |
| Pulse pressure | 757601 | cardiovascular | 30224653 |
| Coronary artery disease | 296525 | cardiovascular | 29212778 |
| Glomerular filtration rate | 567460 | kidney function | 31152163 |
| Rheumatoid arthritis | 80799 | immune | 24390342 |
| Heel bone mineral density | 394929 | skeletal | 30048462 |
| Breast cancer | 139274 | cancer | 29059683 |
| Asthma | 521528 | immune | 32296059 |
| Multiple sclerosis | 109879 | immune | 31604244 |
| Major depressive disorder | 480359 | psychological | 29700475 |
| Schizophrenia | 105318 | psychological | 29483656 |
| Parkinsons disease | 482730 | neurodegenerative | 31701892 |
| HDL cholesterol | 94674 | metabolic | 29507422 |
| LDL cholesterol | 94674 | metabolic | 29507422 |
| Total cholesterol | 94674 | metabolic | 29507422 |
| Triglycerides | 94674 | metabolic | 29507422 |
| Prostate cancer | 140254 | cancer | 29892016 |
| Basophils | 408112 | blood composition | 32888494 |
| Eosinophils | 408112 | blood composition | 32888494 |
| Hematocrit | 408112 | blood composition | 32888494 |
| Hemoglobin | 408112 | blood composition | 32888494 |
| Lymphocytes | 408112 | blood composition | 32888494 |
| Mean corpuscular hemoglobin concentration | 408112 | blood composition | 32888494 |
| Mean corpuscular hemoglobin | 408112 | blood composition | 32888494 |
| Mean corpuscular volume | 408112 | blood composition | 32888494 |
| Monocytes | 408112 | blood composition | 32888494 |
| Mean platelet volume | 408112 | blood composition | 32888494 |
| Neutrophils | 408112 | blood composition | 32888494 |
| Platelets | 408112 | blood composition | 32888494 |
| Red blood cells | 408112 | blood composition | 32888494 |
| Red cell distribution width | 408112 | blood composition | 32888494 |
| White blood cells | 408112 | blood composition | 32888494 |
| Type 1 diabetes | 29652 | immune | 25751624 |
| Type 2 diabetes | 298957 | metabolic | 29632382 |
| C-reactive protein | 418642 | NA | 31900758 |
| Educational attainment | 1131881 | psychological | 30038396 |
| Amyotrophic lateral sclerosis | 80610 | neurodegenerative | 29566793 |
| Colorectal cancer | 387318 | cancer | 30104761 |
| Endometrial cancer | 121885 | cancer | 30093612 |

**Table S2. Enrichment of top 500 genes from average GWAS signal**

Provided separately

**Table S3.** Mapping of GWAS traits to HPO-terms.

| GWAS Trait | Related HPO term | Related HPO ID |
| --- | --- | --- |
| Body mass index | Abnormality of body mass index | HP:0045081 |
| Height | Abnormality of body height | HP:0000002 |
| Inflammatory bowel disease | Increased inflammatory response | HP:0012649 |
| Coeliac disease | Increased inflammatory response | HP:0012649 |
| Diastolic blood pressure | Abnormal systemic blood pressure | HP:0030972 |
| Systolic blood pressure | Abnormal systemic blood pressure | HP:0030972 |
| Pulse pressure | Abnormal systemic blood pressure | HP:0030972 |
| Coronary artery disease | Abnormality of the cardiovascular system | HP:0001626 |
| Glomerular filtration rate | Abnormal renal physiology | HP:0012211 |
| Rheumatoid arthritis | Increased inflammatory response | HP:0012649 |
| Heel bone mineral density | Abnormality of the skeletal system | HP:0000924 |
| Breast cancer | Neoplasm | HP:0002664 |
| Asthma | Increased inflammatory response | HP:0012649 |
| Multiple sclerosis | Increased inflammatory response | HP:0012649 |
| Major depressive disorder | Behavioral abnormality | HP:0000708 |
| Schizophrenia | Behavioral abnormality | HP:0000708 |
| Parkinsons disease | Involuntary movements | HP:0004305 |
| HDL cholesterol | Abnormal HDL cholesterol concentration | HP:0031888 |
| LDL cholesterol | Abnormal LDL cholesterol concentration | HP:0031886 |
| Total cholesterol | Abnormal circulating cholesterol concentration | HP:0003107 |
| Triglycerides | Abnormal circulating cholesterol concentration | HP:0003107 |
| Prostate cancer | Neoplasm | HP:0002664 |
| Basophils | Abnormal basophil count | HP:0031806 |
| Eosinophils | Abnormal eosinophil count | HP:0020064 |
| Hematocrit | Abnormal hematocrit | HP:0031850 |
| Hemoglobin | Anemia | HP:0001903 |
| Lymphocytes | Abnormal lymphocyte count | HP:0040088 |
| Mean corpuscular hemoglobin concentration | Abnormal mean corpuscular hemoglobin concentration | HP:0025546 |
| Mean corpuscular hemoglobin | Abnormal hemoglobin | HP:0011902 |
| Mean corpuscular volume | Abnormal mean corpuscular volume | HP:0025065 |
| Monocytes | Abnormal monocyte count | HP:0012310 |
| Mean platelet volume | Abnormal platelet volume | HP:0011876 |
| Neutrophils | Abnormal neutrophil count | HP:0011991 |
| Platelets | Abnormal platelet count | HP:0011873 |
| Red blood cells | Abnormal erythrocyte morphology | HP:0001877 |
| Red cell distribution width | Abnormal erythrocyte morphology | HP:0001877 |
| White blood cells | Abnormal leukocyte count | HP:0011893 |
| Type 1 diabetes | Increased inflammatory response | HP:0012649 |
| Type 2 diabetes | Diabetes mellitus | HP:0000819 |
| C-reactive protein | Abnormal C-reactive protein level | HP:0032436 |
| Educational attainment | Neurodevelopmental abnormality | HP:0012759 |
| Amyotrophic lateral sclerosis | Motor neuron atrophy | HP:0007373 |
| Colorectal cancer | Neoplasm | HP:0002664 |
| Endometrial cancer | Neoplasm | HP:0002664 |

**Table S4. Enrichment of GWAS genes and key genes per trait**

Provided separately

***Data S1. Key gene prediction and pathway enrichments of the 44 tested traits and diseases***

https://doi.org/10.6084/m9.figshare.16866193.v1

***Data S2. HPO enrichments of genes with significant PASCAL gene p-values***

https://doi.org/10.6084/m9.figshare.16866193.v1

***Data S3. HPO enrichments of genes closest to the GWAS lead variants***

https://doi.org/10.6084/m9.figshare.16866193.v1

***Data S4. HPO enrichments of key genes***

https://doi.org/10.6084/m9.figshare.16866193.v1

### Note S1. Pathway enrichments using co-regulation networks

For most biological pathways, not all the participating genes are known because their identification relies on manual annotation and curation. To expand the gene memberships for pathways, we used our previously developed approach ^76^ to predict additional pathway members (Methods). We did this for pathways and gene sets from Reactome, the Kyoto encyclopaedia of genes and genomes (KEGG), gene ontology (GO), human phenotype ontology (HPO) and the mouse genome informatics phenotypes (MGI). Downstreamer then associated the GWAS gene p-value profile to the expanded pathway annotations.

These pathway enrichments recapitulated previous findings, for example the roles of T cell receptor (TCR) and cytokine signalling in CeD ^78, 79^ and the KEGG pathway representing T1D and asthma genes being significantly enriched for T1D and asthma, respectively (Data S1). However, we also observed new enrichments as a result of the increased statistical power of the predicted, as opposed to the directly annotated-pathway memberships (Fig. S6). For example, the GO biological processes for IL-2 regulation, T cell co-stimulation and cytokine production did not yield a significant enrichment for CeD when using the annotated databases, but did do so when we used the predicted annotations (Fig. S6A). We also observed a similar increase in power for certain pathways for height and IBD (Fig. S6E-L).

Many of the 44 traits shared pathways. For example, the auto-immune diseases showed consistent enrichment for pathways related to TCR and B cell signalling (Fig. S15, Data S1). We also observed other clusters for traits relating to metabolism, red blood cells, white blood cells and the cardiovascular system (Fig. S15), highlighting that the genetic underpinnings of the traits within each cluster are partially impacting the same pathways.

### Note S2. Tissue- and cell type–enrichments

Next, we associated the gene z-score profile of each GWAS to the expression profile of each of the 31,499 tissue and cell-line samples. This gave us an empirical way to estimate which tissues and cell types are likely to be relevant for a given GWAS trait at the mRNA level. Here we observed clear enrichments for relevant tissues. For example, IBD genes are strongly enriched for being expressed in immune and intestinal cells, while genes related to BMI, educational attainment and cholesterol levels were enriched in brain and liver (Fig. S7). The same patterns were also observed when assessing the average expression of genes in GTEx and the human cell atlas (Data S1). These findings recapitulate observations that these GWAS loci are enriched in epigenetic marks^11, 12^ and mRNA levels^80^.

