## Supplementary material for "Linking common and rare disease genetics through gene regulatory networks": Figure S7

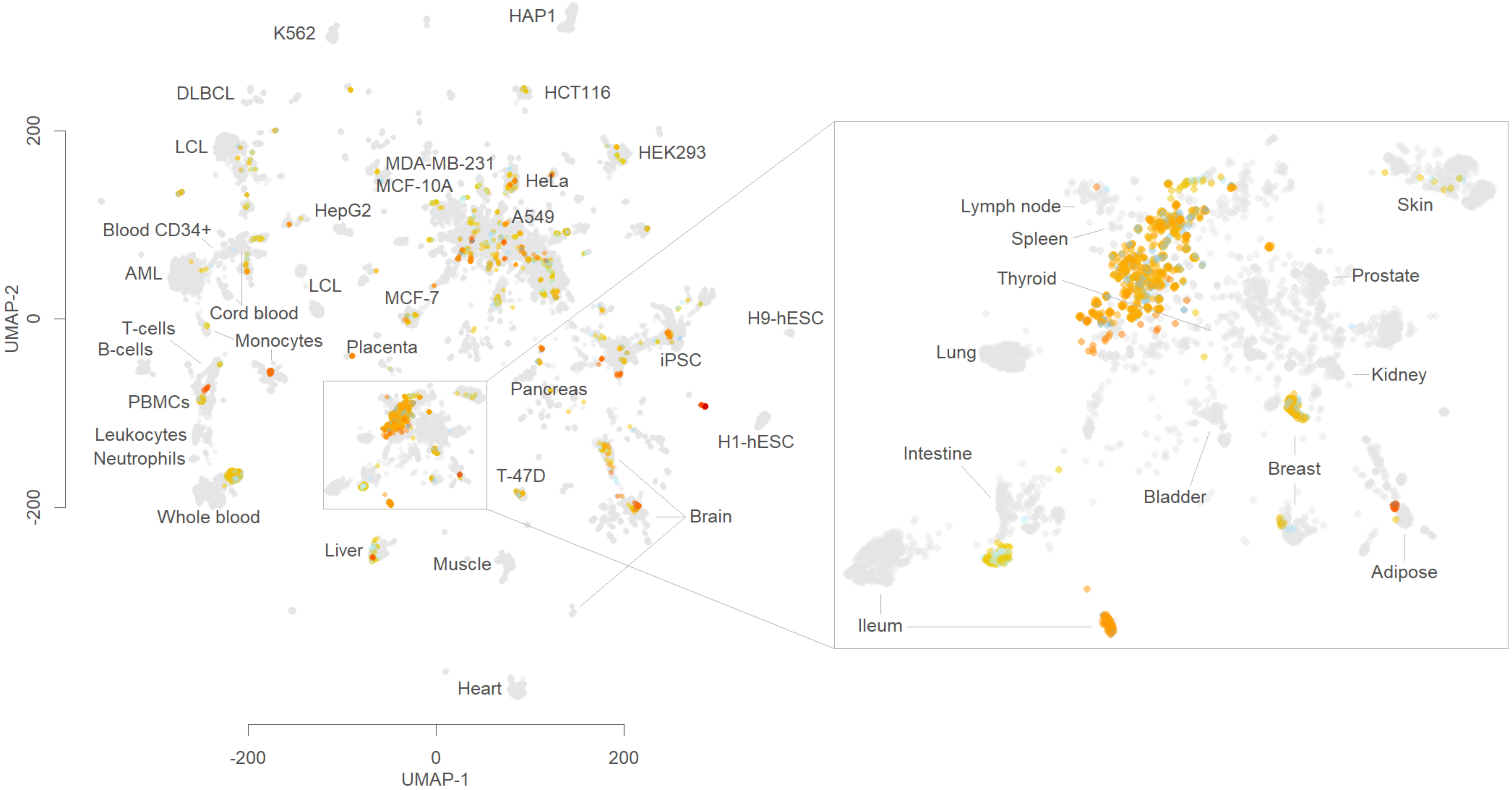

HAP1
K562
DLBCL
HCT116
LCL
HEK293
MDA-MB-231
MCF-10A
HeLa
HepG2
A549
Blood CD34+
AML
LCL
MCF-7
Cord blood
T-cells
B-cells
Monocytes
Placenta
H9-hESC
iPSC
Pancreas
PBMCs
H1-hESC
Leukocytes
Neutrophils
T-47D
Brain
Whole blood
Liver
Muscle
Heart
Lymph node
Spleen
Thyroid
Skin
Prostate
Lung
Kidney
Intestine
Breast
Bladder
Adipose
Ileum
200
0
-200
UMAP-2
-200
0
200
UMAP-1

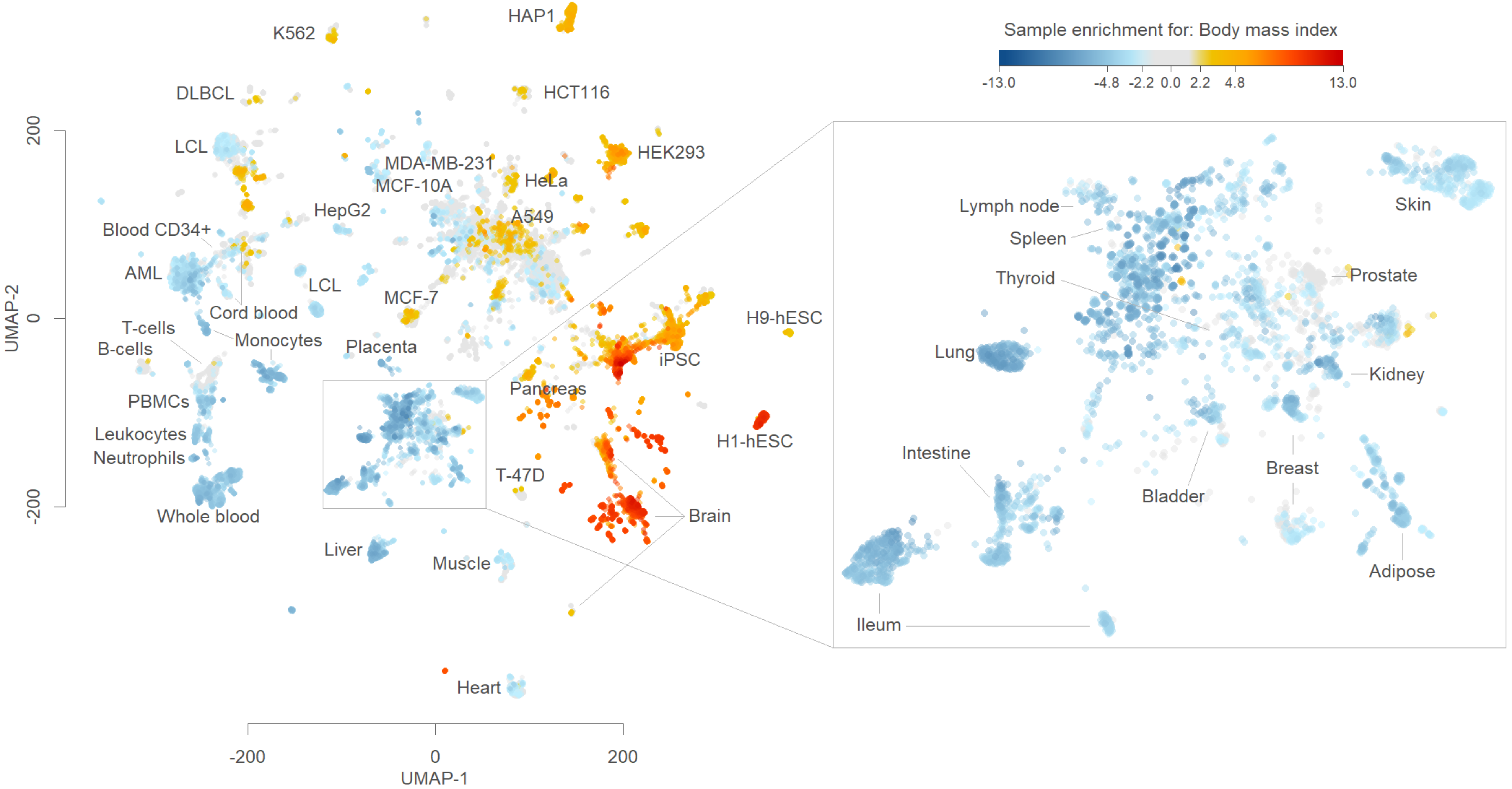

Sample enrichment for: Body mass index
-13.0
-4.8
-2.2
0.0
2.2
4.8
13.0
HAP1
K562
DLBCL
HCT116
LCL
HEK293
MDA-MB-231
MCF-10A
HeLa
HepG2
A549
Blood CD34+
AML
LCL
MCF-7
Cord blood
H9-hESC
T-cells
Monocytes
B-cells
Placenta
iPSC
PBMCs
Pancreas
Leukocytes
H1-hESC
Neutrophils
T-47D
Whole blood
Brain
Liver
Muscle
Heart
Lymph node
Spleen
Thyroid
Skin
Prostate
Lung
Kidney
Intestine
Breast
Bladder
Adipose
Ileum
UMAP-2
UMAP-1
200
0
-200

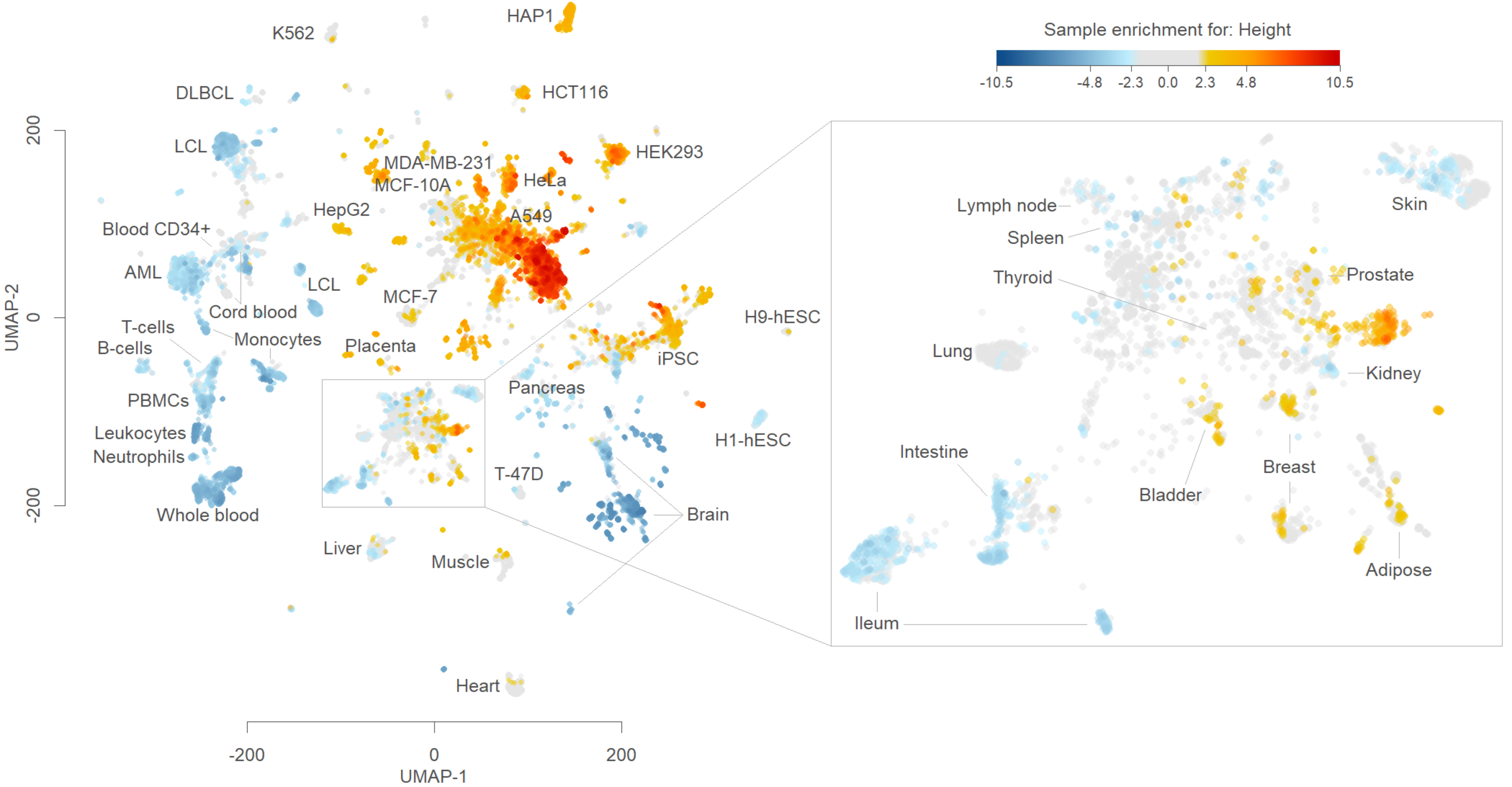

Sample enrichment for: Height
-10.5
-4.8
-2.3
0.0
2.3
4.8
10.5
K562
HAP1
DLBCL
HCT116
LCL
HEK293
MDA-MB-231
MCF-10A
HeLa
HepG2
A549
Blood CD34+
AML
LCL
Cord blood
MCF-7
T-cells
B-cells
Monocytes
Placenta
iPSC
H9-hESC
PBMCs
Pancreas
Leukocytes
Neutrophils
H1-hESC
T-47D
Brain
Whole blood
Liver
Muscle
Heart
UMAP-1
UMAP-2
-200
0
200
Lymph node
Spleen
Thyroid
Lung
Skin
Prostate
Kidney
Intestine
Bladder
Breast
Adipose
Ileum

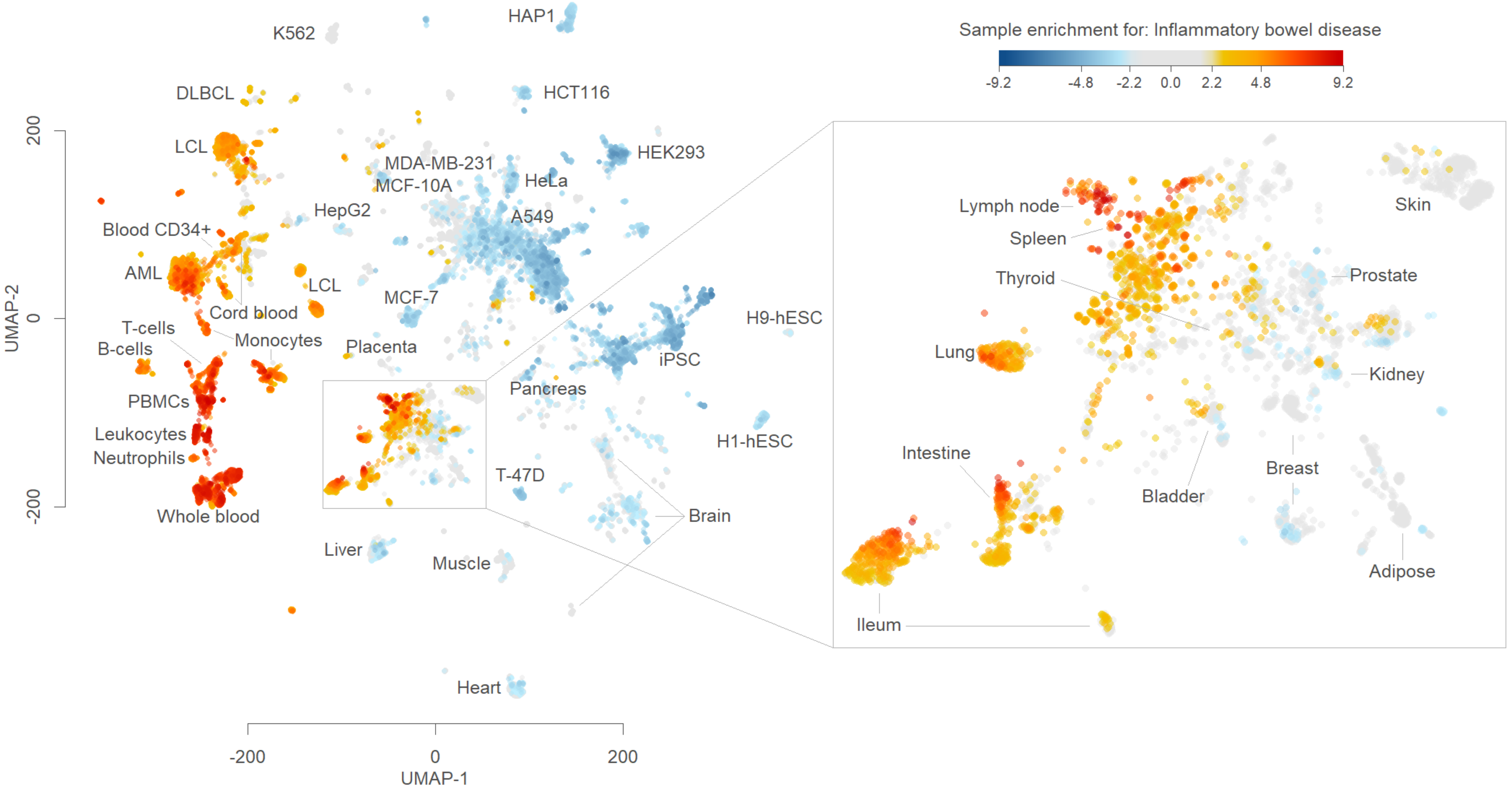

Sample enrichment for: Inflammatory bowel disease
-9.2
-4.8
-2.2
0.0
2.2
4.8
9.2
UMAP-2
UMAP-1
200
0
-200
K562
HAP1
DLBCL
HCT116
LCL
HEK293
MDA-MB-231
MCF-10A
HeLa
HepG2
A549
Blood CD34+
AML
LCL
MCF-7
Cord blood
T-cells
Monocytes
B-cells
Placenta
iPSC
H9-hESC
PBMCs
Pancreas
Leukocytes
Neutrophils
H1-hESC
T-47D
Whole blood
Brain
Liver
Muscle
Heart
Lymph node
Spleen
Thyroid
Skin
Prostate
Lung
Kidney
Intestine
Bladder
Breast
Adipose
Ileum

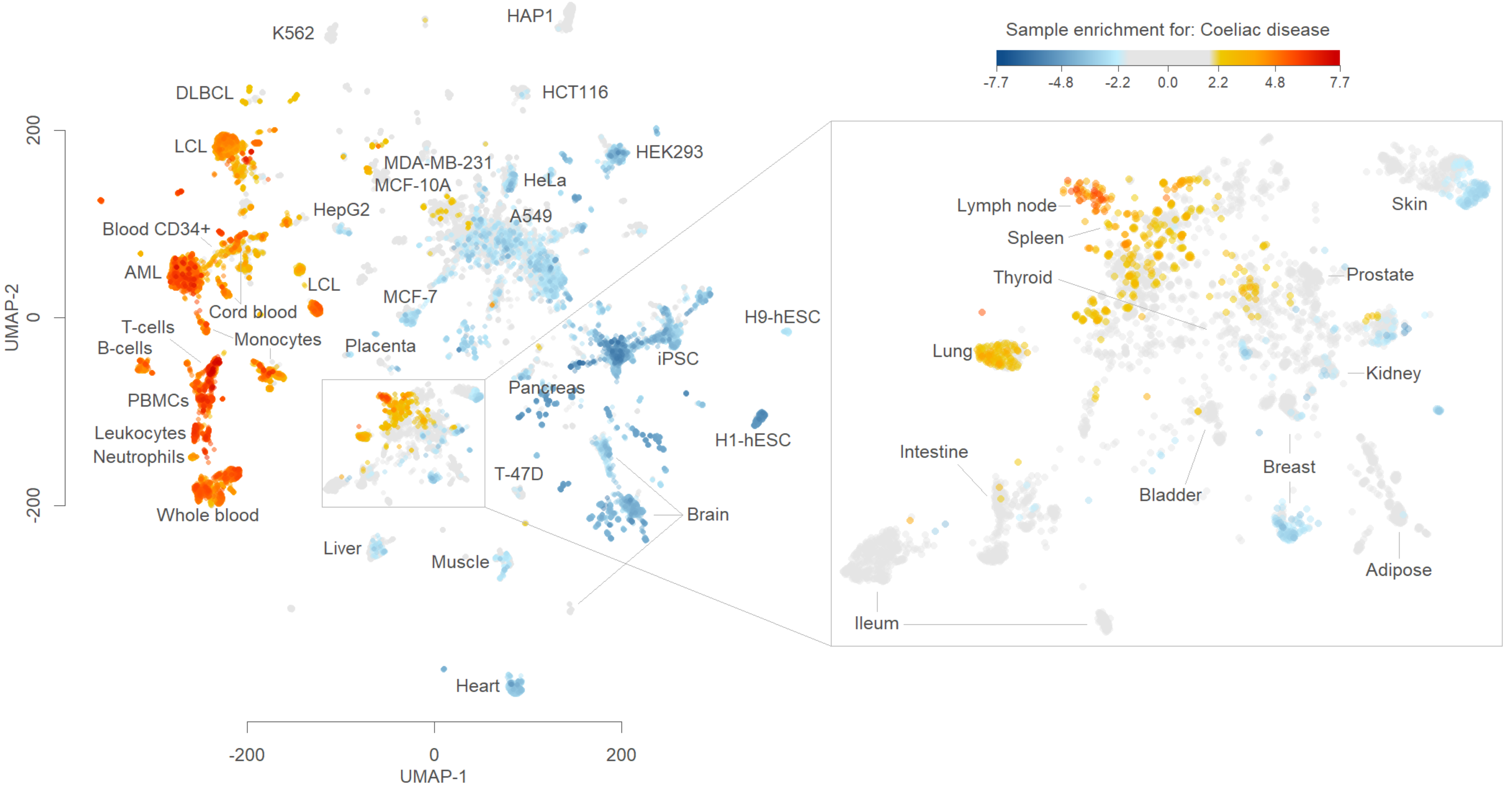

Sample enrichment for: Coeliac disease
-7.7
-4.8
-2.2
0.0
2.2
4.8
7.7
K562
HAP1
DLBCL
HCT116
LCL
HEK293
MDA-MB-231
MCF-10A
HeLa
HepG2
A549
Blood CD34+
AML
LCL
Cord blood
MCF-7
T-cells
B-cells
Monocytes
Placenta
iPSC
H9-hESC
Pancreas
PBMCs
Leukocytes
Neutrophils
H1-hESC
T-47D
Whole blood
Brain
Liver
Muscle
Heart
Lymph node
Spleen
Thyroid
Skin
Prostate
Lung
Kidney
Intestine
Bladder
Breast
Ileum
Adipose
UMAP-2
UMAP-1
200
0
-200
-200
0
200

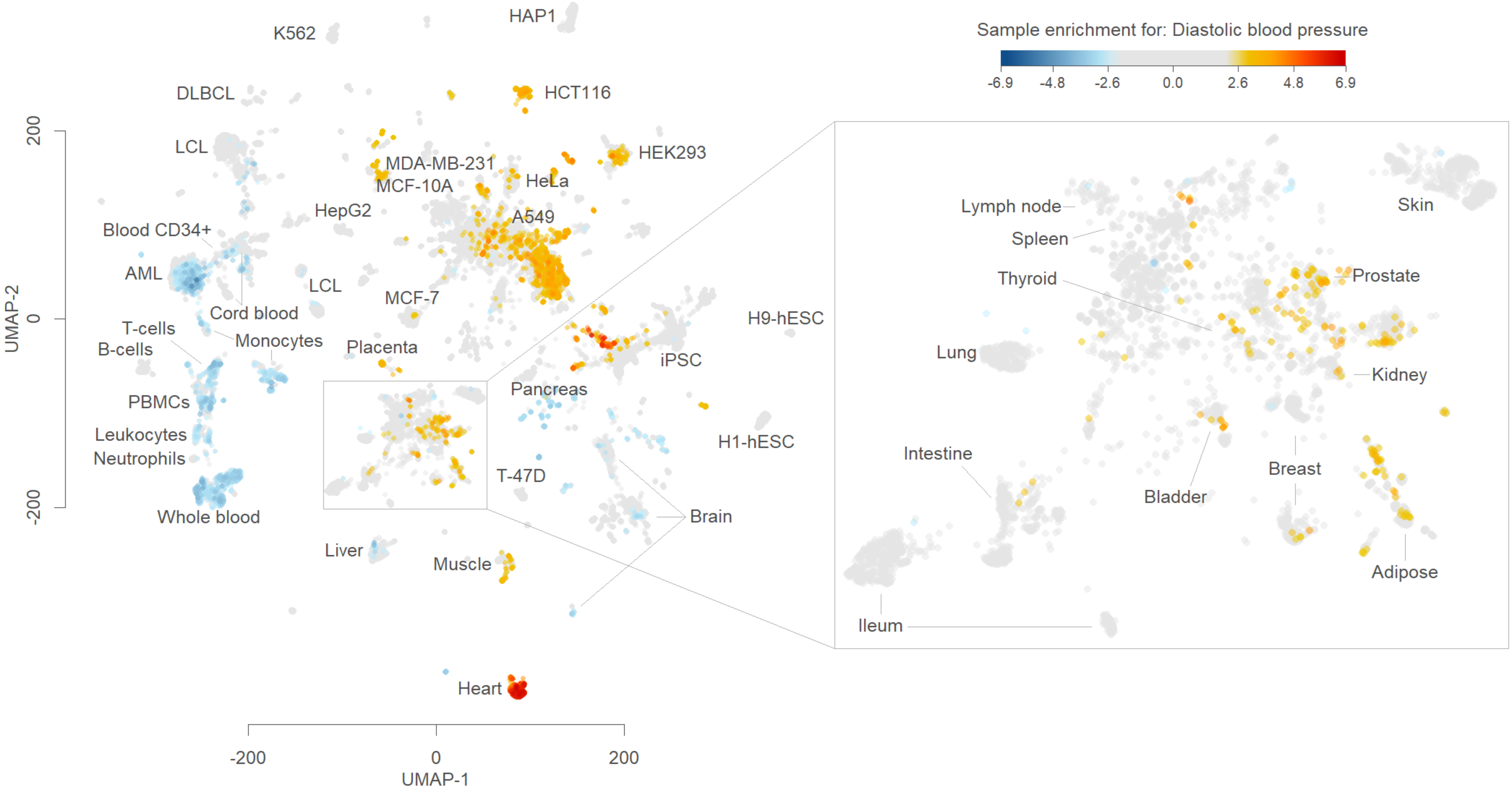

Sample enrichment for: Diastolic blood pressure
-6.9
-4.8
-2.6
0.0
2.6
4.8
6.9
HAP1
K562
DLBCL
HCT116
LCL
HEK293
MDA-MB-231
MCF-10A
HeLa
HepG2
A549
Blood CD34+
AML
LCL
MCF-7
Cord blood
T-cells
B-cells
Monocytes
Placenta
H9-hESC
iPSC
Pancreas
PBMCs
Leukocytes
Neutrophils
T-47D
H1-hESC
Brain
Whole blood
Liver
Muscle
Heart
Lymph node
Spleen
Thyroid
Lung
Intestine
Ileum
Skin
Prostate
Kidney
Bladder
Breast
Adipose
UMAP-2
200
0
-200
UMAP-1
-200
0
200

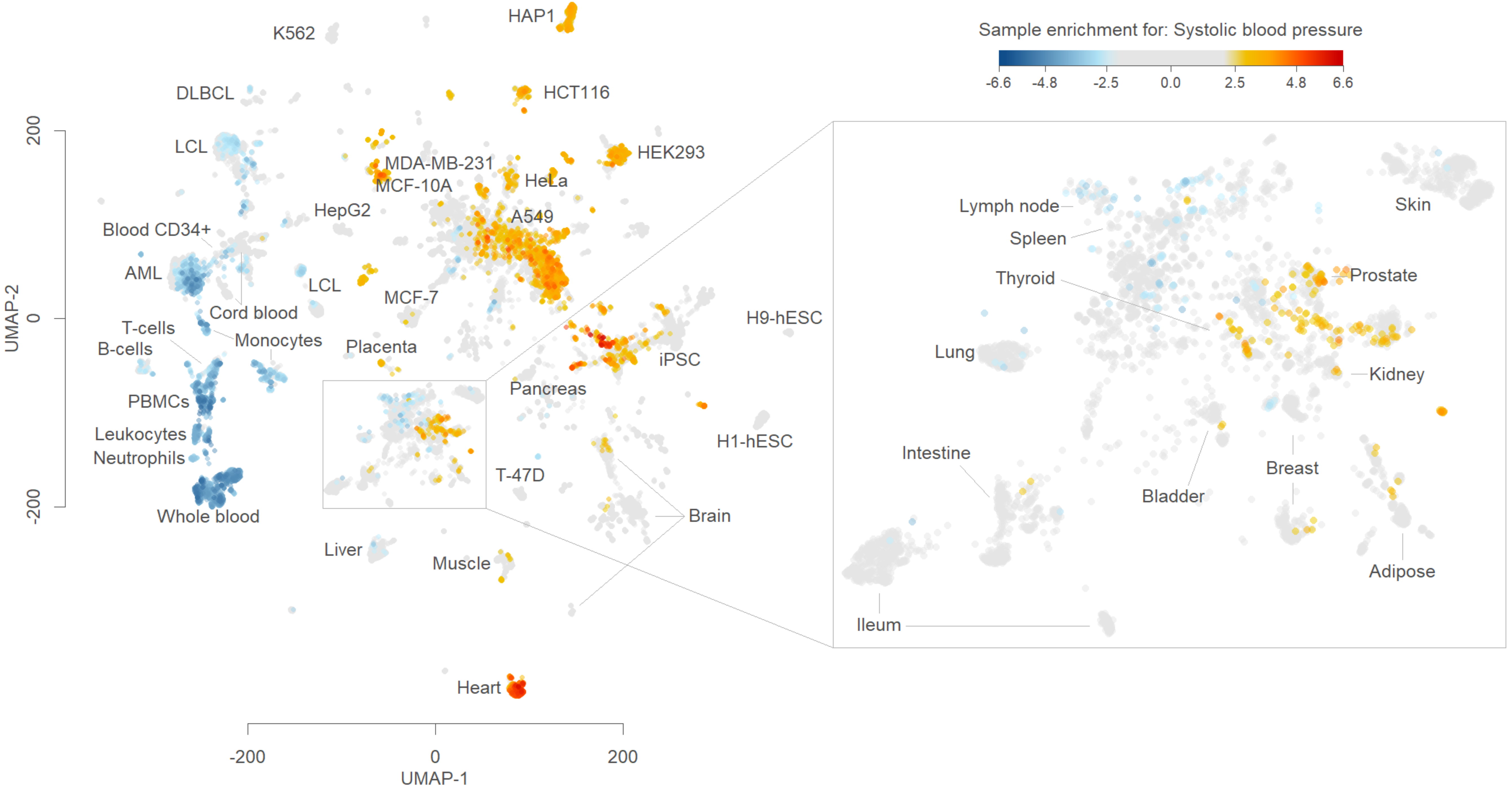
Sample enrichment for: Systolic blood pressure
-6.6
-4.8
-2.5
0.0
2.5
4.8
6.6
UMAP-2
UMAP-1
200
0
-200
-200
0
200
HAP1
K562
DLBCL
HCT116
LCL
HEK293
MDA-MB-231
MCF-10A
HeLa
HepG2
A549
Blood CD34+
AML
LCL
MCF-7
Cord blood
T-cells
B-cells
Monocytes
Placenta
H9-hESC
iPSC
PBMCs
Pancreas
Leukocytes
Neutrophils
H1-hESC
T-47D
Whole blood
Brain
Liver
Muscle
Heart
Lymph node
Spleen
Skin
Thyroid
Prostate
Lung
Kidney
Intestine
Breast
Bladder
Adipose
Ileum

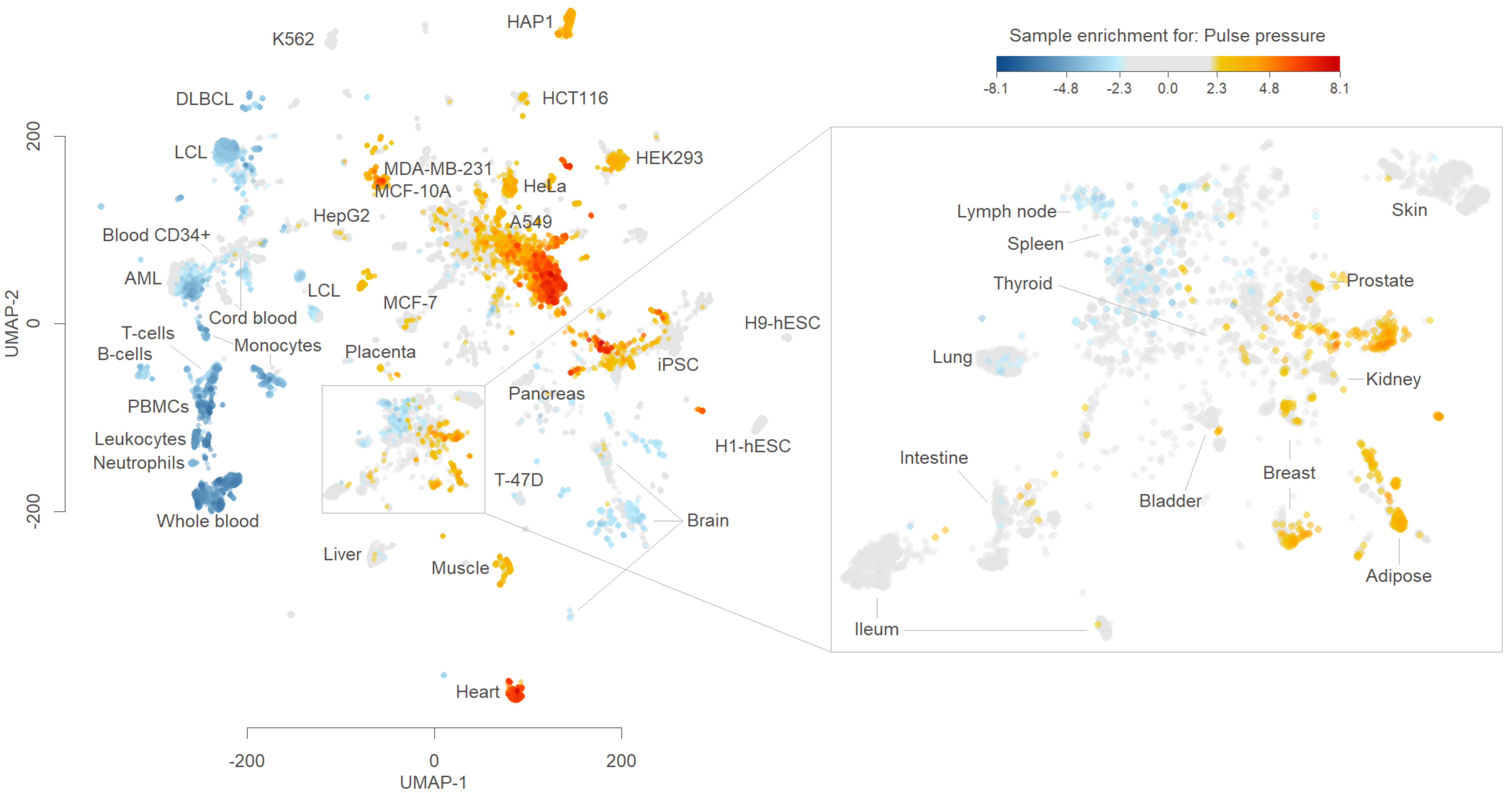
Sample enrichment for: Pulse pressure
-8.1
-4.8
-2.3
0.0
2.3
4.8
8.1
UMAP-1
UMAP-2
-200
0
200
K562
HAP1
DLBCL
HCT116
LCL
HEK293
MDA-MB-231
MCF-10A
HeLa
HepG2
A549
Blood CD34+
AML
LCL
MCF-7
Cord blood
T-cells
B-cells
Monocytes
Placenta
H9-hESC
iPSC
PBMCs
Pancreas
Leukocytes
Neutrophils
H1-hESC
T-47D
Whole blood
Brain
Liver
Muscle
Heart
Lymph node
Spleen
Thyroid
Skin
Prostate
Lung
Kidney
Intestine
Bladder
Breast
Adipose
Ileum

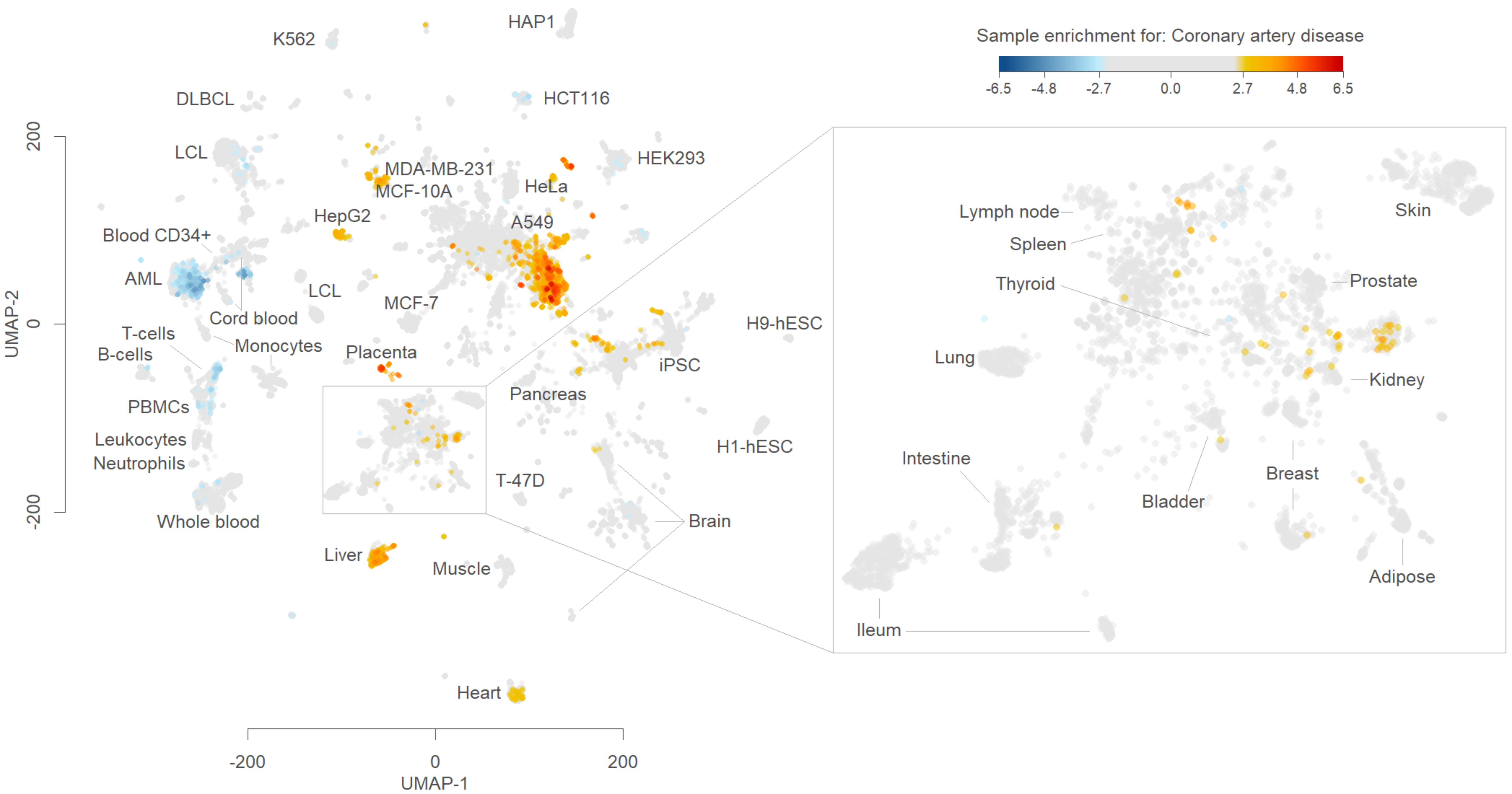
Sample enrichment for: Coronary artery disease
-6.5
-4.8
-2.7
0.0
2.7
4.8
6.5
K562
HAP1
DLBCL
HCT116
LCL
HEK293
MDA-MB-231
MCF-10A
HeLa
HepG2
A549
Blood CD34+
AML
LCL
Cord blood
MCF-7
T-cells
B-cells
Monocytes
Placenta
H9-hESC
iPSC
Pancreas
PBMCs
Leukocytes
Neutrophils
H1-hESC
T-47D
Brain
Whole blood
Liver
Muscle
Heart
UMAP-1
UMAP-2
-200
0
200
Lymph node
Spleen
Thyroid
Skin
Prostate
Lung
Kidney
Intestine
Breast
Bladder
Adipose
Ileum

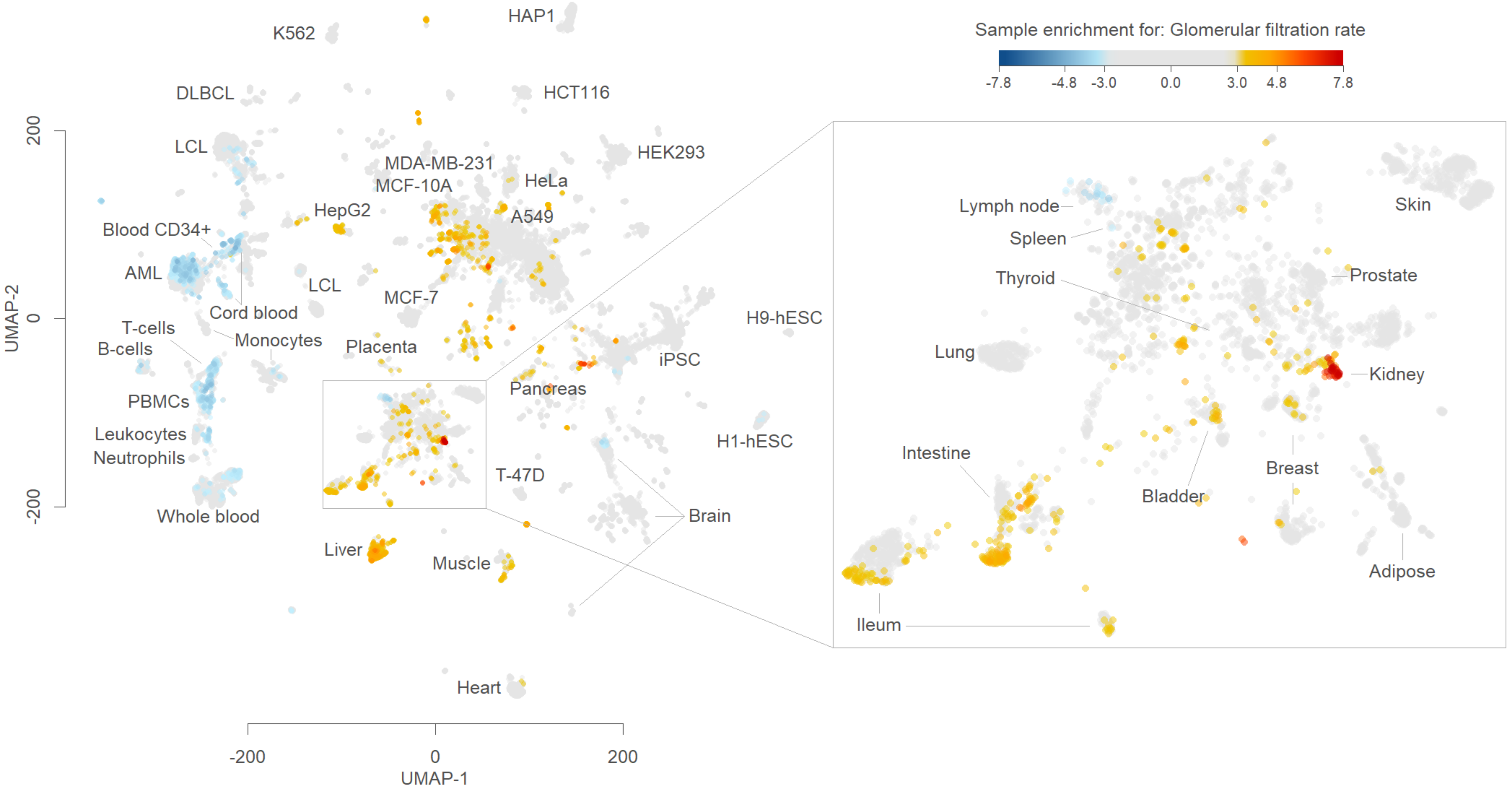
Sample enrichment for: Glomerular filtration rate
-7.8
-4.8
-3.0
0.0
3.0
4.8
7.8
UMAP-2
UMAP-1
200
0
-200
-200
0
200
K562
HAP1
DLBCL
HCT116
LCL
HEK293
MDA-MB-231
MCF-10A
HeLa
HepG2
A549
Blood CD34+
AML
LCL
MCF-7
Cord blood
H9-hESC
T-cells
B-cells
Monocytes
Placenta
iPSC
PBMCs
Pancreas
Leukocytes
Neutrophils
H1-hESC
T-47D
Whole blood
Brain
Liver
Muscle
Heart
Lymph node
Spleen
Thyroid
Skin
Prostate
Lung
Kidney
Intestine
Breast
Bladder
Adipose
Ileum

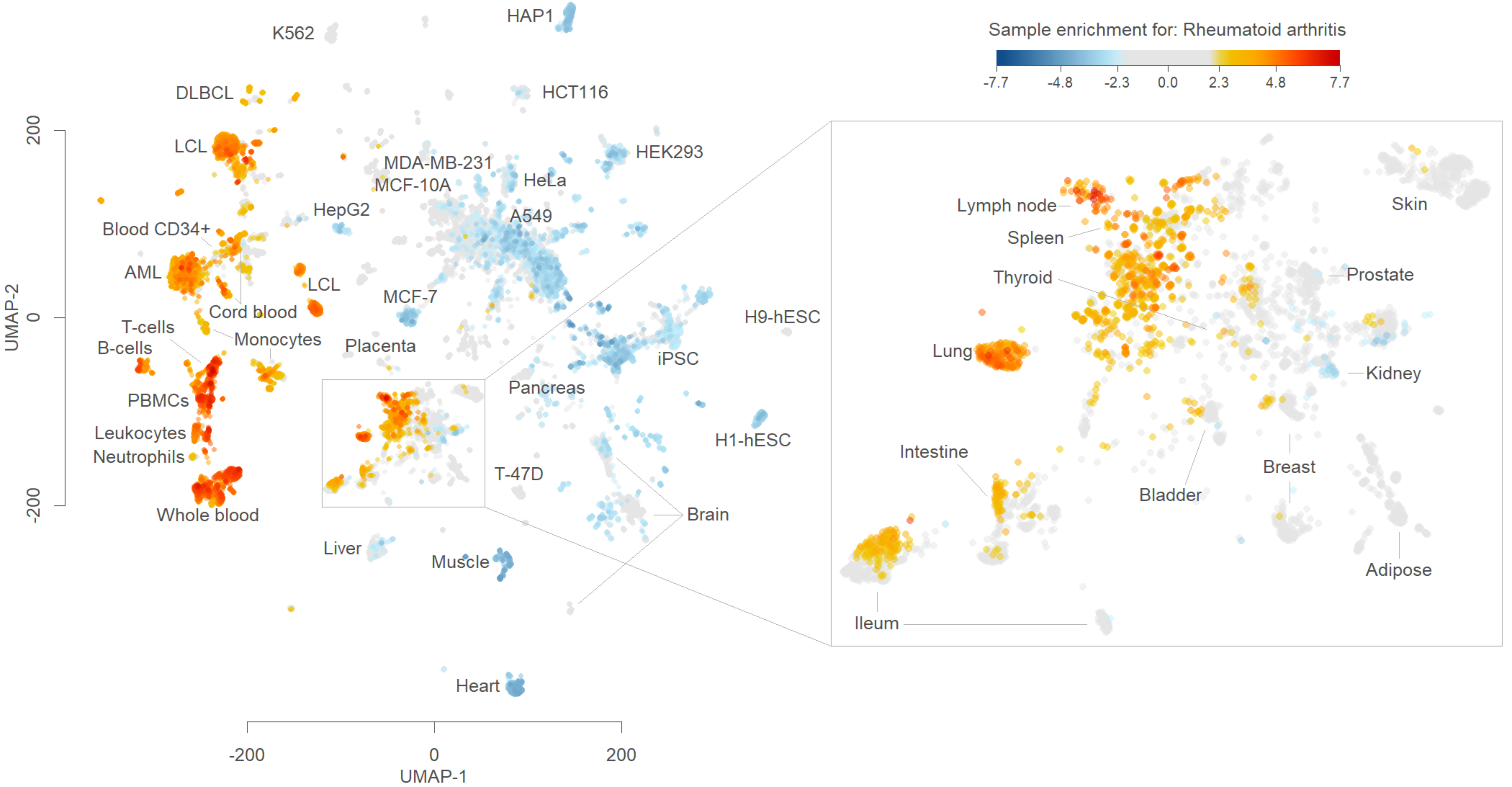
Sample enrichment for: Rheumatoid arthritis
-7.7
-4.8
-2.3
0.0
2.3
4.8
7.7
HAP1
K562
HCT116
DLBCL
HEK293
LCL
MDA-MB-231
HeLa
MCF-10A
HepG2
A549
Blood CD34+
AML
LCL
MCF-7
Cord blood
T-cells
Monocytes
B-cells
Placenta
iPSC
H9-hESC
PBMCs
Pancreas
Leukocytes
H1-hESC
Neutrophils
T-47D
Whole blood
Brain
Liver
Muscle
Heart
Lymph node
Spleen
Thyroid
Lung
Skin
Prostate
Kidney
Intestine
Bladder
Breast
Adipose
Ileum
200
0
-200
UMAP-2
-200
0
200
UMAP-1

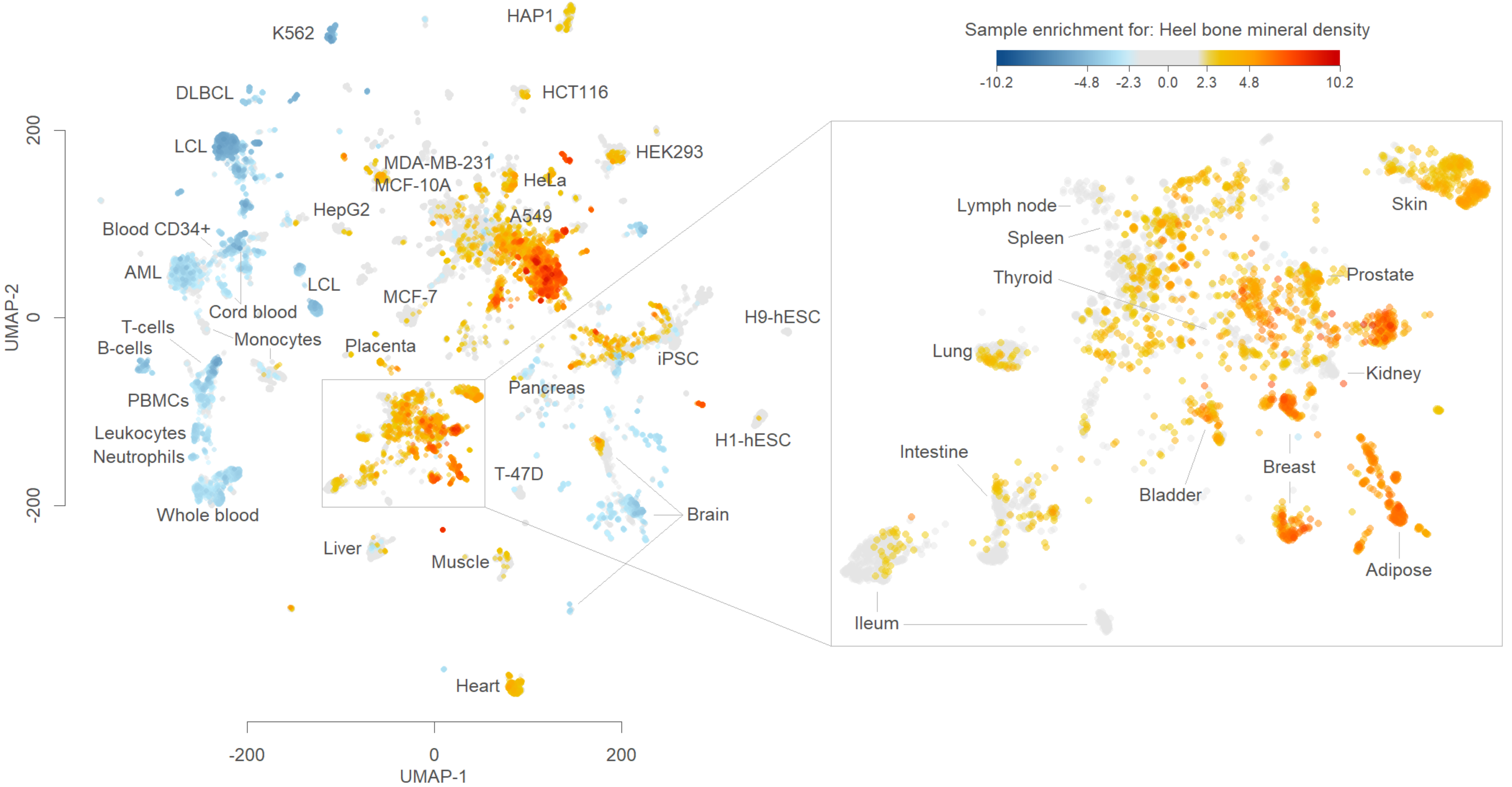

Sample enrichment for: Heel bone mineral density
-10.2
-4.8
-2.3
0.0
2.3
4.8
10.2
K562
HAP1
DLBCL
HCT116
LCL
HEK293
MDA-MB-231
MCF-10A
HeLa
HepG2
A549
Blood CD34+
AML
LCL
MCF-7
Cord blood
T-cells
B-cells
Monocytes
Placenta
iPSC
H9-hESC
PBMCs
Pancreas
Leukocytes
Neutrophils
H1-hESC
T-47D
Brain
Whole blood
Liver
Muscle
Heart
UMAP-2
200
0
-200
UMAP-1
-200
0
200
Lymph node
Spleen
Thyroid
Skin
Prostate
Lung
Kidney
Intestine
Breast
Bladder
Adipose
Ileum

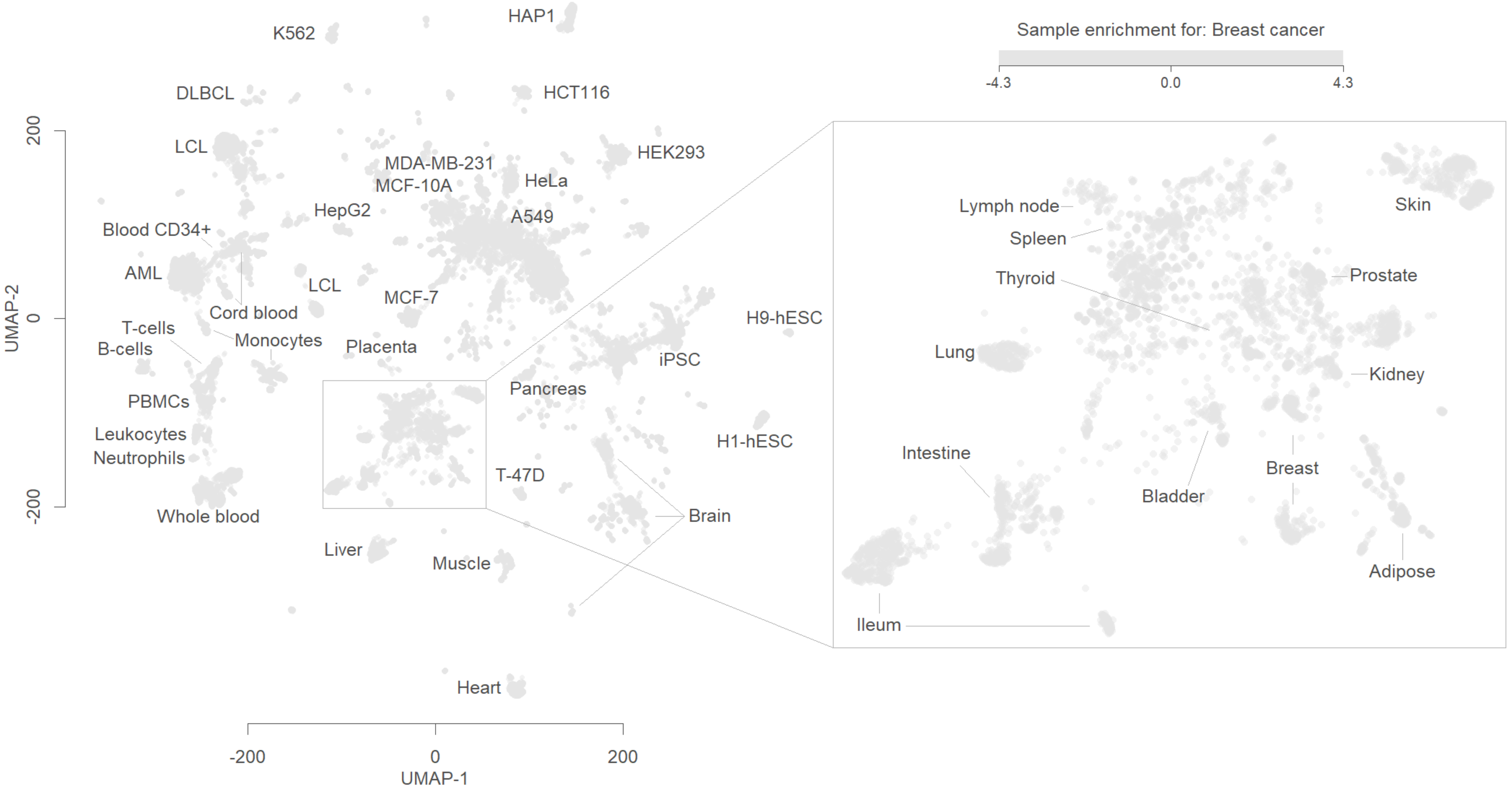

Sample enrichment for: Breast cancer
-4.3
0.0
4.3
UMAP-2
UMAP-1
200
0
-200
-200
0
200
K562
HAP1
DLBCL
HCT116
LCL
HEK293
MDA-MB-231
MCF-10A
HeLa
HepG2
A549
Blood CD34+
AML
LCL
MCF-7
Cord blood
T-cells
B-cells
Monocytes
Placenta
H9-hESC
iPSC
PBMCs
Pancreas
Leukocytes
Neutrophils
H1-hESC
T-47D
Brain
Whole blood
Liver
Muscle
Heart
Lymph node
Spleen
Thyroid
Skin
Prostate
Lung
Kidney
Intestine
Bladder
Breast
Adipose
Ileum

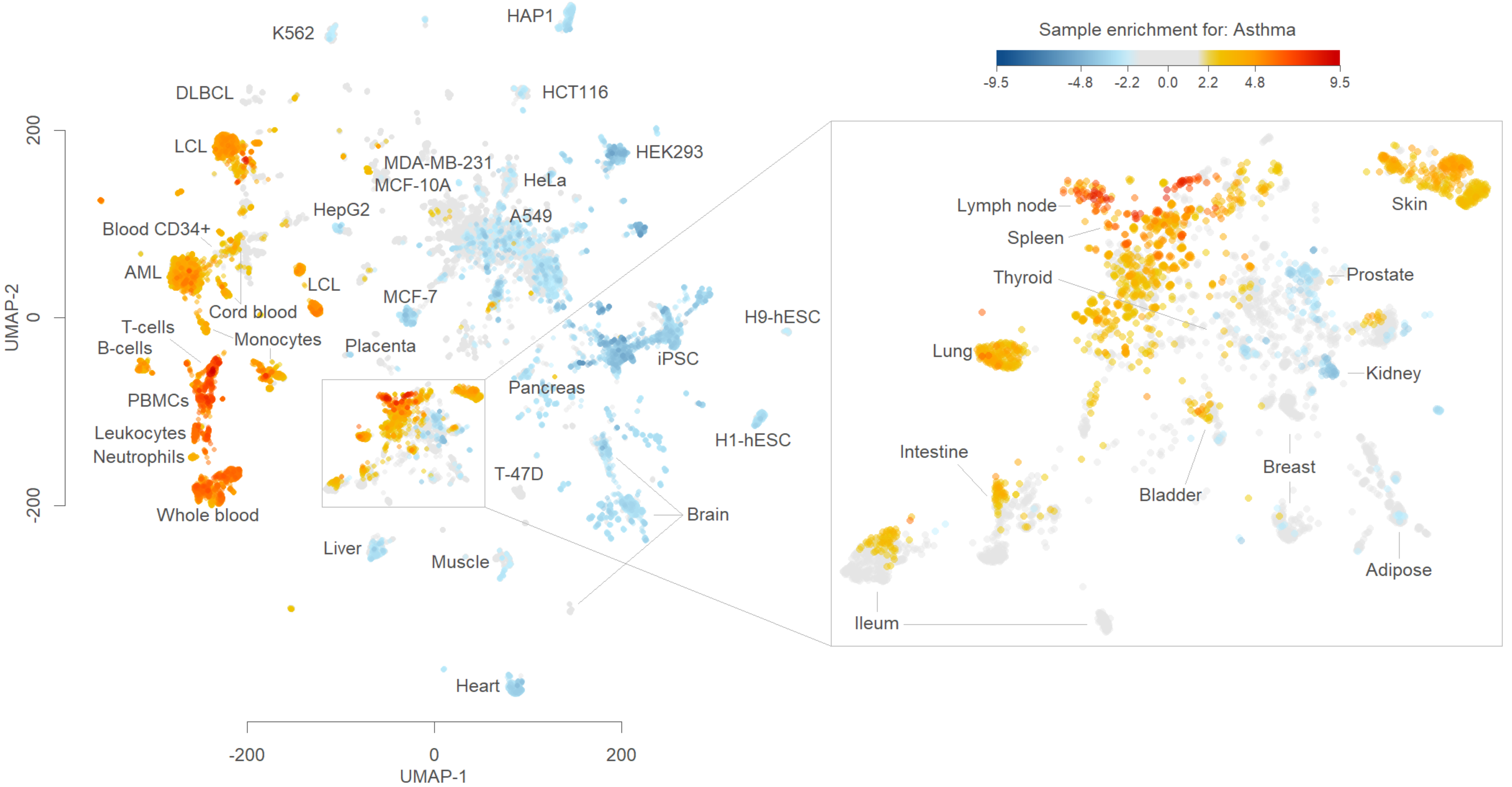
Sample enrichment for: Asthma
-9.5
-4.8
-2.2
0.0
2.2
4.8
9.5
HAP1
K562
DLBCL
HCT116
LCL
HEK293
MDA-MB-231
MCF-10A
HeLa
HepG2
A549
Blood CD34+
AML
LCL
MCF-7
Cord blood
T-cells
B-cells
Monocytes
Placenta
H9-hESC
iPSC
PBMCs
Pancreas
Leukocytes
Neutrophils
H1-hESC
T-47D
Whole blood
Brain
Liver
Muscle
Heart
UMAP-2
UMAP-1
200
0
-200
Lymph node
Spleen
Thyroid
Skin
Prostate
Lung
Kidney
Intestine
Breast
Bladder
Adipose
Ileum

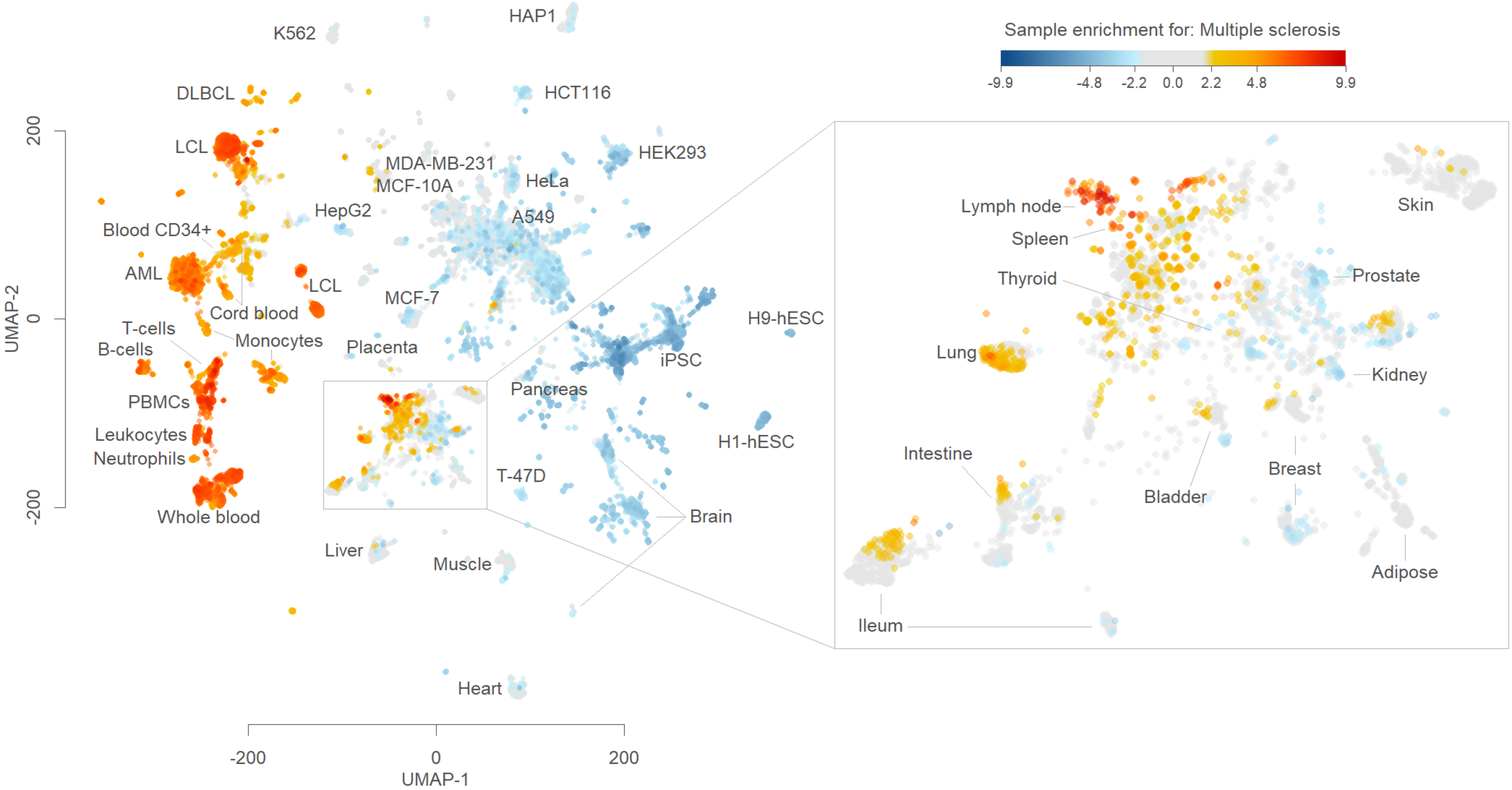

Sample enrichment for: Multiple sclerosis
-9.9
-4.8
-2.2
0.0
2.2
4.8
9.9
UMAP-2
UMAP-1
200
0
-200
-200
0
200
HAP1
K562
DLBCL
HCT116
LCL
HEK293
MDA-MB-231
MCF-10A
HeLa
HepG2
A549
Blood CD34+
AML
LCL
MCF-7
Cord blood
T-cells
Monocytes
B-cells
Placenta
iPSC
H9-hESC
PBMCs
Pancreas
Leukocytes
H1-hESC
Neutrophils
T-47D
Whole blood
Brain
Liver
Muscle
Heart
Lymph node
Spleen
Thyroid
Skin
Prostate
Lung
Kidney
Intestine
Breast
Bladder
Adipose
Ileum

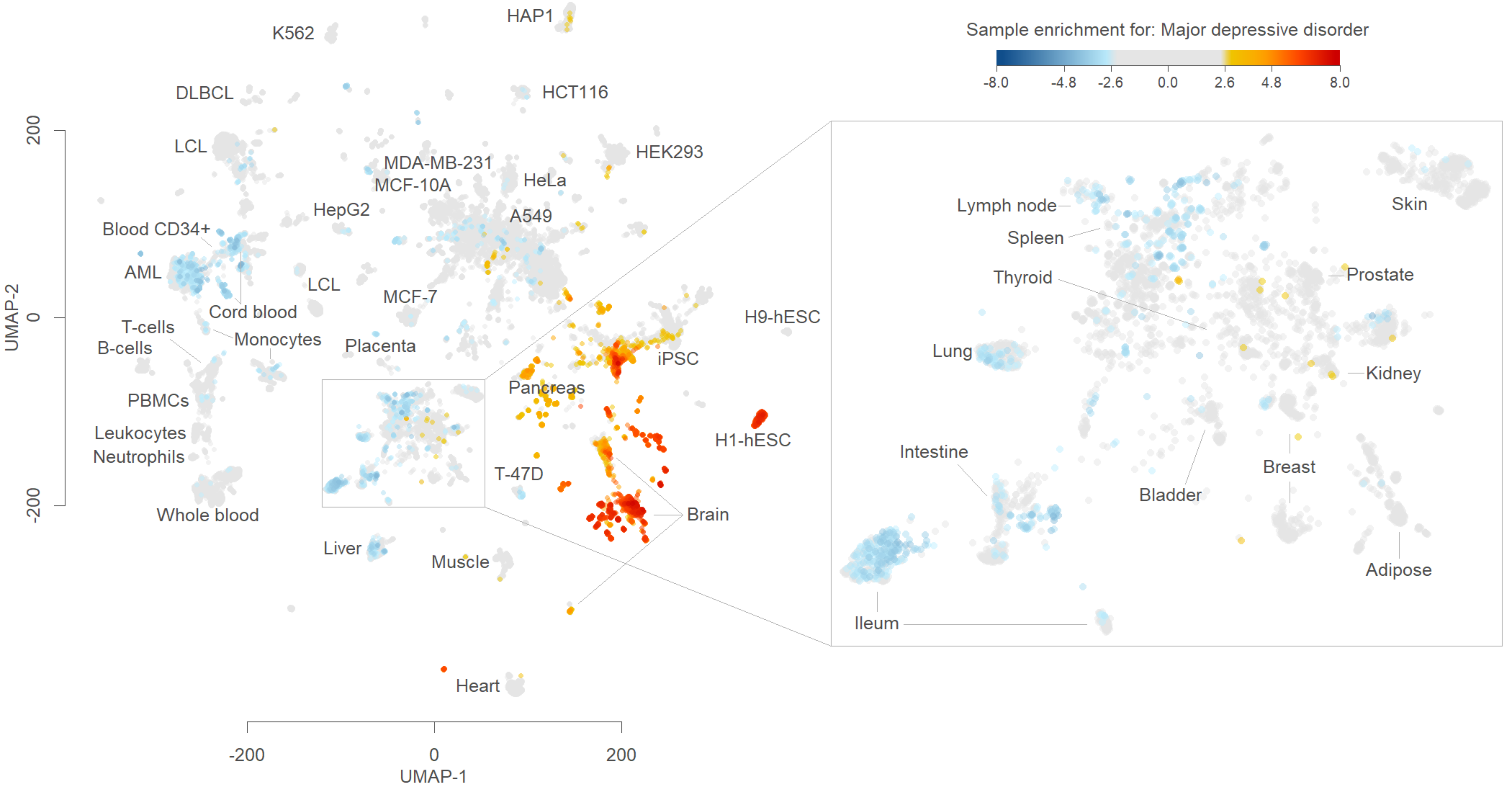

Sample enrichment for: Major depressive disorder
-8.0
-4.8
-2.6
0.0
2.6
4.8
8.0
UMAP-2
UMAP-1
200
0
-200
-200
0
200
K562
HAP1
DLBCL
HCT116
LCL
HEK293
MDA-MB-231
MCF-10A
HeLa
HepG2
A549
Blood CD34+
AML
LCL
Cord blood
MCF-7
T-cells
B-cells
Monocytes
Placenta
iPSC
H9-hESC
PBMCs
Pancreas
Leukocytes
Neutrophils
H1-hESC
T-47D
Whole blood
Brain
Liver
Muscle
Heart
Lymph node
Spleen
Thyroid
Skin
Prostate
Lung
Kidney
Intestine
Breast
Bladder
Adipose
Ileum

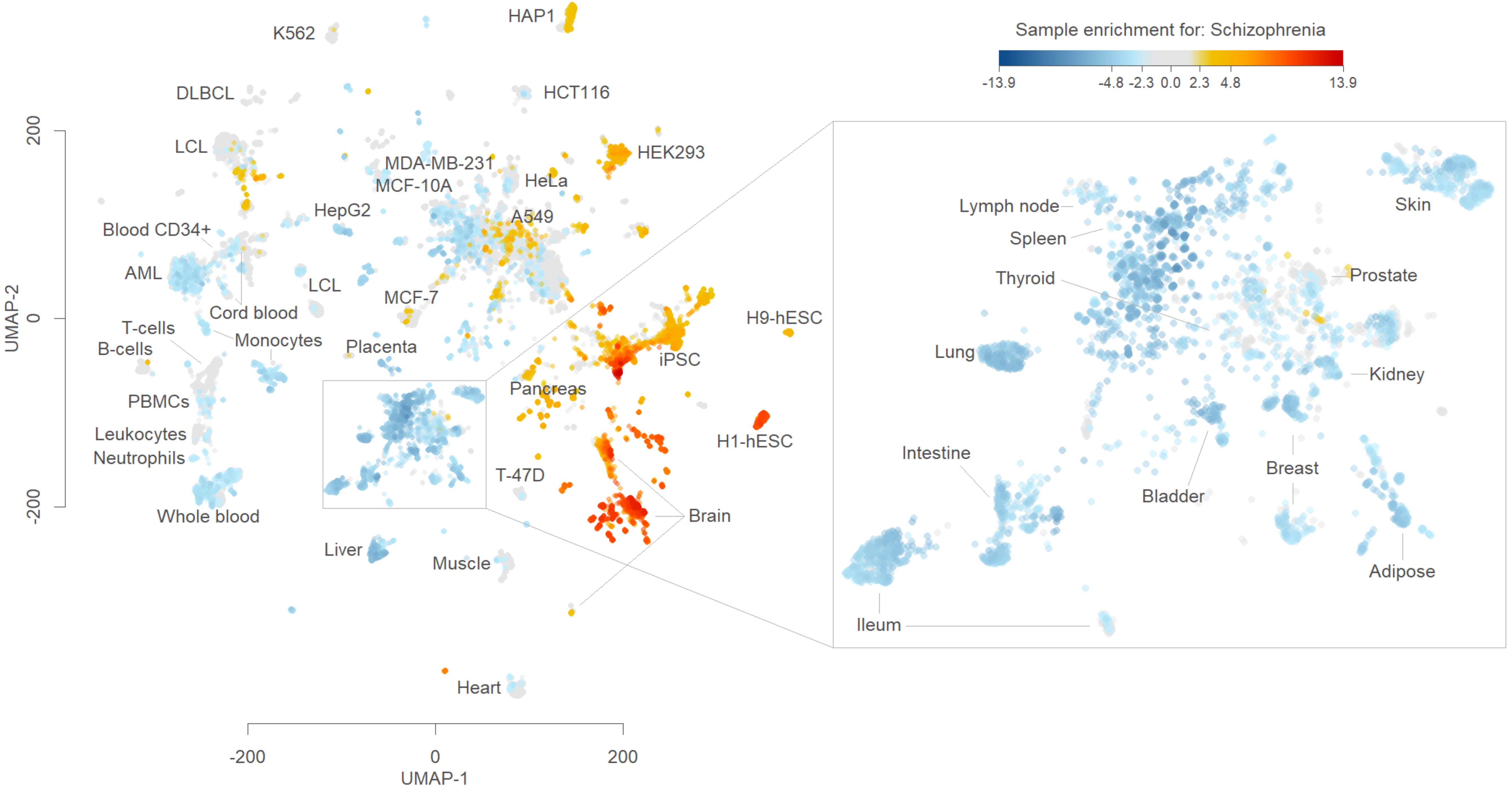
Sample enrichment for: Schizophrenia
-13.9
-4.8
-2.3
0.0
2.3
4.8
13.9
K562
HAP1
DLBCL
HCT116
LCL
HEK293
MDA-MB-231
HeLa
MCF-10A
HepG2
A549
Blood CD34+
AML
LCL
MCF-7
Cord blood
T-cells
Monocytes
B-cells
Placenta
iPSC
H9-hESC
Pancreas
PBMCs
Leukocytes
Neutrophils
H1-hESC
T-47D
Brain
Whole blood
Liver
Muscle
Heart
Lymph node
Spleen
Thyroid
Skin
Prostate
Lung
Kidney
Intestine
Breast
Bladder
Adipose
Ileum
UMAP-1
UMAP-2
-200
0
200

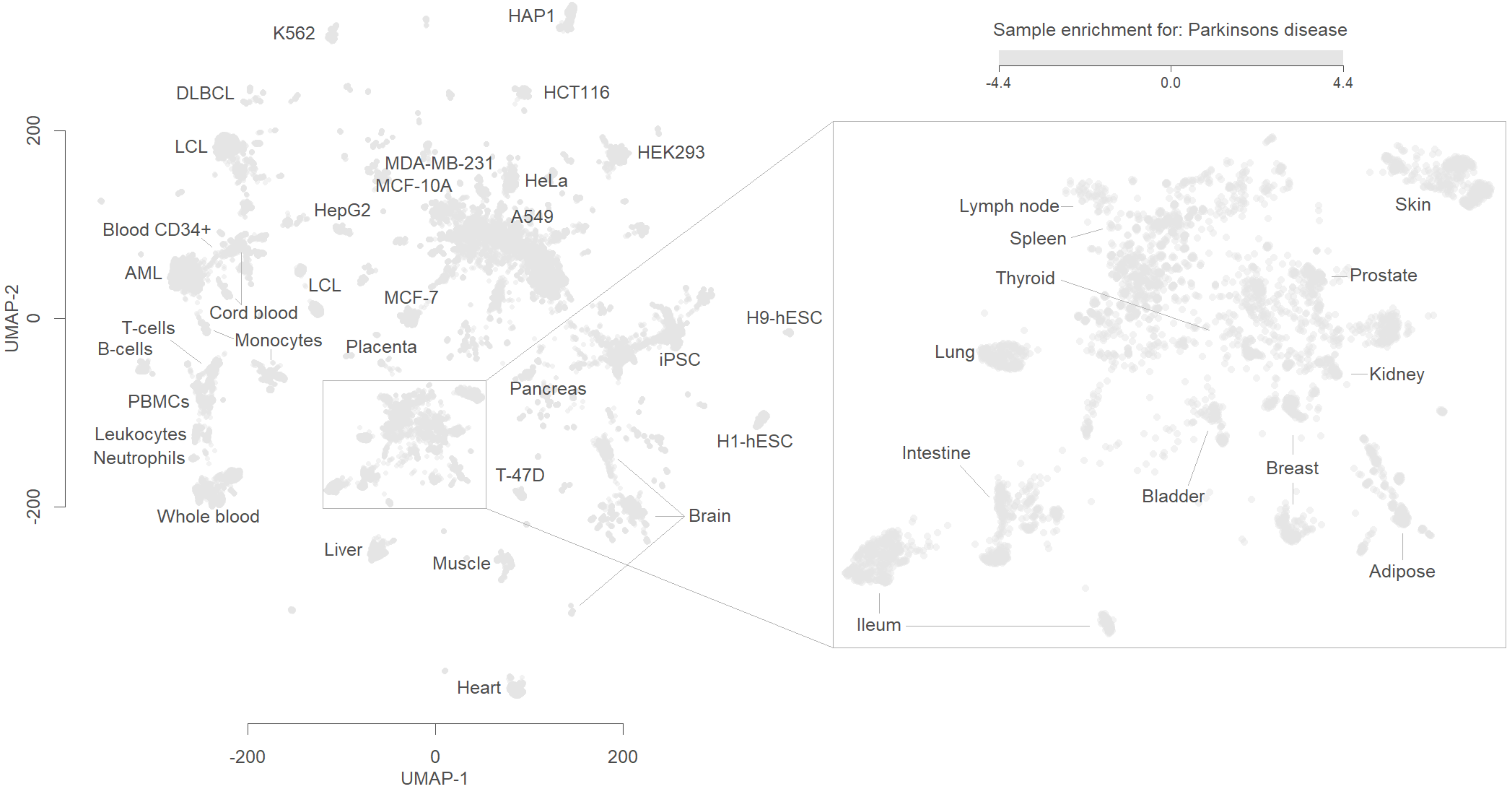
Sample enrichment for: Parkinsons disease
-4.4
0.0
4.4
HAP1
K562
DLBCL
HCT116
LCL
HEK293
MDA-MB-231
MCF-10A
HeLa
HepG2
A549
Blood CD34+
AML
LCL
MCF-7
Cord blood
T-cells
B-cells
Monocytes
Placenta
PBMCs
Leukocytes
Neutrophils
Whole blood
H9-hESC
iPSC
Pancreas
H1-hESC
T-47D
Brain
Liver
Muscle
Heart
Lymph node
Spleen
Thyroid
Skin
Prostate
Lung
Kidney
Intestine
Breast
Bladder
Adipose
Ileum
UMAP-2
200
0
-200
-200
0
200
UMAP-1

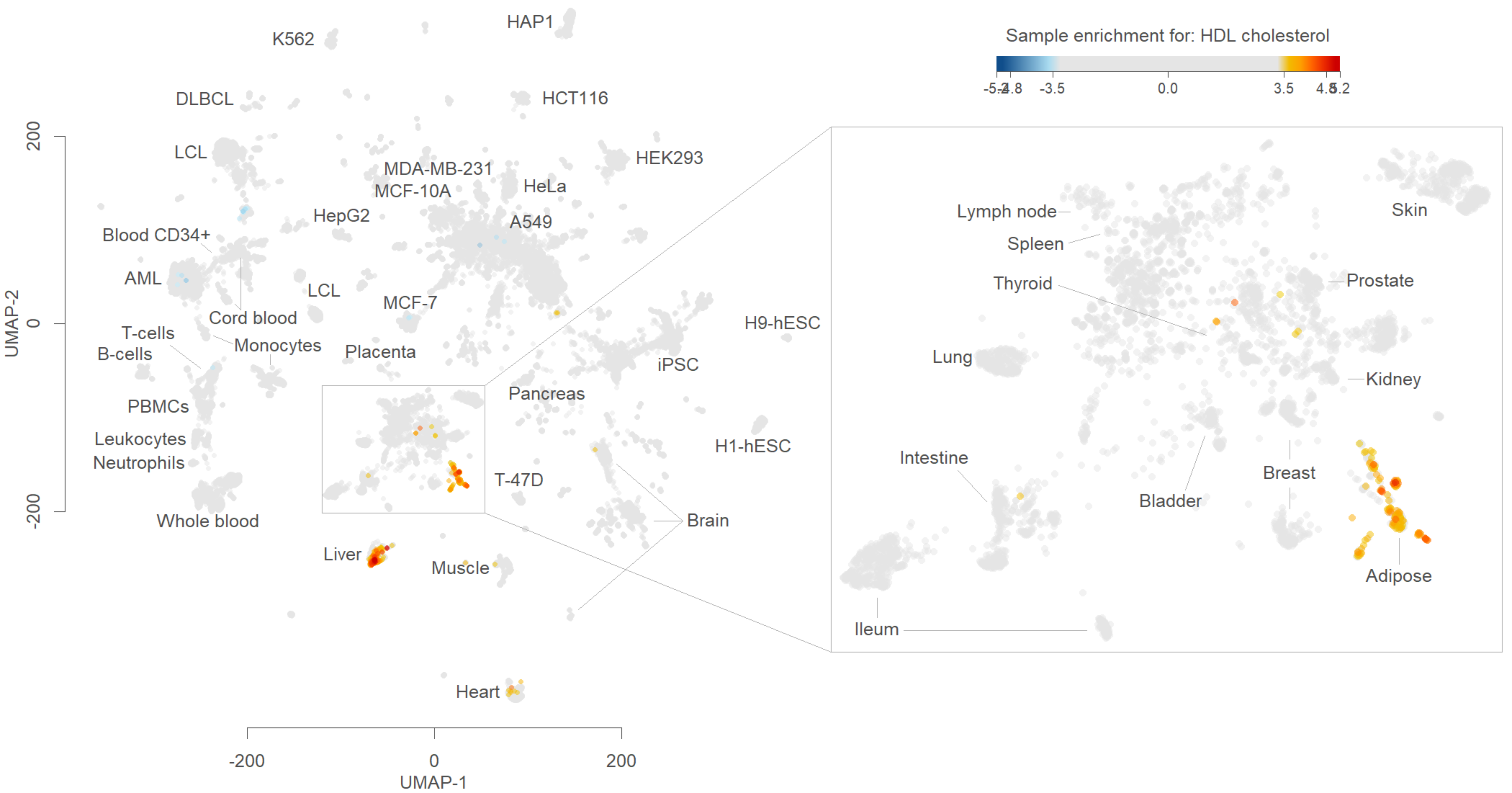

Sample enrichment for: HDL cholesterol
-5.2
-4.8
-3.5
0.0
3.5
4.8
5.2
HAP1
K562
DLBCL
HCT116
LCL
MDA-MB-231
MCF-10A
HeLa
HEK293
HepG2
A549
Blood CD34+
AML
LCL
MCF-7
Cord blood
T-cells
B-cells
Monocytes
Placenta
H9-hESC
iPSC
PBMCs
Pancreas
Leukocytes
Neutrophils
H1-hESC
T-47D
Whole blood
Brain
Liver
Muscle
Heart
UMAP-2
UMAP-1
200
0
-200
-200
0
200
Lymph node
Spleen
Thyroid
Skin
Prostate
Lung
Kidney
Intestine
Breast
Bladder
Adipose
Ileum

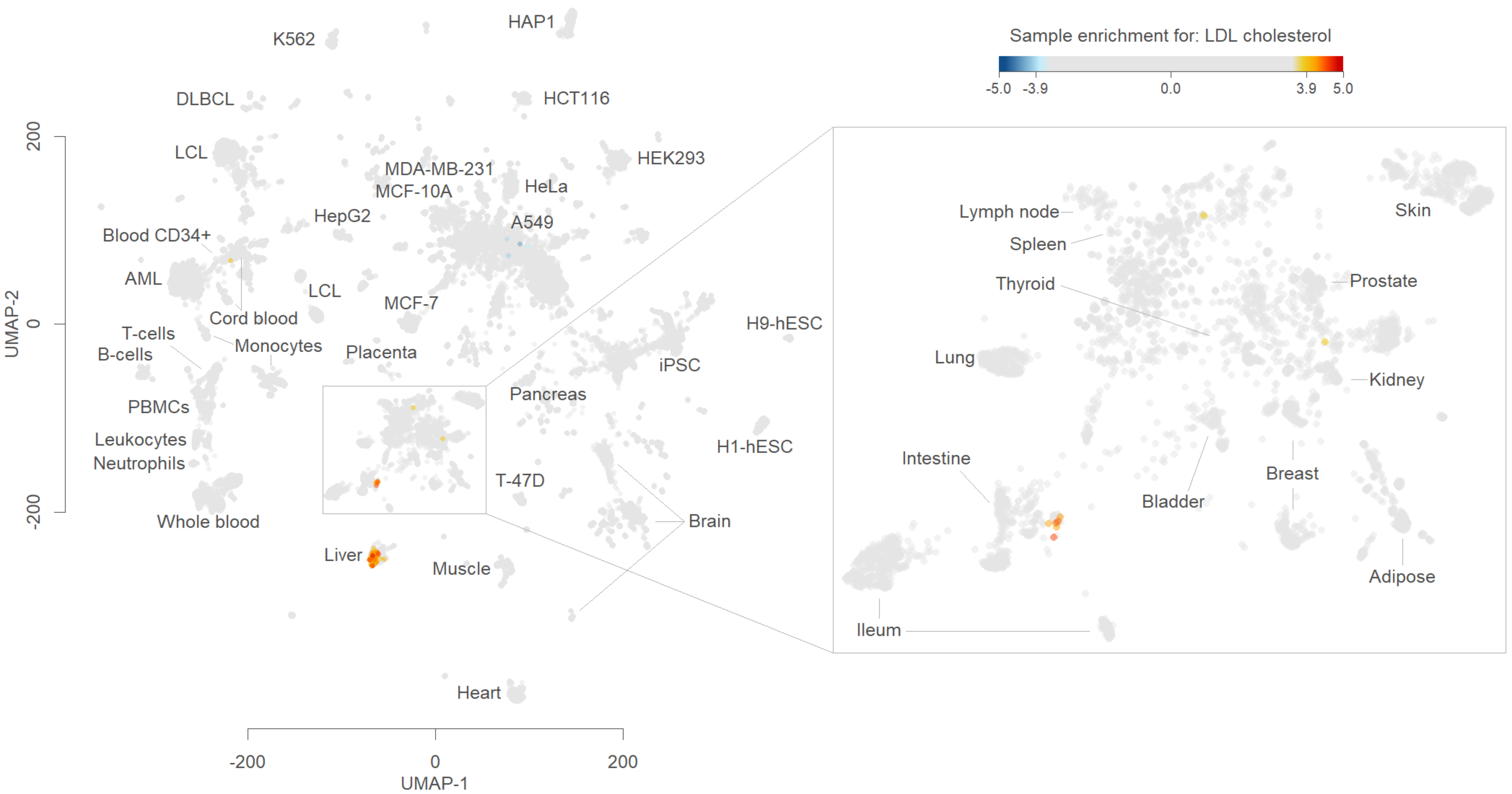

Sample enrichment for: LDL cholesterol
-5.0
-3.9
0.0
3.9
5.0
HAP1
K562
DLBCL
HCT116
LCL
HEK293
MDA-MB-231
MCF-10A
HeLa
HepG2
A549
Blood CD34+
AML
LCL
MCF-7
Cord blood
T-cells
B-cells
Monocytes
Placenta
H9-hESC
iPSC
PBMCs
Pancreas
Leukocytes
Neutrophils
H1-hESC
T-47D
Whole blood
Brain
Liver
Muscle
Heart
Lymph node
Spleen
Thyroid
Skin
Prostate
Lung
Kidney
Intestine
Bladder
Breast
Adipose
Ileum
UMAP-2
200
0
-200
UMAP-1
-200
0
200

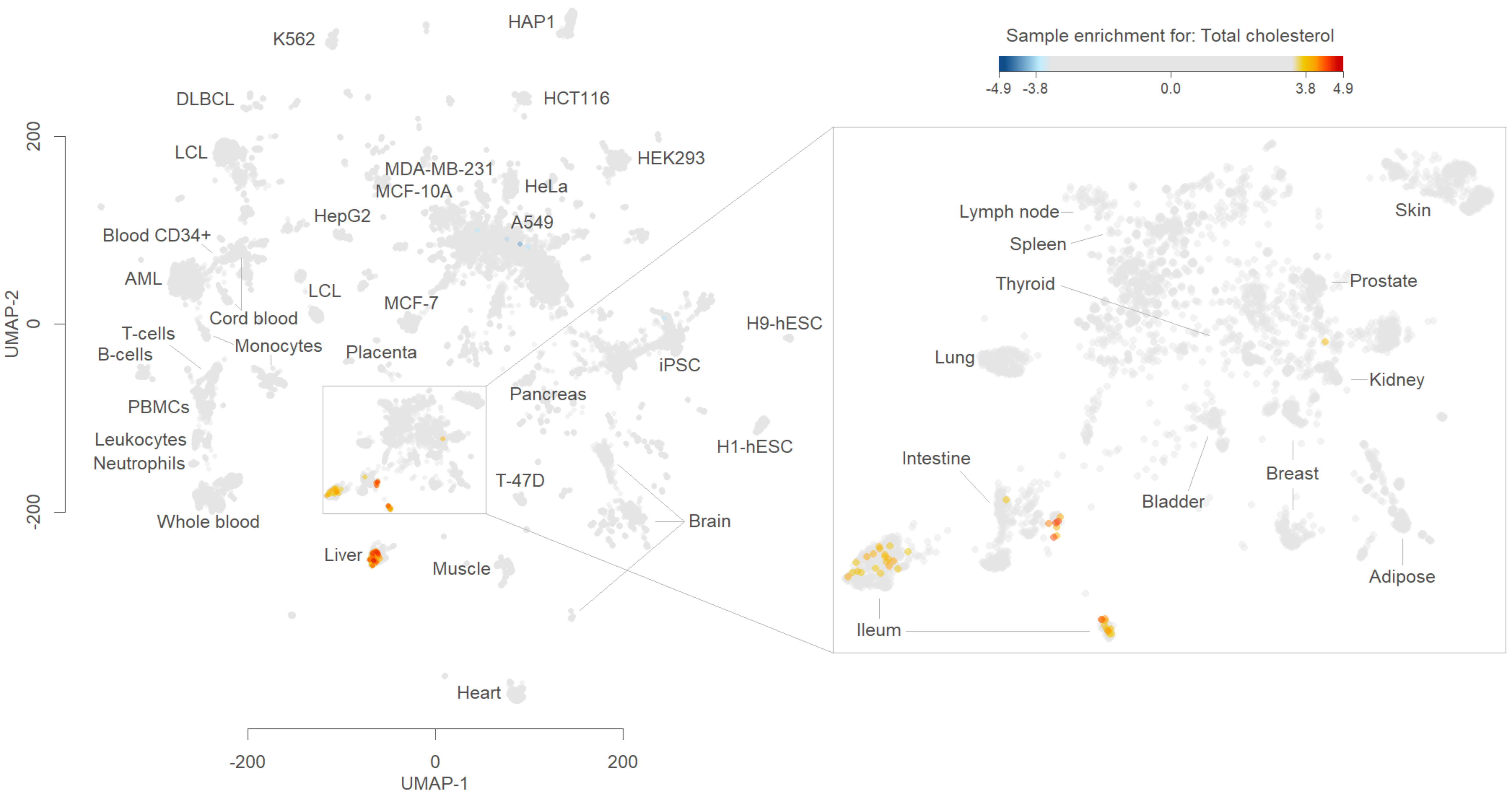
Sample enrichment for: Total cholesterol
-4.9
-3.8
0.0
3.8
4.9
UMAP-2
200
0
-200
UMAP-1
-200
0
200
HAP1
K562
DLBCL
HCT116
LCL
HEK293
MDA-MB-231
MCF-10A
HeLa
HepG2
A549
Blood CD34+
AML
LCL
MCF-7
Cord blood
T-cells
B-cells
Monocytes
Placenta
H9-hESC
iPSC
PBMCs
Pancreas
Leukocytes
Neutrophils
H1-hESC
T-47D
Whole blood
Brain
Liver
Muscle
Heart
Lymph node
Spleen
Thyroid
Skin
Prostate
Lung
Kidney
Intestine
Bladder
Breast
Adipose
Ileum

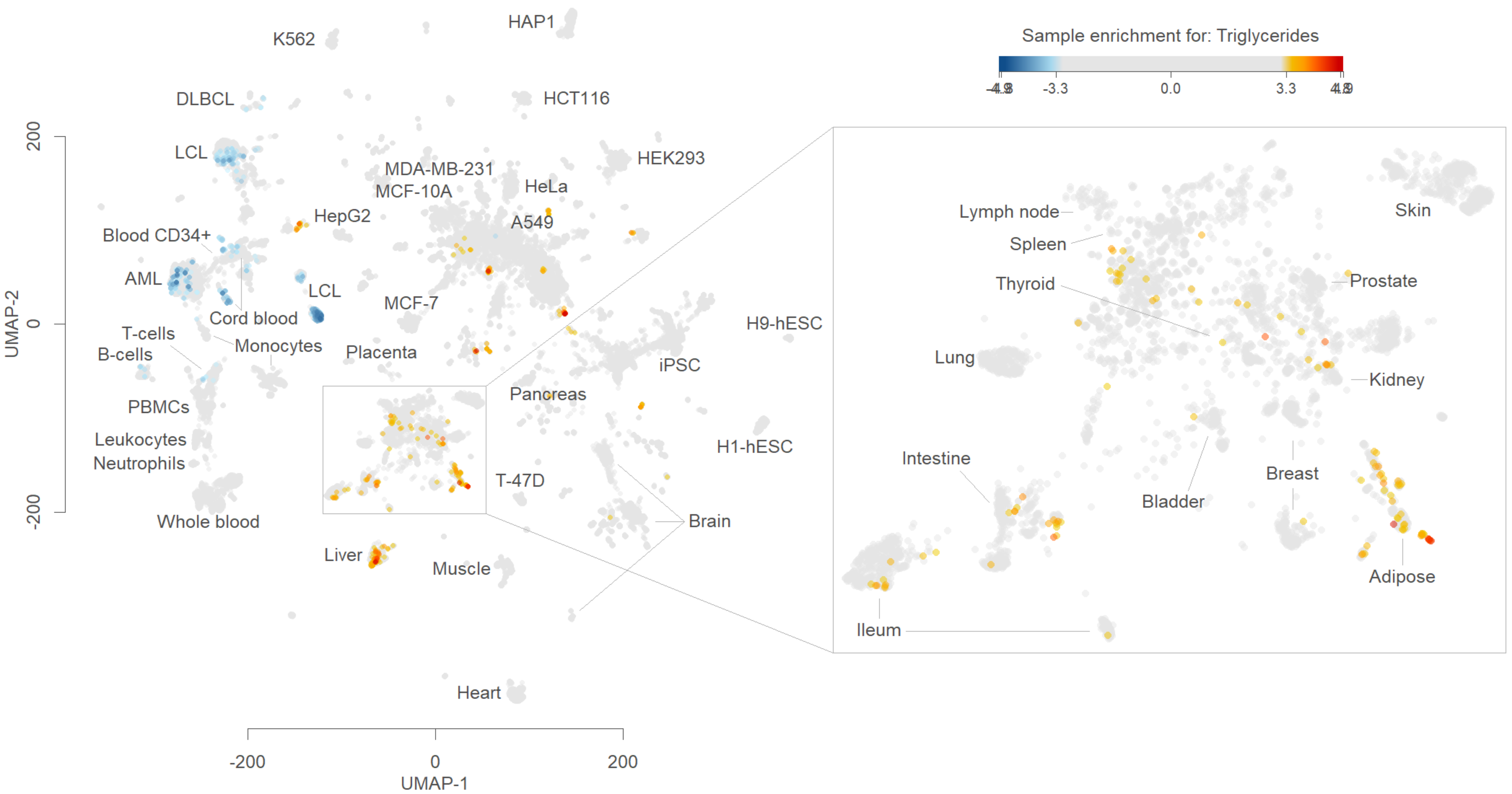

Sample enrichment for: Triglycerides
-4.8
-3.3
0.0
3.3
4.8
K562
HAP1
DLBCL
HCT116
LCL
HEK293
MDA-MB-231
MCF-10A
HeLa
HepG2
A549
Blood CD34+
AML
LCL
Cord blood
MCF-7
H9-hESC
T-cells
B-cells
Monocytes
Placenta
iPSC
Pancreas
PBMCs
Leukocytes
Neutrophils
H1-hESC
T-47D
Brain
Whole blood
Liver
Muscle
Heart
UMAP-2
UMAP-1
200
0
-200
Lymph node
Spleen
Thyroid
Skin
Prostate
Lung
Kidney
Intestine
Breast
Bladder
Adipose
Ileum

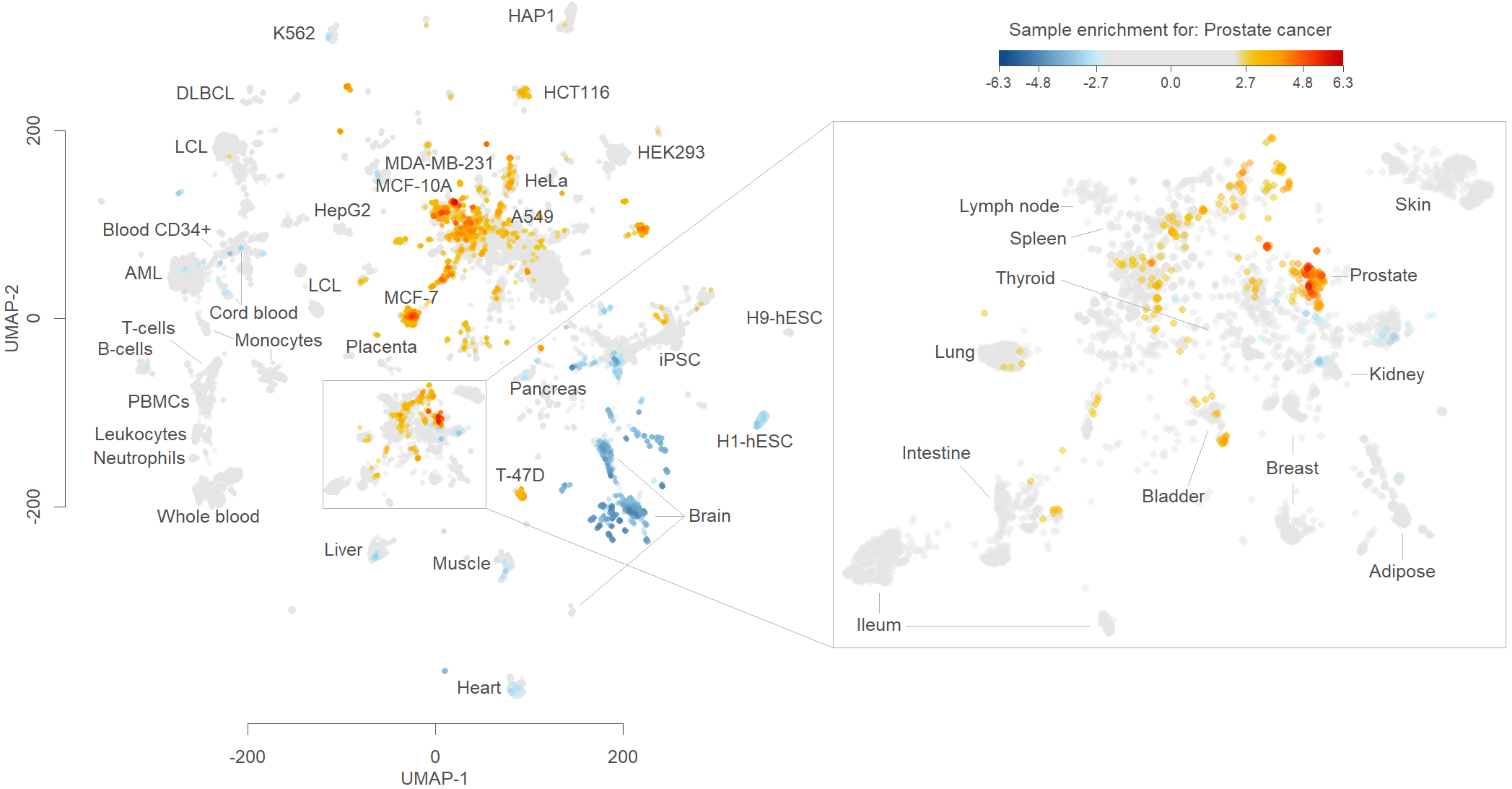

Sample enrichment for: Prostate cancer
-6.3
-4.8
-2.7
0.0
2.7
4.8
6.3
UMAP-2
UMAP-1
200
0
-200
HAP1
K562
DLBCL
HCT116
LCL
HEK293
MDA-MB-231
MCF-10A
HeLa
HepG2
A549
Blood CD34+
AML
LCL
MCF-7
Cord blood
T-cells
B-cells
Monocytes
Placenta
H9-hESC
iPSC
Pancreas
PBMCs
Leukocytes
Neutrophils
H1-hESC
T-47D
Brain
Whole blood
Liver
Muscle
Heart
Lymph node
Spleen
Thyroid
Lung
Skin
Prostate
Kidney
Intestine
Bladder
Breast
Adipose
Ileum

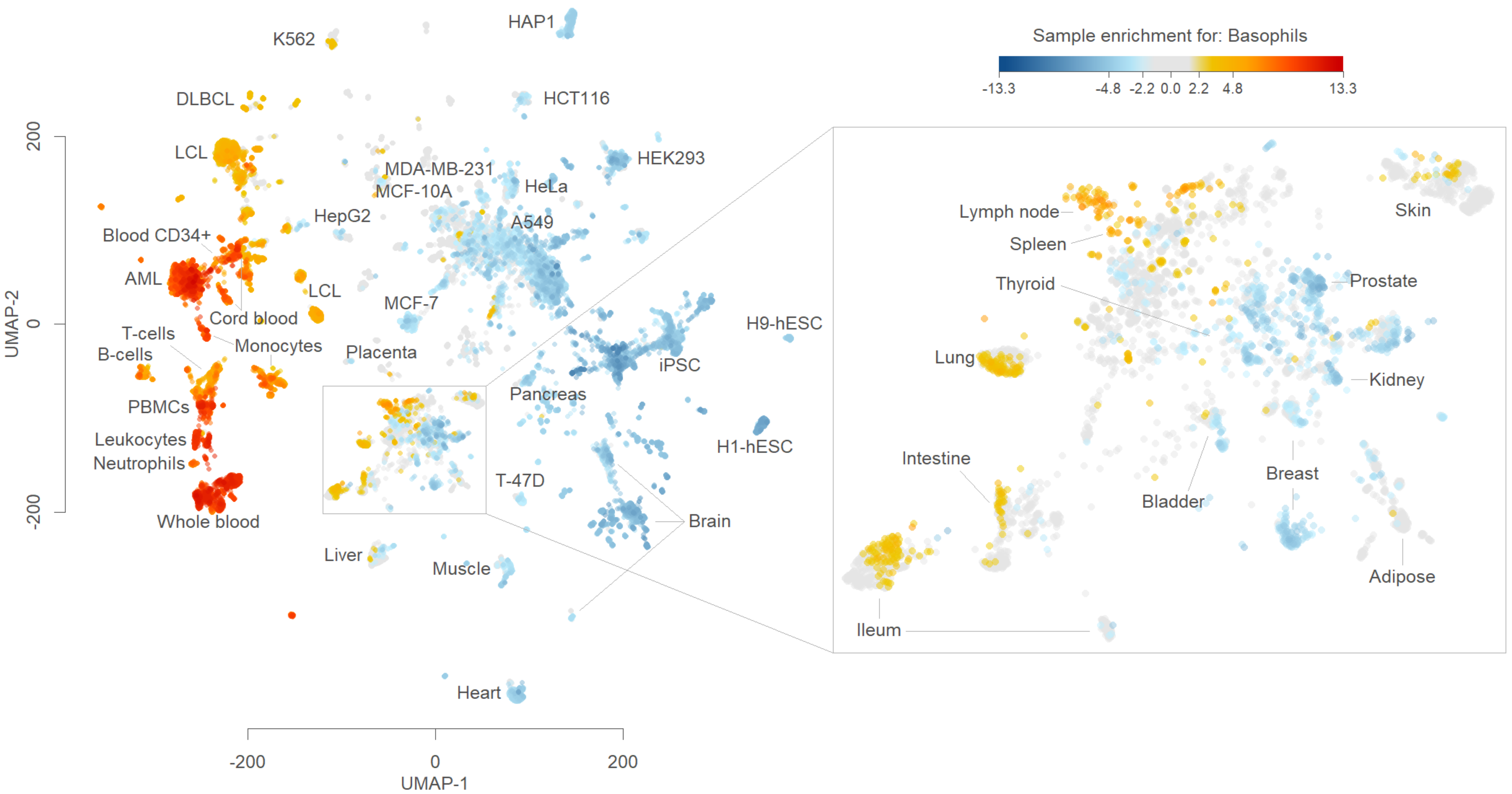

Sample enrichment for: Basophils
-13.3
-4.8
-2.2
0.0
2.2
4.8
13.3
K562
HAP1
DLBCL
HCT116
LCL
HEK293
MDA-MB-231
MCF-10A
HeLa
HepG2
A549
Blood CD34+
AML
LCL
MCF-7
Cord blood
T-cells
B-cells
Monocytes
Placenta
iPSC
H9-hESC
PBMCs
Pancreas
Leukocytes
Neutrophils
H1-hESC
T-47D
Whole blood
Brain
Liver
Muscle
Heart
UMAP-2
200
0
-200
UMAP-1
-200
0
200
Lymph node
Spleen
Thyroid
Skin
Prostate
Lung
Kidney
Intestine
Breast
Bladder
Adipose
Ileum

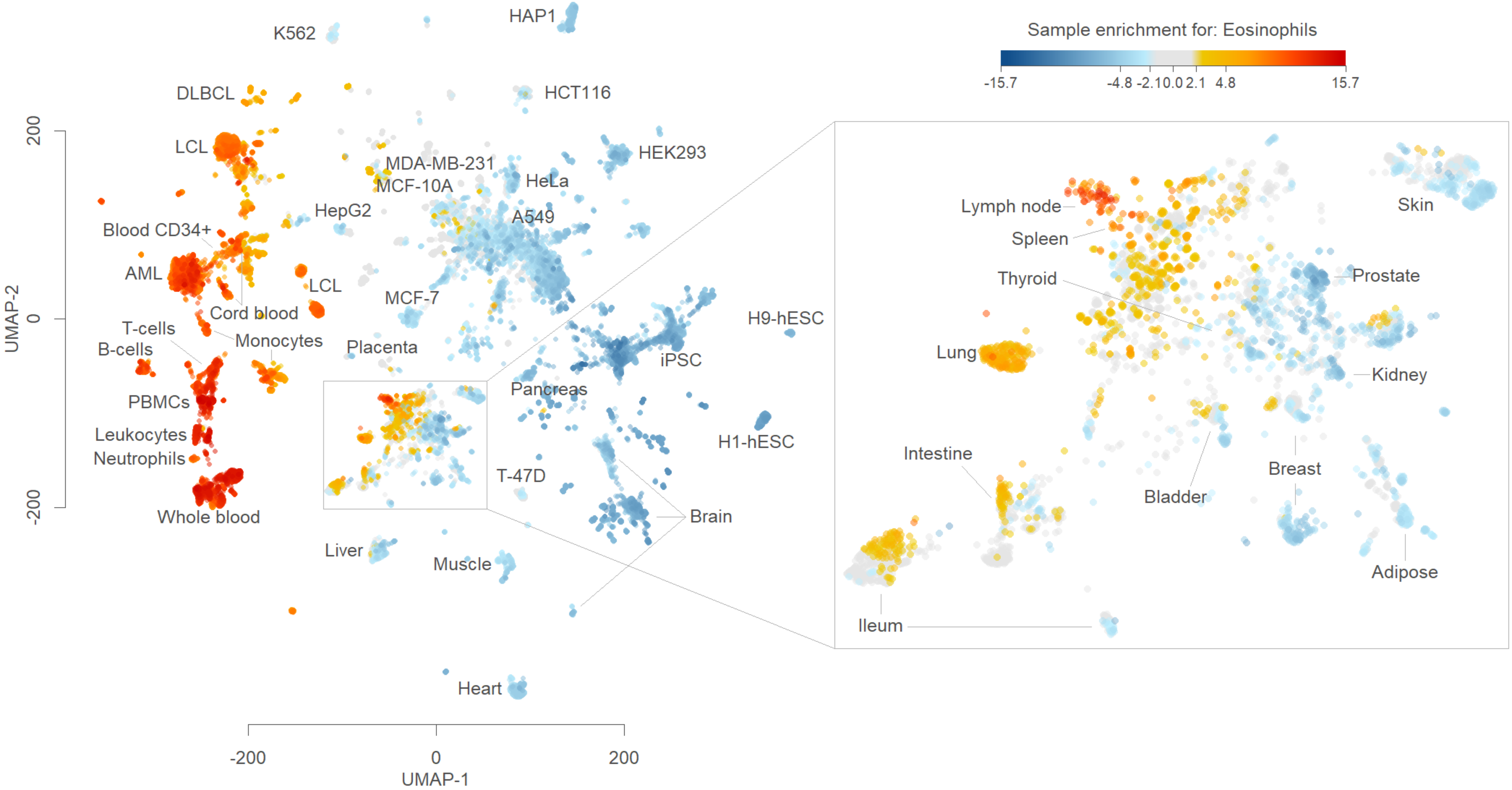

Sample enrichment for: Eosinophils
-15.7
-4.8
-2.1
0.0
2.1
4.8
15.7
K562
HAP1
DLBCL
HCT116
LCL
HEK293
MDA-MB-231
MCF-10A
HeLa
HepG2
A549
Blood CD34+
AML
LCL
Cord blood
MCF-7
T-cells
B-cells
Monocytes
Placenta
iPSC
H9-hESC
PBMCs
Pancreas
Leukocytes
Neutrophils
H1-hESC
T-47D
Whole blood
Brain
Liver
Muscle
Heart
UMAP-1
UMAP-2
200
0
-200
Lymph node
Spleen
Thyroid
Lung
Skin
Prostate
Kidney
Intestine
Bladder
Breast
Adipose
Ileum

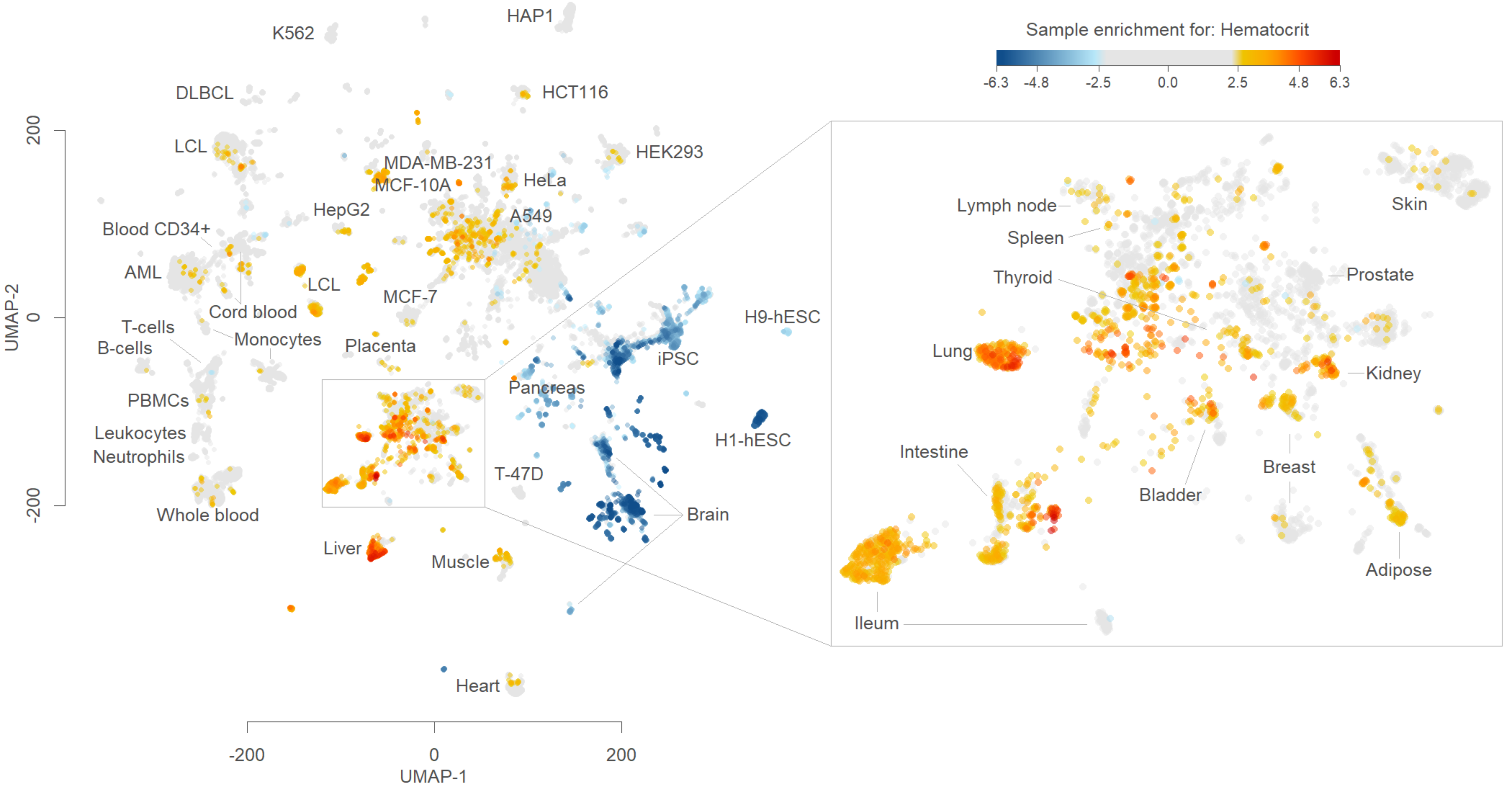

Sample enrichment for: Hematocrit
-6.3
-4.8
-2.5
0.0
2.5
4.8
6.3
HAP1
K562
DLBCL
HCT116
LCL
HEK293
MDA-MB-231
MCF-10A
HeLa
HepG2
A549
Blood CD34+
AML
LCL
Cord blood
MCF-7
T-cells
B-cells
Monocytes
Placenta
H9-hESC
iPSC
PBMCs
Pancreas
Leukocytes
Neutrophils
H1-hESC
T-47D
Brain
Whole blood
Liver
Muscle
Heart
Lymph node
Spleen
Thyroid
Skin
Prostate
Lung
Kidney
Intestine
Bladder
Breast
Adipose
Ileum
UMAP-2
200
0
-200
UMAP-1
-200
0
200

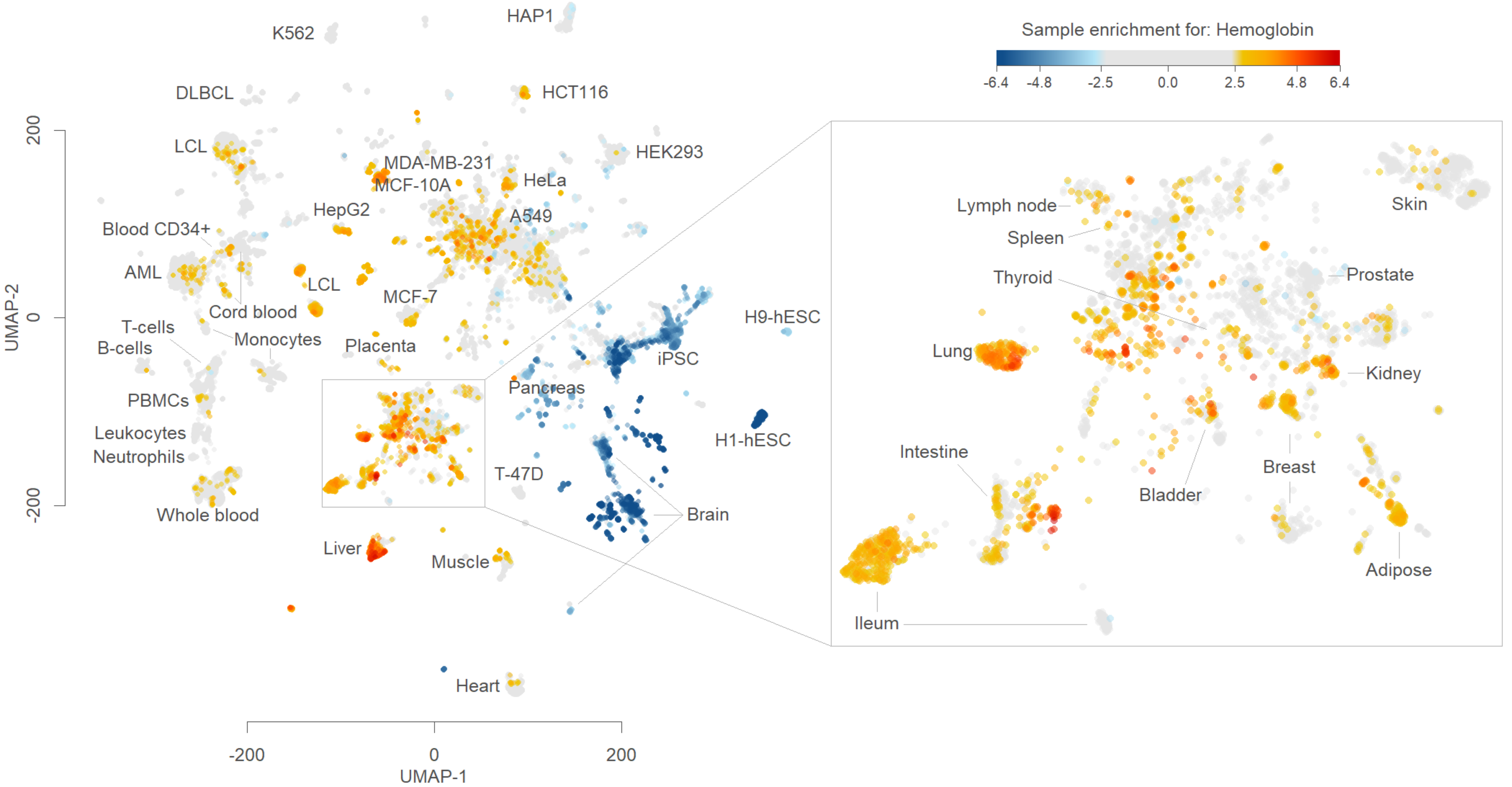
Sample enrichment for: Hemoglobin
-6.4
-4.8
-2.5
0.0
2.5
4.8
6.4
HAP1
K562
DLBCL
HCT116
LCL
HEK293
MDA-MB-231
MCF-10A
HeLa
HepG2
A549
Blood CD34+
AML
LCL
Cord blood
MCF-7
T-cells
B-cells
Monocytes
Placenta
iPSC
H9-hESC
PBMCs
Pancreas
Leukocytes
Neutrophils
H1-hESC
T-47D
Whole blood
Brain
Liver
Muscle
Heart
UMAP-2
200
0
-200
UMAP-1
-200
0
200
Lymph node
Spleen
Thyroid
Skin
Prostate
Lung
Kidney
Intestine
Bladder
Breast
Adipose
Ileum

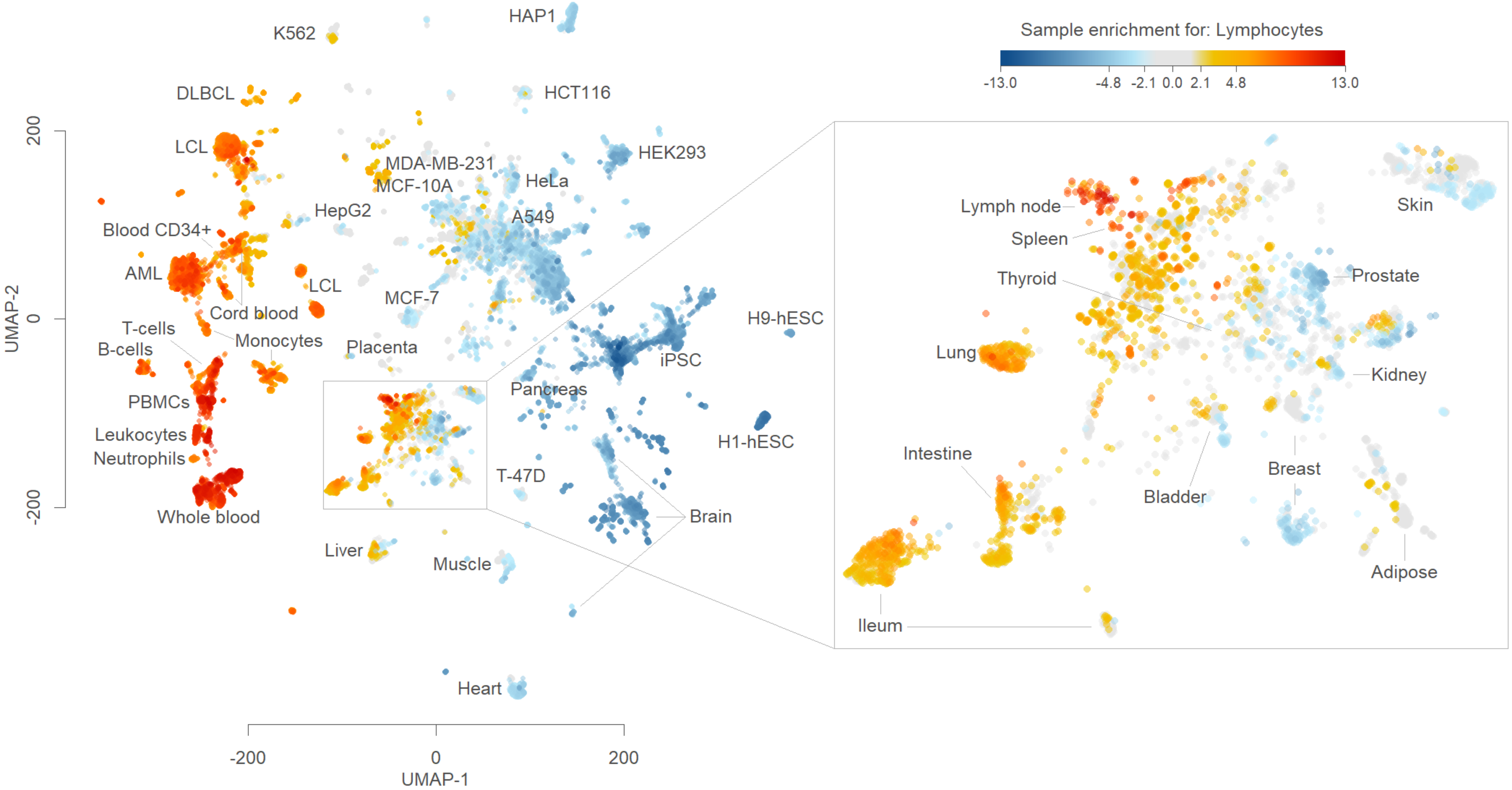
Sample enrichment for: Lymphocytes
-13.0
-4.8
-2.1
0.0
2.1
4.8
13.0
UMAP-2
UMAP-1
200
0
-200
K562
HAP1
DLBCL
HCT116
LCL
HEK293
MDA-MB-231
MCF-10A
HeLa
HepG2
A549
Blood CD34+
AML
LCL
Cord blood
MCF-7
H9-hESC
T-cells
B-cells
Monocytes
Placenta
iPSC
PBMCs
Pancreas
Leukocytes
Neutrophils
H1-hESC
T-47D
Brain
Whole blood
Liver
Muscle
Heart
Lymph node
Spleen
Thyroid
Lung
Skin
Prostate
Kidney
Intestine
Bladder
Breast
Adipose
Ileum

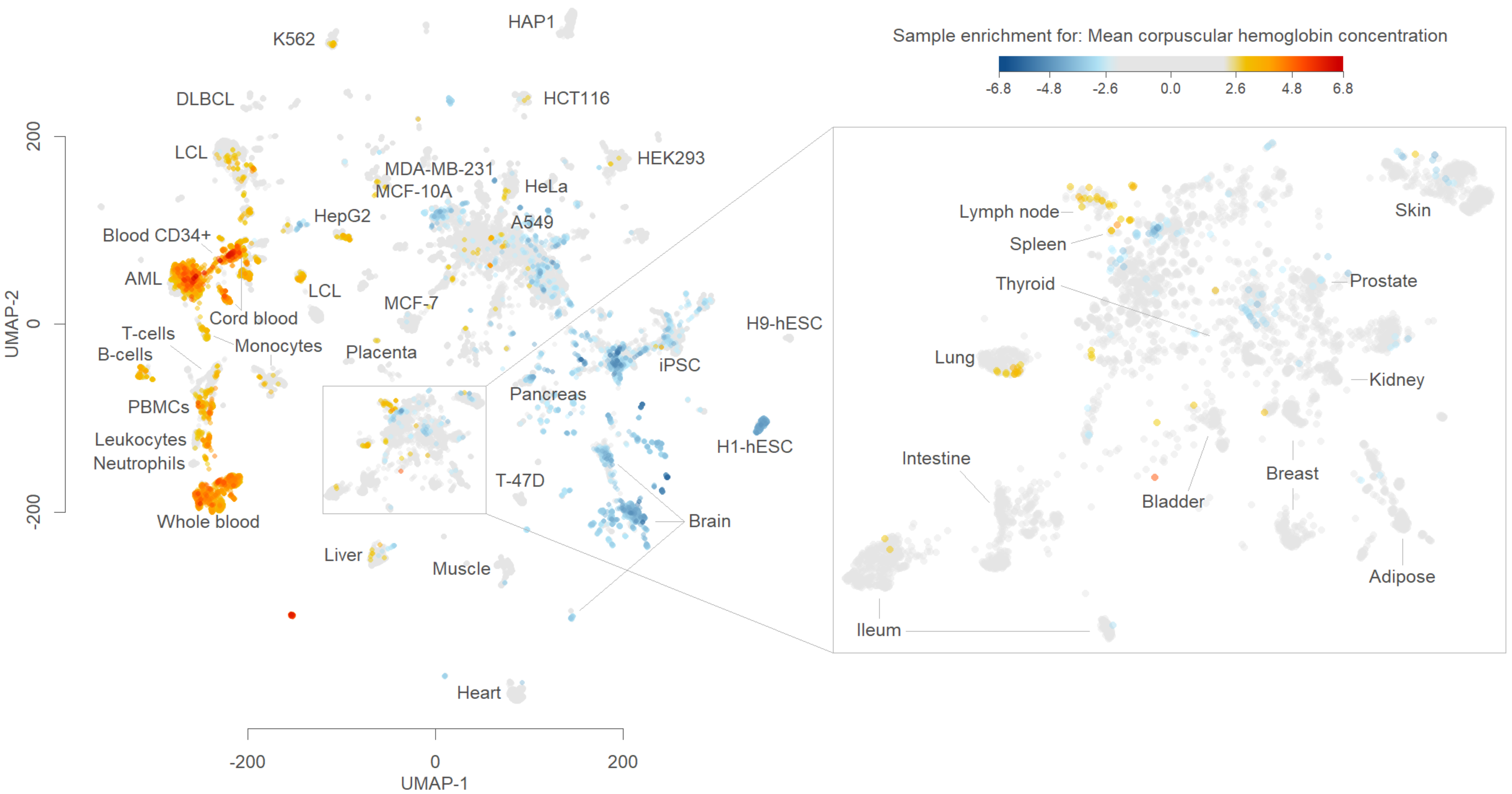

Sample enrichment for: Mean corpuscular hemoglobin concentration
-6.8
-4.8
-2.6
0.0
2.6
4.8
6.8
K562
HAP1
DLBCL
HCT116
LCL
MDA-MB-231
MCF-10A
HeLa
HEK293
HepG2
A549
Blood CD34+
AML
LCL
MCF-7
Cord blood
T-cells
B-cells
Monocytes
Placenta
H9-hESC
iPSC
Pancreas
PBMCs
Leukocytes
Neutrophils
H1-hESC
T-47D
Brain
Whole blood
Liver
Muscle
Heart
UMAP-2
UMAP-1
200
0
-200
Lymph node
Spleen
Thyroid
Skin
Prostate
Lung
Kidney
Intestine
Breast
Bladder
Adipose
Ileum

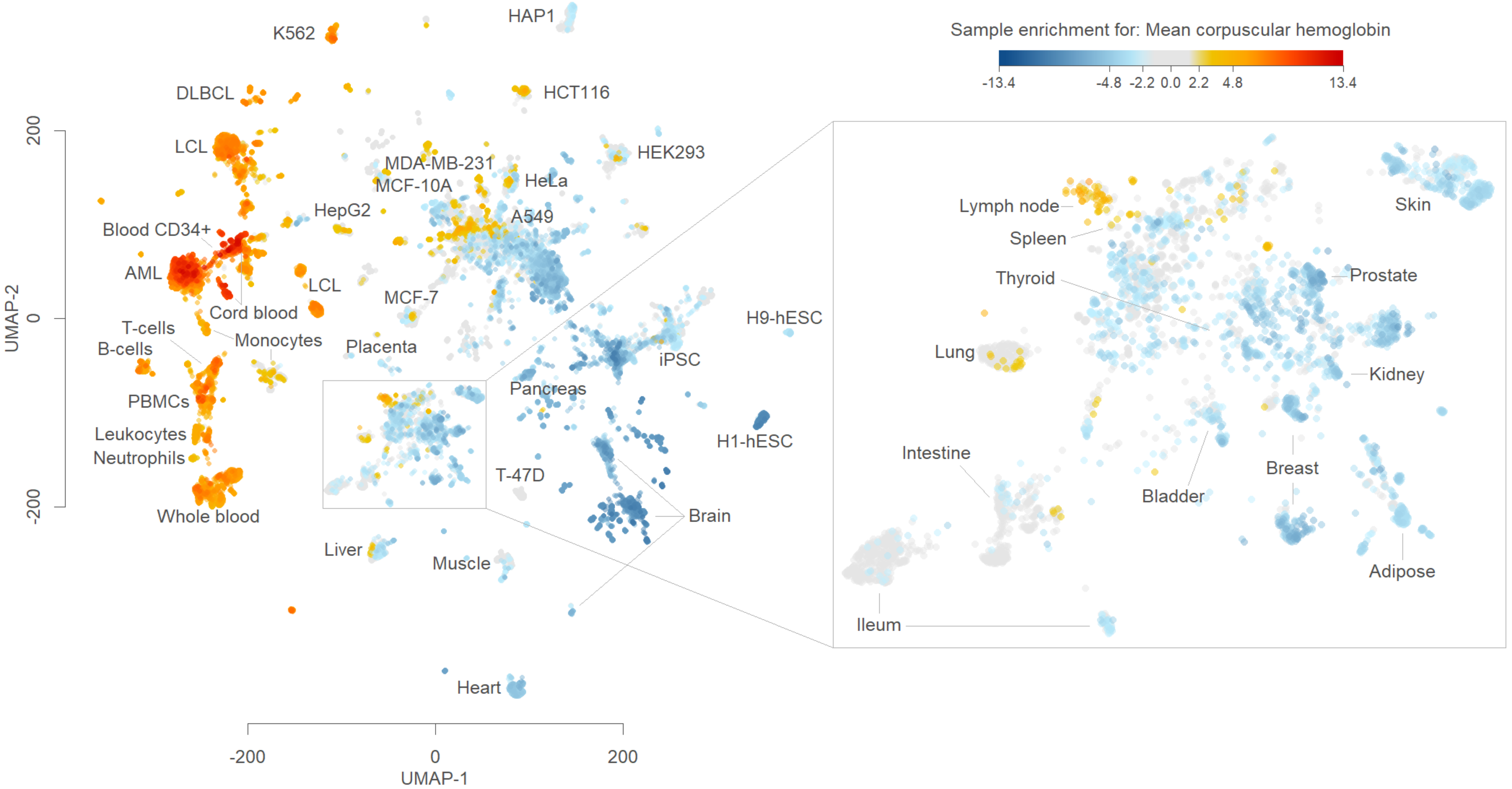
Sample enrichment for: Mean corpuscular hemoglobin
-13.4
-4.8
-2.2
0.0
2.2
4.8
13.4
HAP1
K562
DLBCL
HCT116
LCL
HEK293
MDA-MB-231
MCF-10A
HeLa
HepG2
A549
Blood CD34+
AML
LCL
MCF-7
Cord blood
T-cells
B-cells
Monocytes
Placenta
iPSC
H9-hESC
PBMCs
Pancreas
Leukocytes
Neutrophils
H1-hESC
T-47D
Brain
Whole blood
Liver
Muscle
Heart
UMAP-2
UMAP-1
200
0
-200
-200
0
200
Lymph node
Spleen
Thyroid
Lung
Skin
Prostate
Kidney
Intestine
Breast
Bladder
Adipose
Ileum

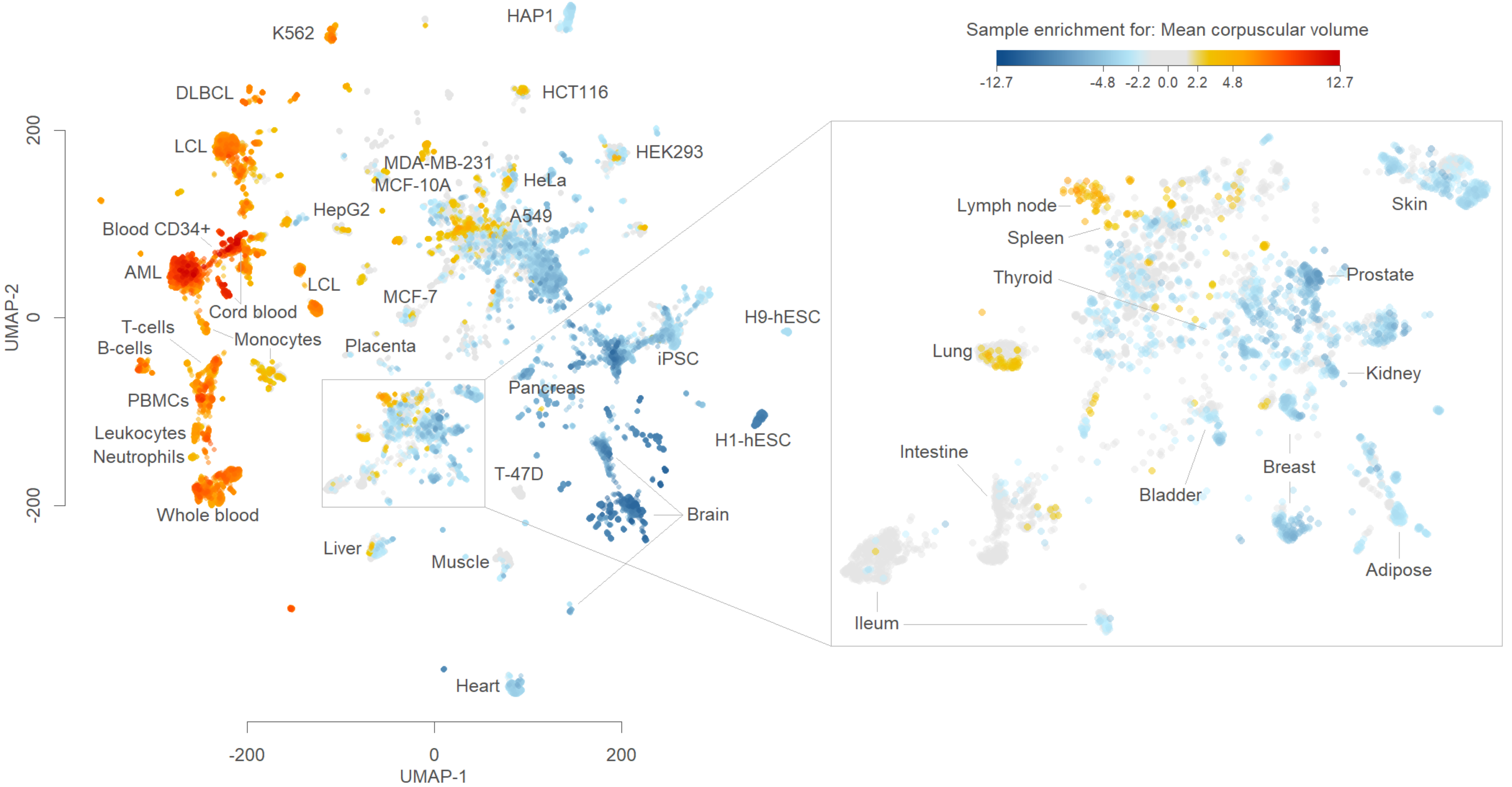

Sample enrichment for: Mean corpuscular volume
-12.7
-4.8
-2.2
0.0
2.2
4.8
12.7
UMAP-2
200
0
-200
UMAP-1
-200
0
200
HAP1
K562
DLBCL
HCT116
LCL
HEK293
MDA-MB-231
MCF-10A
HeLa
HepG2
A549
Blood CD34+
AML
LCL
MCF-7
Cord blood
T-cells
B-cells
Monocytes
Placenta
iPSC
H9-hESC
PBMCs
Pancreas
Leukocytes
Neutrophils
H1-hESC
T-47D
Whole blood
Brain
Liver
Muscle
Heart
Lymph node
Spleen
Thyroid
Skin
Prostate
Lung
Kidney
Intestine
Breast
Bladder
Adipose
Ileum

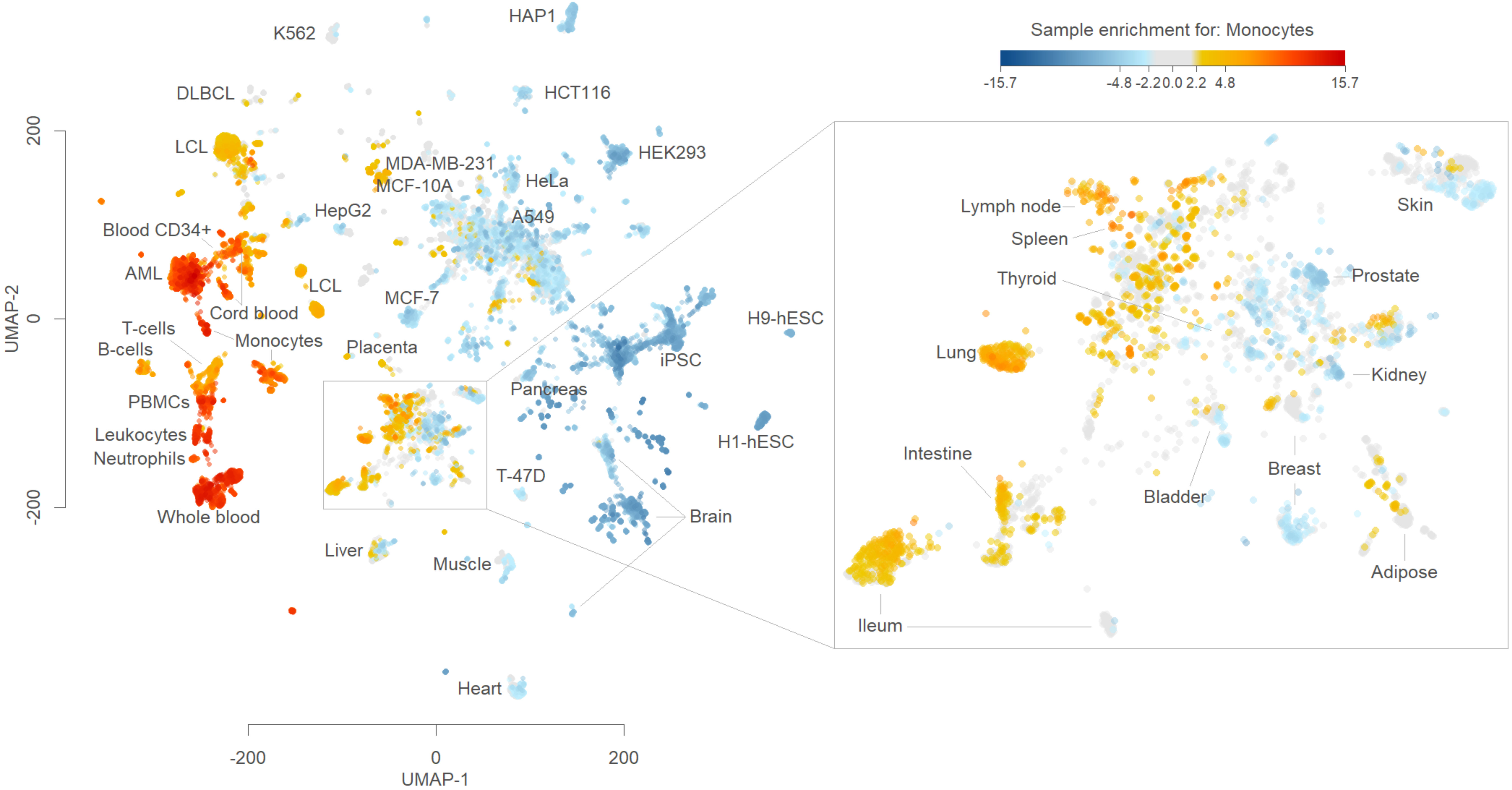
Sample enrichment for: Monocytes
-15.7
-4.8
-2.2
0.0
2.2
4.8
15.7
HAP1
K562
DLBCL
HCT116
LCL
HEK293
MDA-MB-231
MCF-10A
HeLa
HepG2
A549
Blood CD34+
AML
LCL
Cord blood
MCF-7
H9-hESC
T-cells
B-cells
Monocytes
Placenta
iPSC
PBMCs
Pancreas
Leukocytes
Neutrophils
H1-hESC
T-47D
Whole blood
Brain
Liver
Muscle
Heart
UMAP-1
UMAP-2
200
0
-200
Lymph node
Spleen
Thyroid
Skin
Prostate
Lung
Kidney
Intestine
Breast
Bladder
Adipose
Ileum

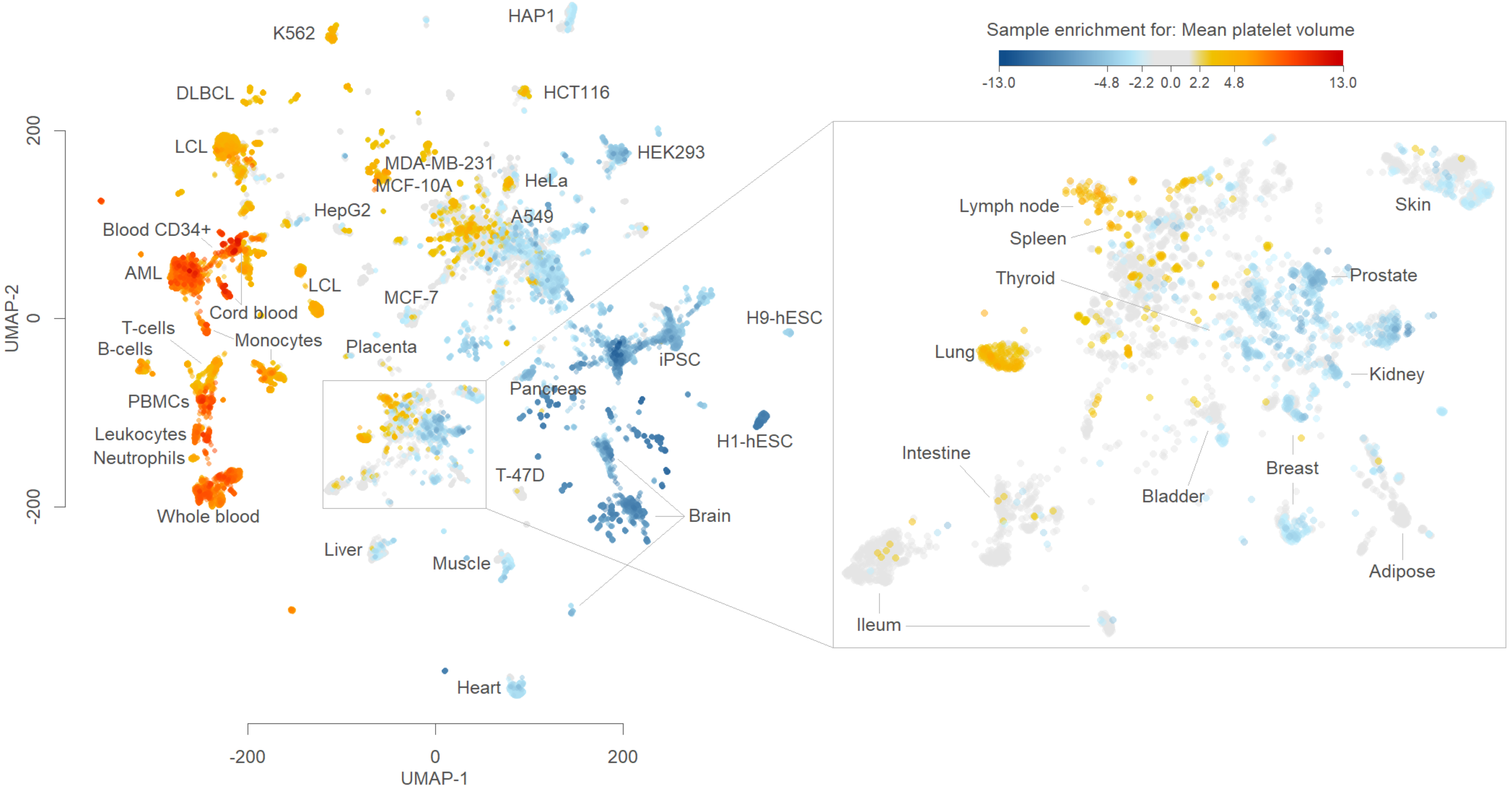

Sample enrichment for: Mean platelet volume
-13.0
-4.8
-2.2
0.0
2.2
4.8
13.0
HAP1
K562
DLBCL
HCT116
LCL
HEK293
MDA-MB-231
MCF-10A
HeLa
HepG2
A549
Blood CD34+
AML
LCL
MCF-7
Cord blood
T-cells
Monocytes
B-cells
Placenta
iPSC
H9-hESC
PBMCs
Pancreas
Leukocytes
H1-hESC
Neutrophils
T-47D
Whole blood
Brain
Liver
Muscle
Heart
Lymph node
Spleen
Thyroid
Skin
Prostate
Lung
Kidney
Intestine
Breast
Bladder
Adipose
Ileum
UMAP-2
200
0
-200
UMAP-1
-200
0
200

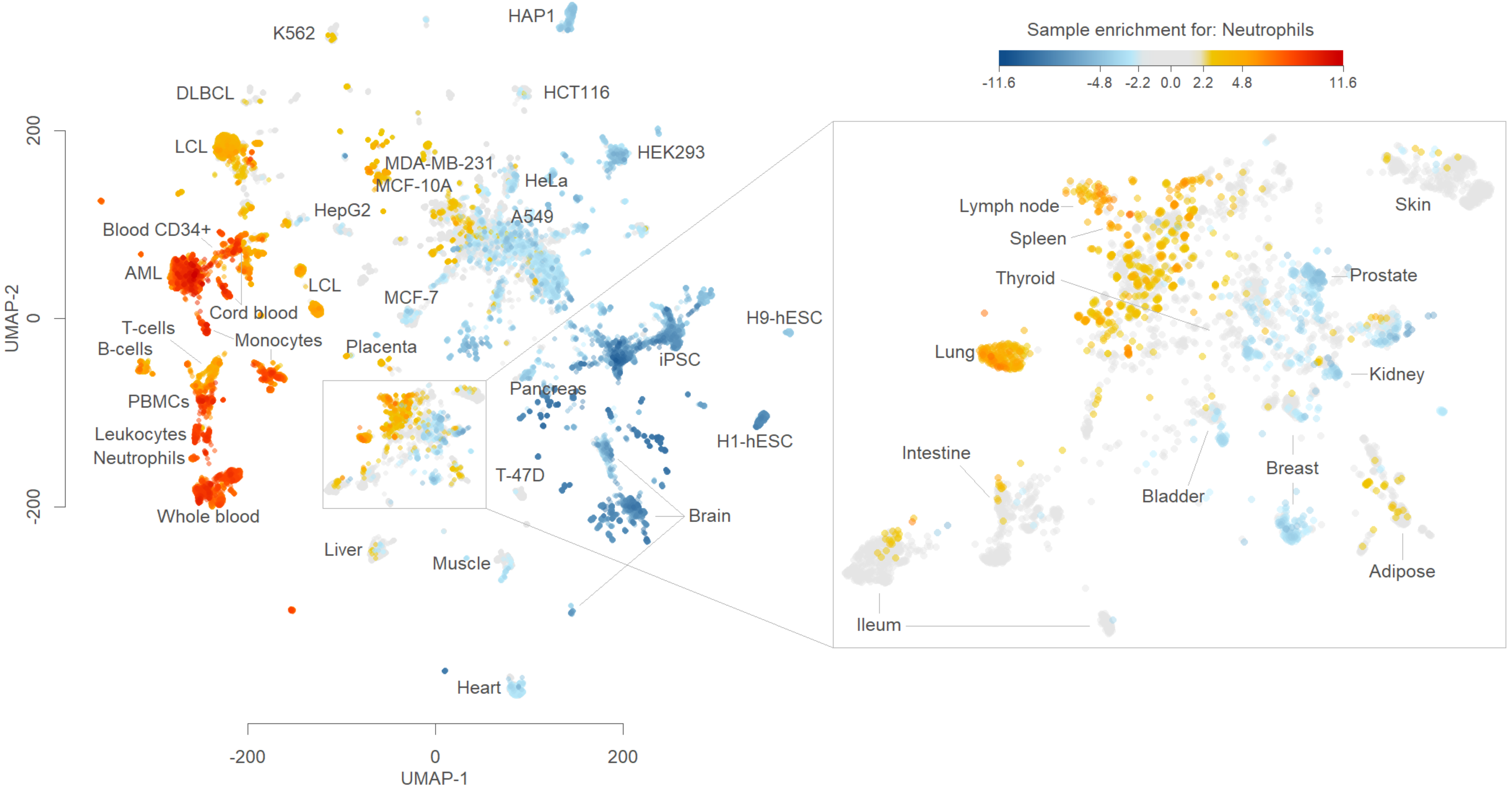
Sample enrichment for: Neutrophils
-11.6
-4.8
-2.2
0.0
2.2
4.8
11.6
UMAP-2
UMAP-1
200
0
-200
-200
0
200
HAP1
K562
DLBCL
HCT116
LCL
HEK293
MDA-MB-231
MCF-10A
HeLa
HepG2
A549
Blood CD34+
AML
LCL
MCF-7
Cord blood
T-cells
B-cells
Monocytes
Placenta
H9-hESC
iPSC
PBMCs
Pancreas
Leukocytes
Neutrophils
H1-hESC
T-47D
Whole blood
Brain
Liver
Muscle
Heart
Lymph node
Spleen
Thyroid
Skin
Prostate
Lung
Kidney
Intestine
Breast
Bladder
Adipose
Ileum

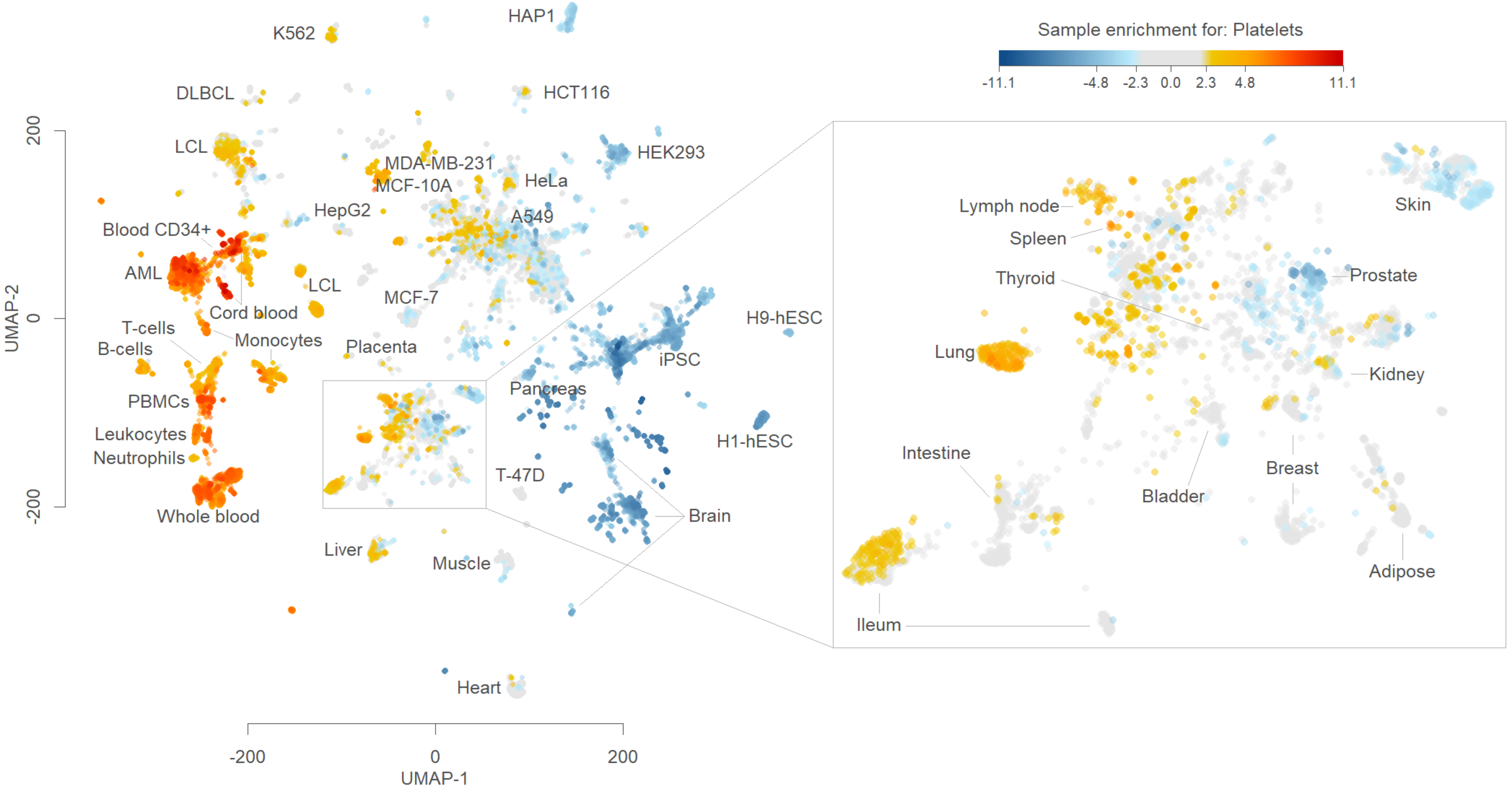

Sample enrichment for: Platelets
-11.1
-4.8
-2.3
0.0
2.3
4.8
11.1
HAP1
K562
DLBCL
HCT116
LCL
MDA-MB-231
HEK293
MCF-10A
HeLa
HepG2
A549
Blood CD34+
AML
LCL
MCF-7
Cord blood
T-cells
Monocytes
B-cells
Placenta
iPSC
H9-hESC
Pancreas
PBMCs
Leukocytes
Neutrophils
T-47D
H1-hESC
Whole blood
Brain
Liver
Muscle
Heart
UMAP-1
UMAP-2
200
0
-200
-200
0
200
Lymph node
Spleen
Thyroid
Skin
Prostate
Lung
Kidney
Intestine
Bladder
Breast
Adipose
Ileum

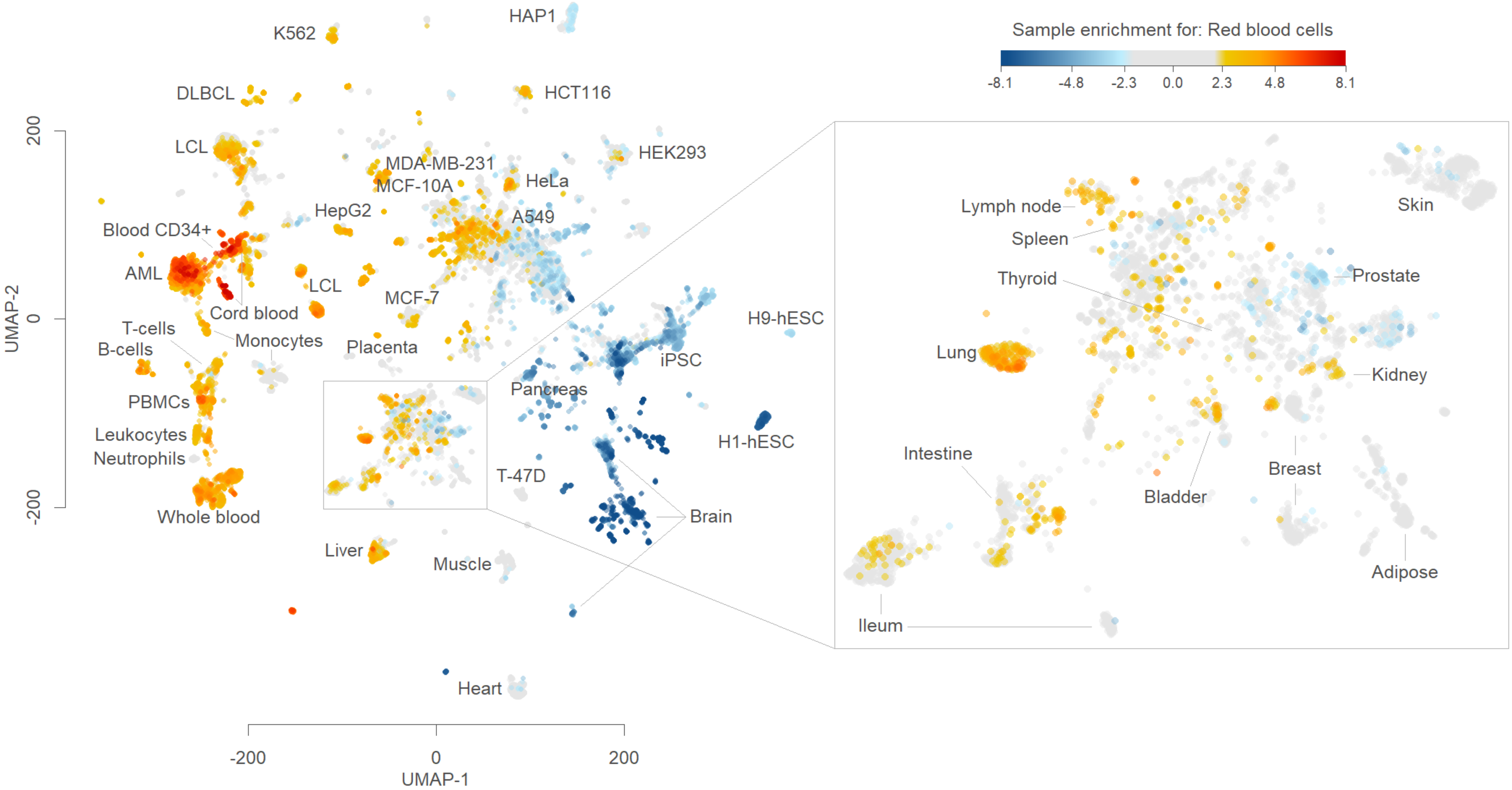

Sample enrichment for: Red blood cells
-8.1
-4.8
-2.3
0.0
2.3
4.8
8.1
UMAP-2
UMAP-1
200
0
-200
-200
0
200
K562
HAP1
DLBCL
HCT116
LCL
HEK293
MDA-MB-231
MCF-10A
HeLa
HepG2
A549
Blood CD34+
AML
LCL
MCF-7
Cord blood
T-cells
B-cells
Monocytes
Placenta
iPSC
H9-hESC
Pancreas
PBMCs
Leukocytes
Neutrophils
H1-hESC
T-47D
Whole blood
Brain
Liver
Muscle
Heart
Lymph node
Spleen
Thyroid
Skin
Prostate
Lung
Kidney
Intestine
Bladder
Breast
Ileum
Adipose

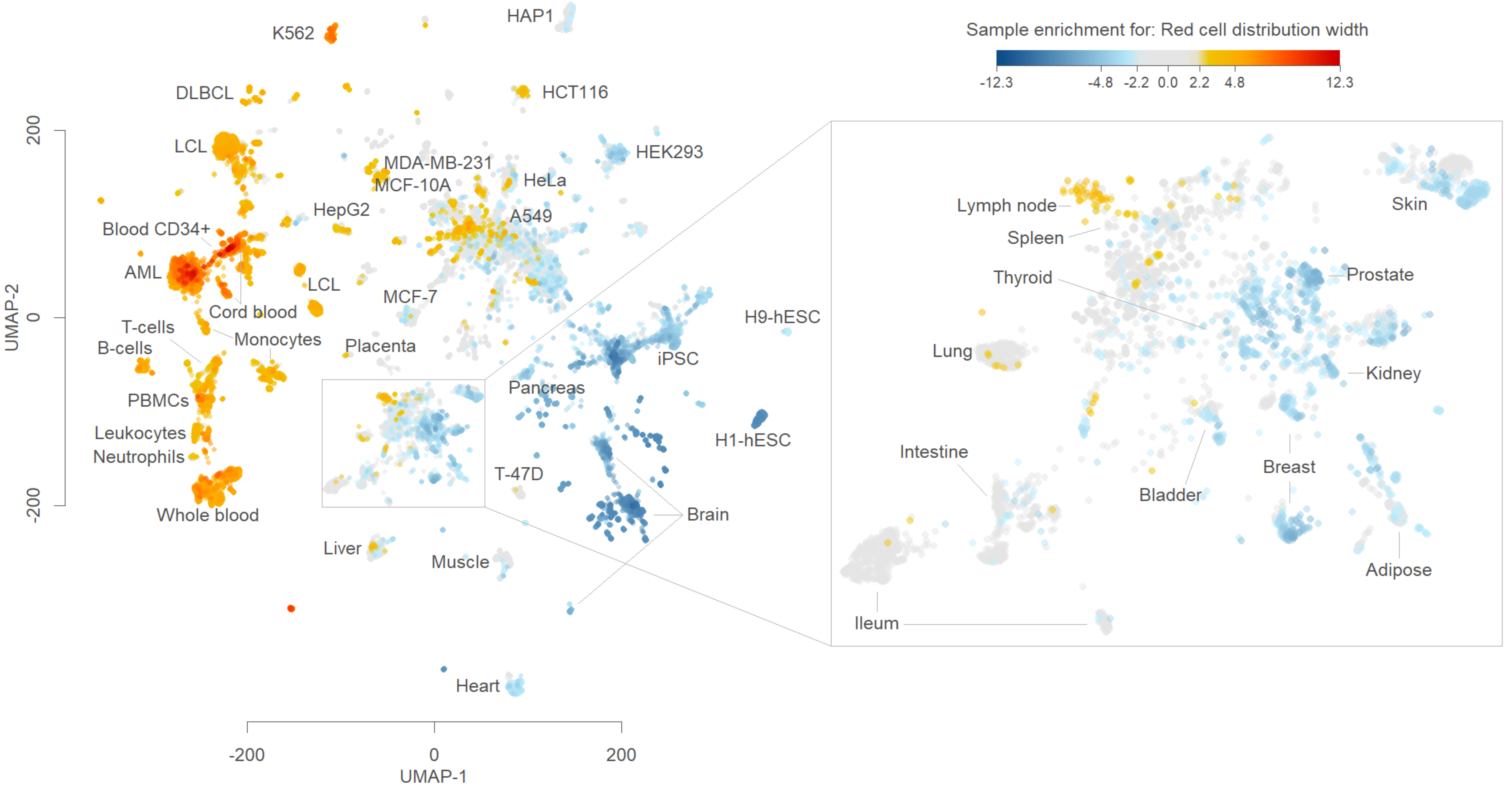
Sample enrichment for: Red cell distribution width
-12.3
-4.8
-2.2
0.0
2.2
4.8
12.3
UMAP-2
UMAP-1
200
0
-200
HAP1
K562
DLBCL
HCT116
LCL
HEK293
MDA-MB-231
MCF-10A
HeLa
HepG2
A549
Blood CD34+
AML
LCL
MCF-7
Cord blood
T-cells
Monocytes
B-cells
Placenta
iPSC
H9-hESC
PBMCs
Pancreas
Leukocytes
H1-hESC
Neutrophils
T-47D
Whole blood
Brain
Liver
Muscle
Heart
Lymph node
Spleen
Thyroid
Skin
Prostate
Lung
Kidney
Intestine
Breast
Bladder
Adipose
Ileum

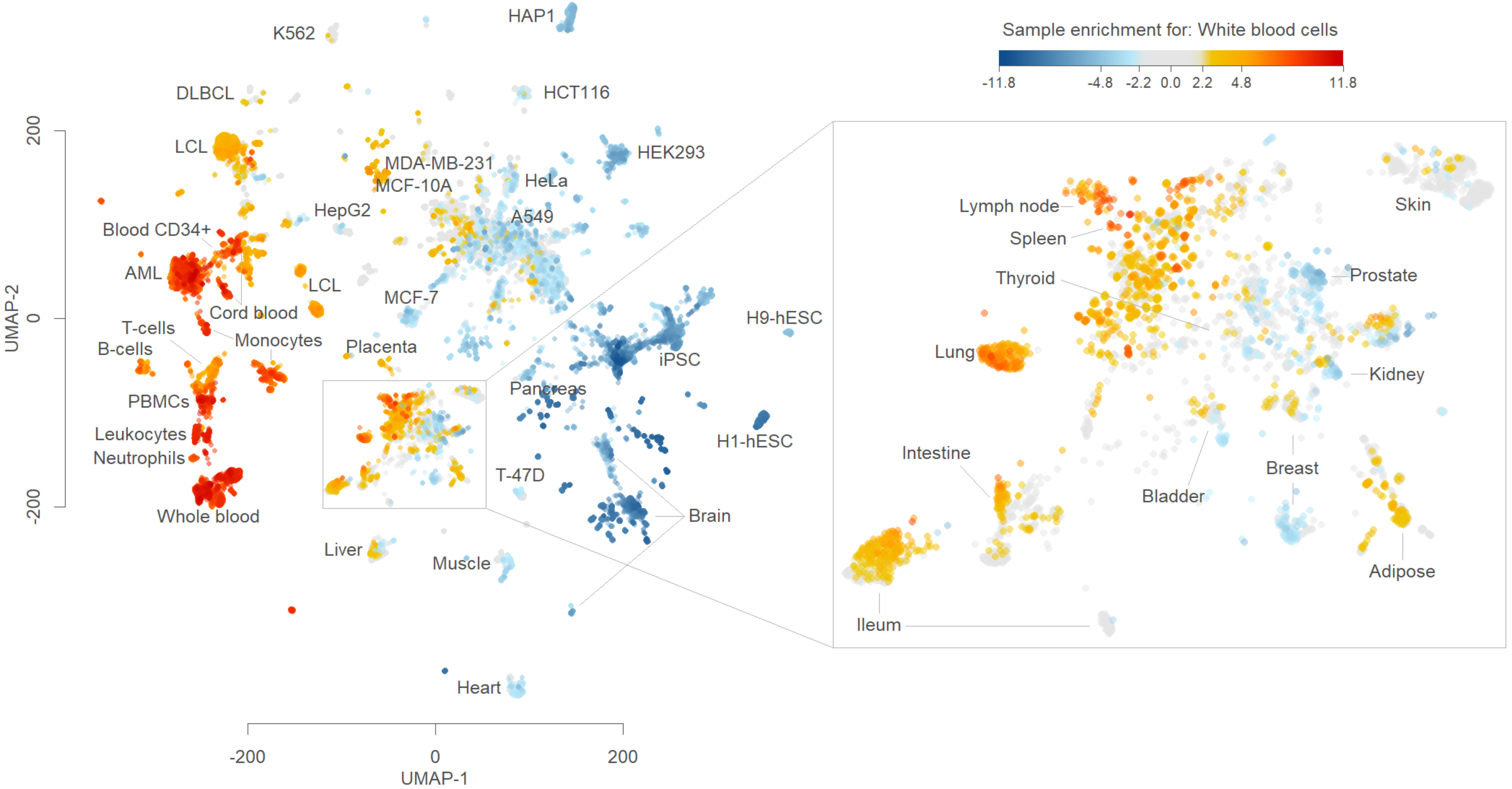

Sample enrichment for: White blood cells
-11.8
-4.8
-2.2
0.0
2.2
4.8
11.8
HAP1
K562
DLBCL
HCT116
LCL
HEK293
MDA-MB-231
MCF-10A
HeLa
HepG2
A549
Blood CD34+
AML
LCL
MCF-7
Cord blood
T-cells
B-cells
Monocytes
Placenta
iPSC
H9-hESC
PBMCs
Pancreas
Leukocytes
Neutrophils
H1-hESC
T-47D
Brain
Whole blood
Liver
Muscle
Heart
UMAP-1
UMAP-2
-200
0
200
Lymph node
Spleen
Thyroid
Skin
Prostate
Lung
Kidney
Intestine
Bladder
Breast
Adipose
Ileum

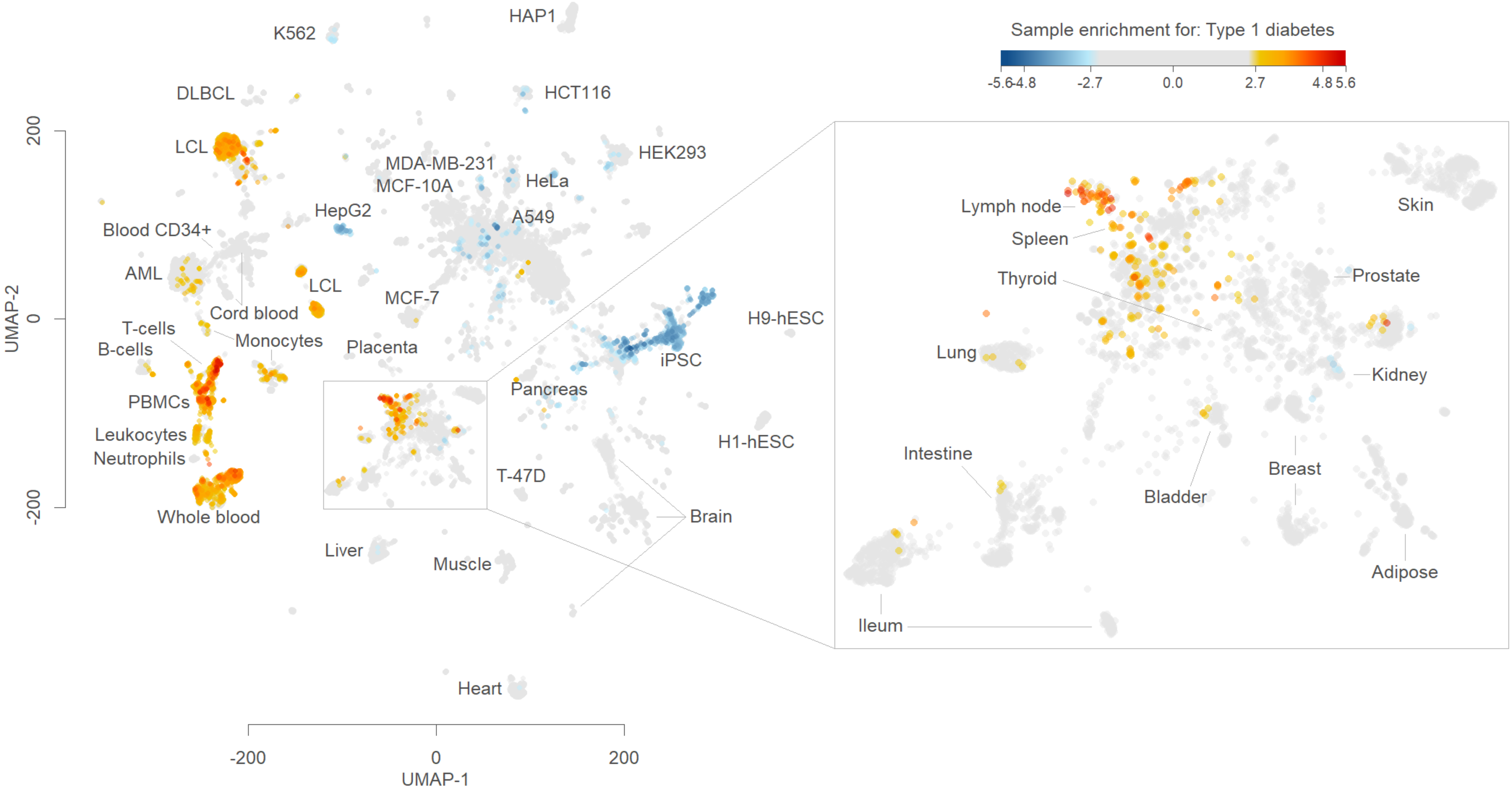

Sample enrichment for: Type 1 diabetes
-5.6 -4.8 -2.7 0.0 2.7 4.8 5.6
HAP1
K562
DLBCL
HCT116
LCL
HEK293
MDA-MB-231
MCF-10A
HeLa
HepG2
A549
Blood CD34+
AML
LCL
MCF-7
Cord blood
T-cells
B-cells
Monocytes
Placenta
iPSC
H9-hESC
PBMCs
Leukocytes
Neutrophils
Pancreas
H1-hESC
T-47D
Whole blood
Brain
Liver
Muscle
Heart
Lymph node
Spleen
Thyroid
Lung
Skin
Prostate
Kidney
Intestine
Bladder
Breast
Adipose
Ileum
UMAP-2
200
0
-200
UMAP-1
-200
0
200

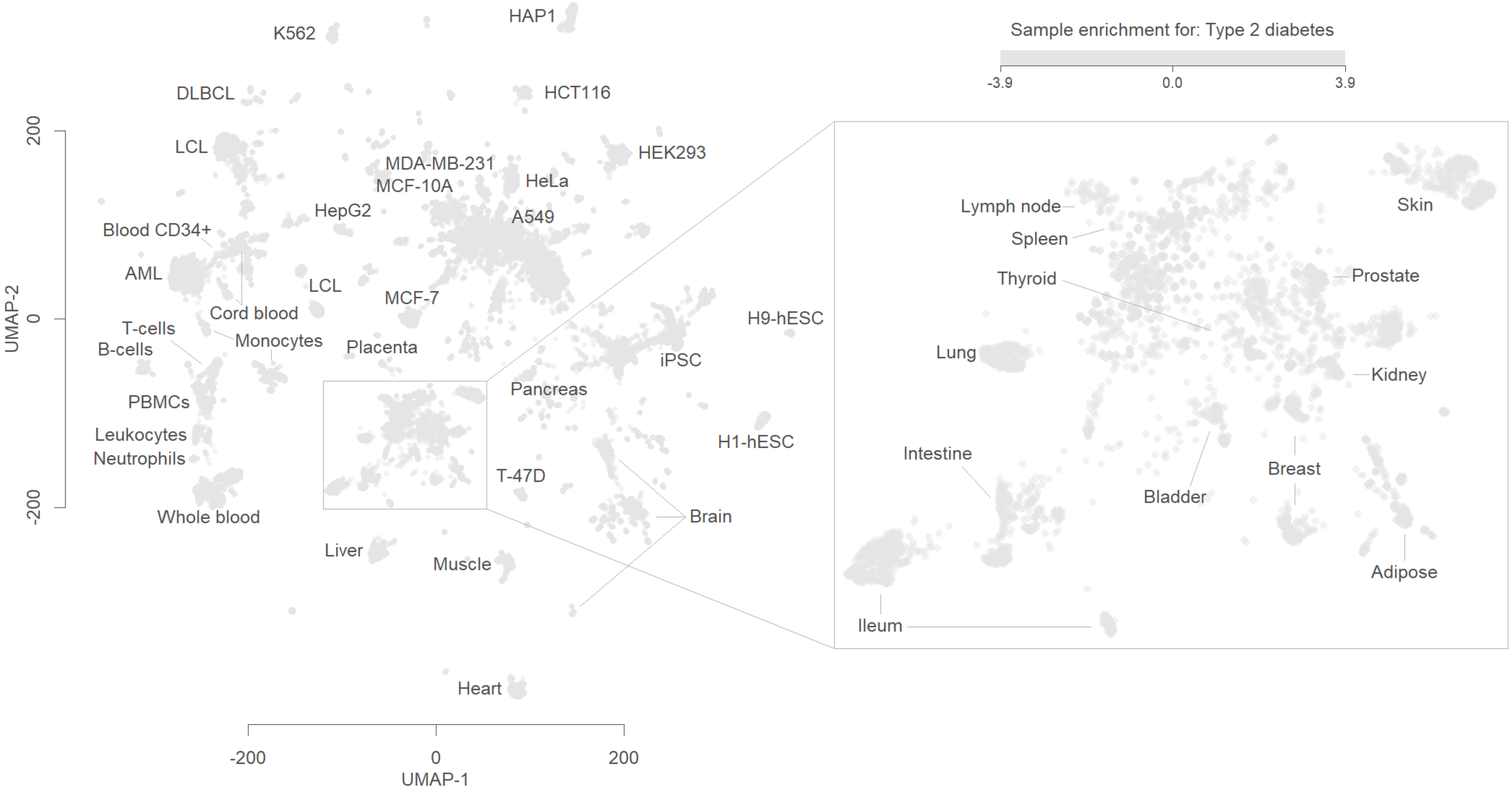
Sample enrichment for: Type 2 diabetes
-3.9
0.0
3.9
UMAP-1
UMAP-2
-200
0
200
HAP1
K562
DLBCL
HCT116
LCL
HEK293
MDA-MB-231
MCF-10A
HeLa
HepG2
A549
Blood CD34+
AML
LCL
MCF-7
Cord blood
H9-hESC
T-cells
B-cells
Monocytes
Placenta
iPSC
PBMCs
Pancreas
Leukocytes
Neutrophils
H1-hESC
T-47D
Whole blood
Brain
Liver
Muscle
Heart
Lymph node
Spleen
Thyroid
Skin
Prostate
Lung
Kidney
Intestine
Bladder
Breast
Adipose
Ileum

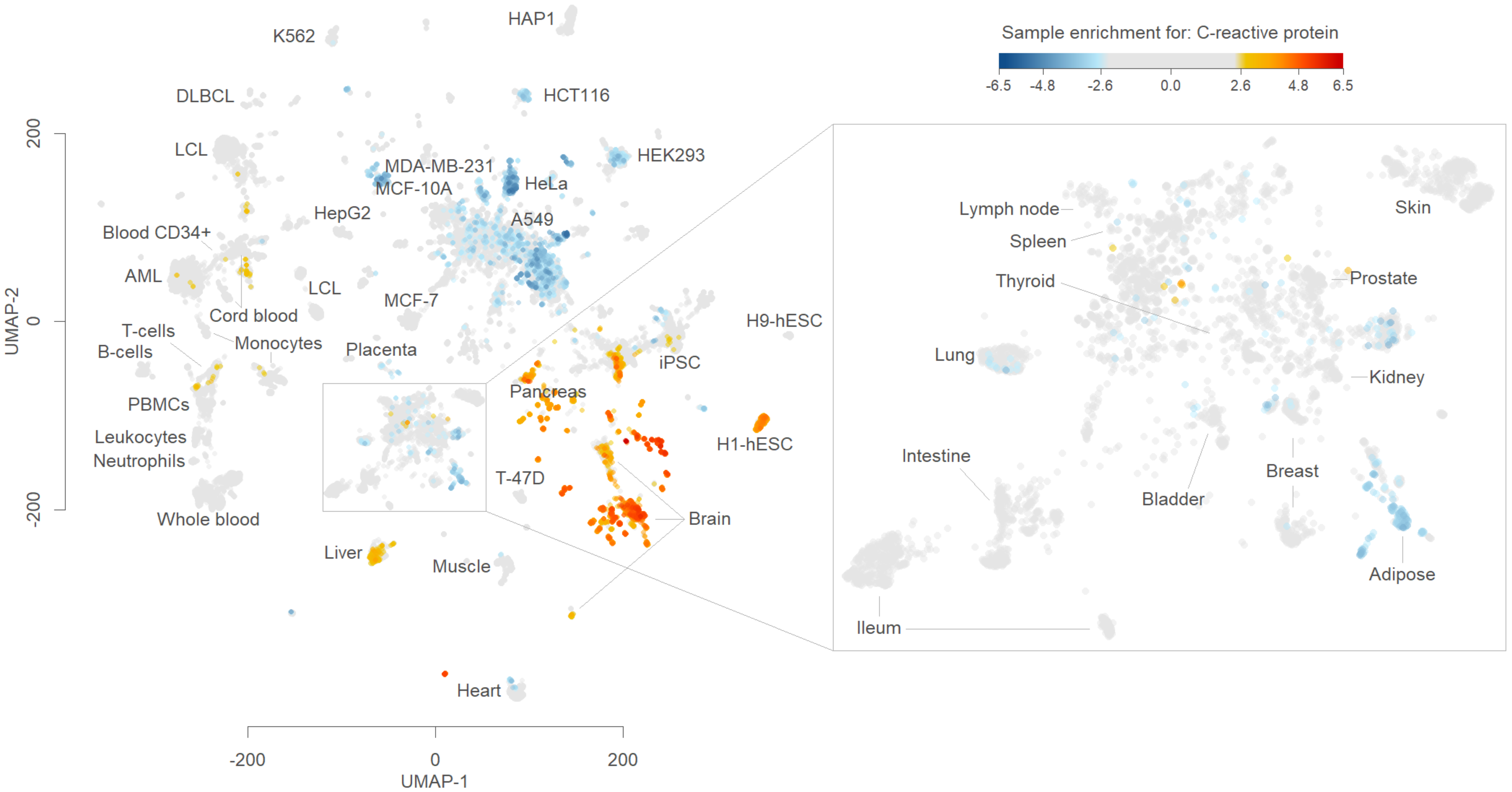
Sample enrichment for: C-reactive protein
-6.5
-4.8
-2.6
0.0
2.6
4.8
6.5
K562
HAP1
DLBCL
HCT116
LCL
HEK293
MDA-MB-231
MCF-10A
HeLa
HepG2
A549
Blood CD34+
AML
LCL
MCF-7
Cord blood
T-cells
B-cells
Monocytes
Placenta
H9-hESC
iPSC
PBMCs
Pancreas
Leukocytes
Neutrophils
H1-hESC
T-47D
Brain
Whole blood
Liver
Muscle
Heart
UMAP-1
UMAP-2
-200
0
200
Lymph node
Spleen
Thyroid
Skin
Prostate
Lung
Kidney
Intestine
Breast
Bladder
Adipose
Ileum

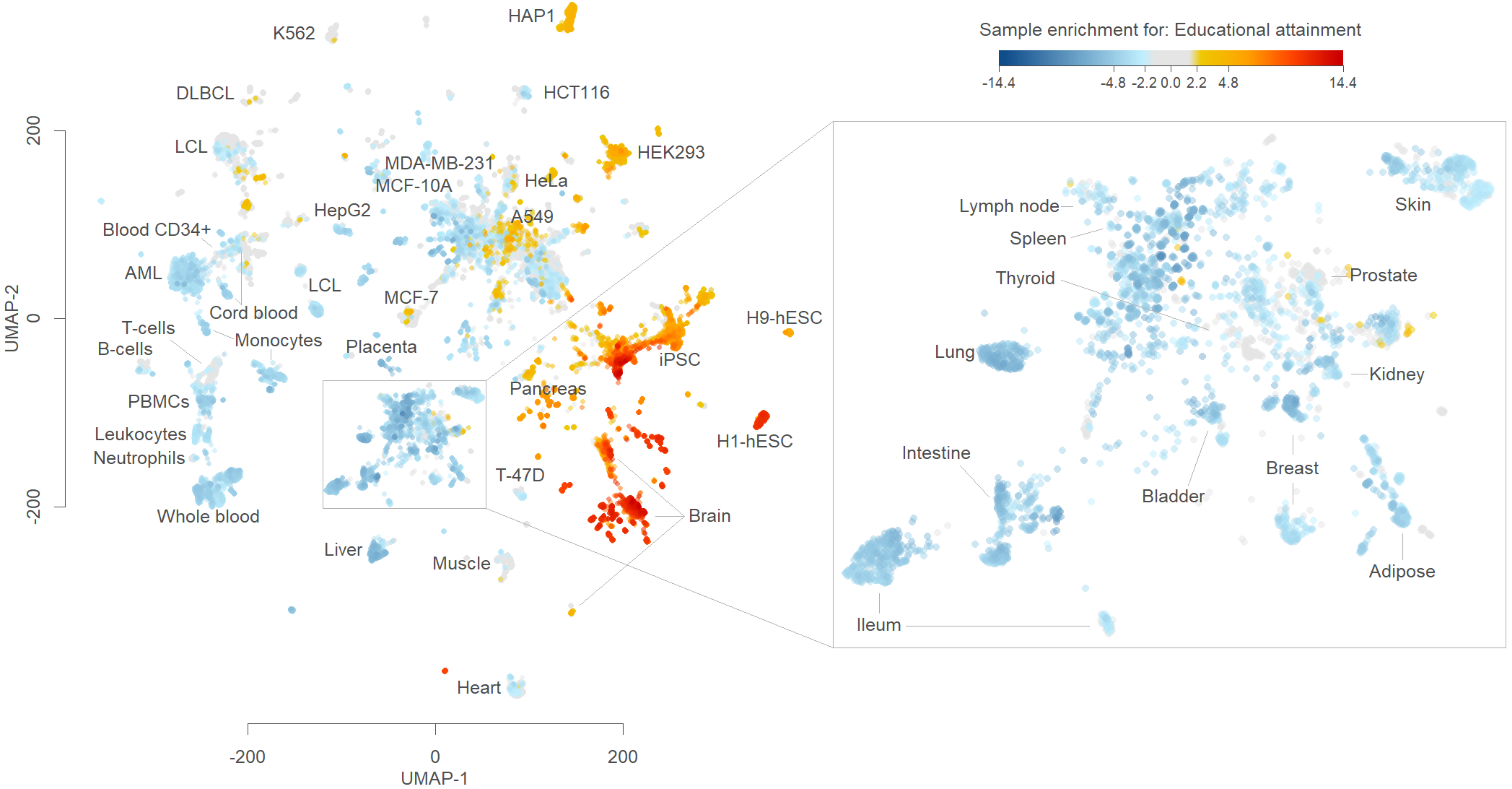
Sample enrichment for: Educational attainment
-14.4
-4.8
-2.2
0.0
2.2
4.8
14.4
HAP1
K562
DLBCL
HCT116
LCL
HEK293
MDA-MB-231
MCF-10A
HeLa
HepG2
A549
Blood CD34+
AML
LCL
Cord blood
MCF-7
T-cells
B-cells
Monocytes
Placenta
iPSC
H9-hESC
PBMCs
Pancreas
Leukocytes
Neutrophils
H1-hESC
T-47D
Whole blood
Brain
Liver
Muscle
Heart
Lymph node
Spleen
Thyroid
Skin
Prostate
Lung
Kidney
Intestine
Breast
Bladder
Adipose
Ileum
UMAP-2
UMAP-1
200
0
-200
-200
0
200

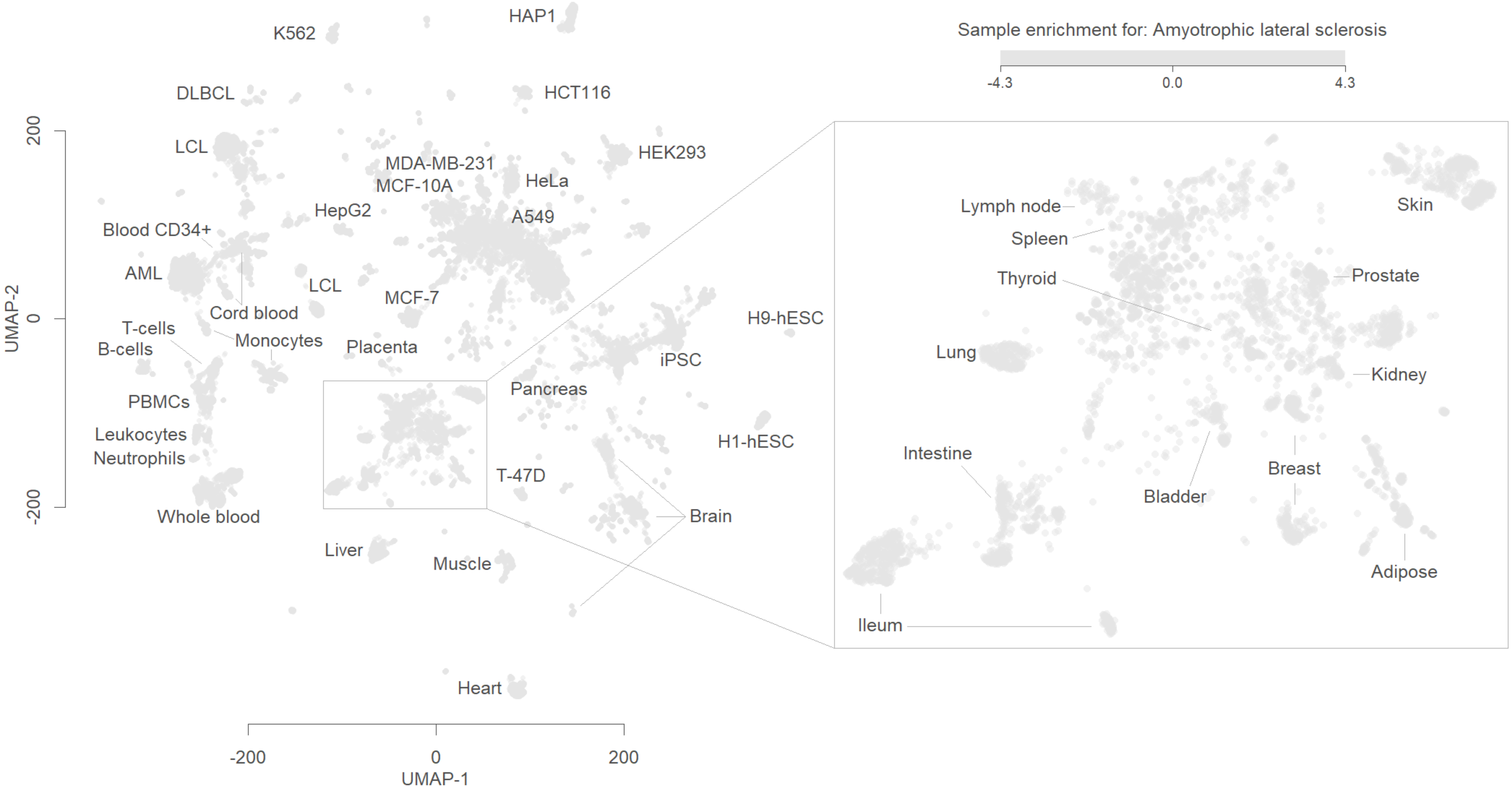
Sample enrichment for: Amyotrophic lateral sclerosis
-4.3
0.0
4.3
HAP1
K562
DLBCL
HCT116
LCL
HEK293
MDA-MB-231
MCF-10A
HeLa
HepG2
A549
Blood CD34+
AML
LCL
Cord blood
MCF-7
T-cells
B-cells
Monocytes
Placenta
H9-hESC
iPSC
Pancreas
PBMCs
Leukocytes
Neutrophils
H1-hESC
T-47D
Brain
Whole blood
Liver
Muscle
Heart
Lymph node
Spleen
Thyroid
Skin
Prostate
Lung
Kidney
Intestine
Bladder
Breast
Adipose
Ileum
UMAP-2
200
0
-200
UMAP-1
-200
0
200

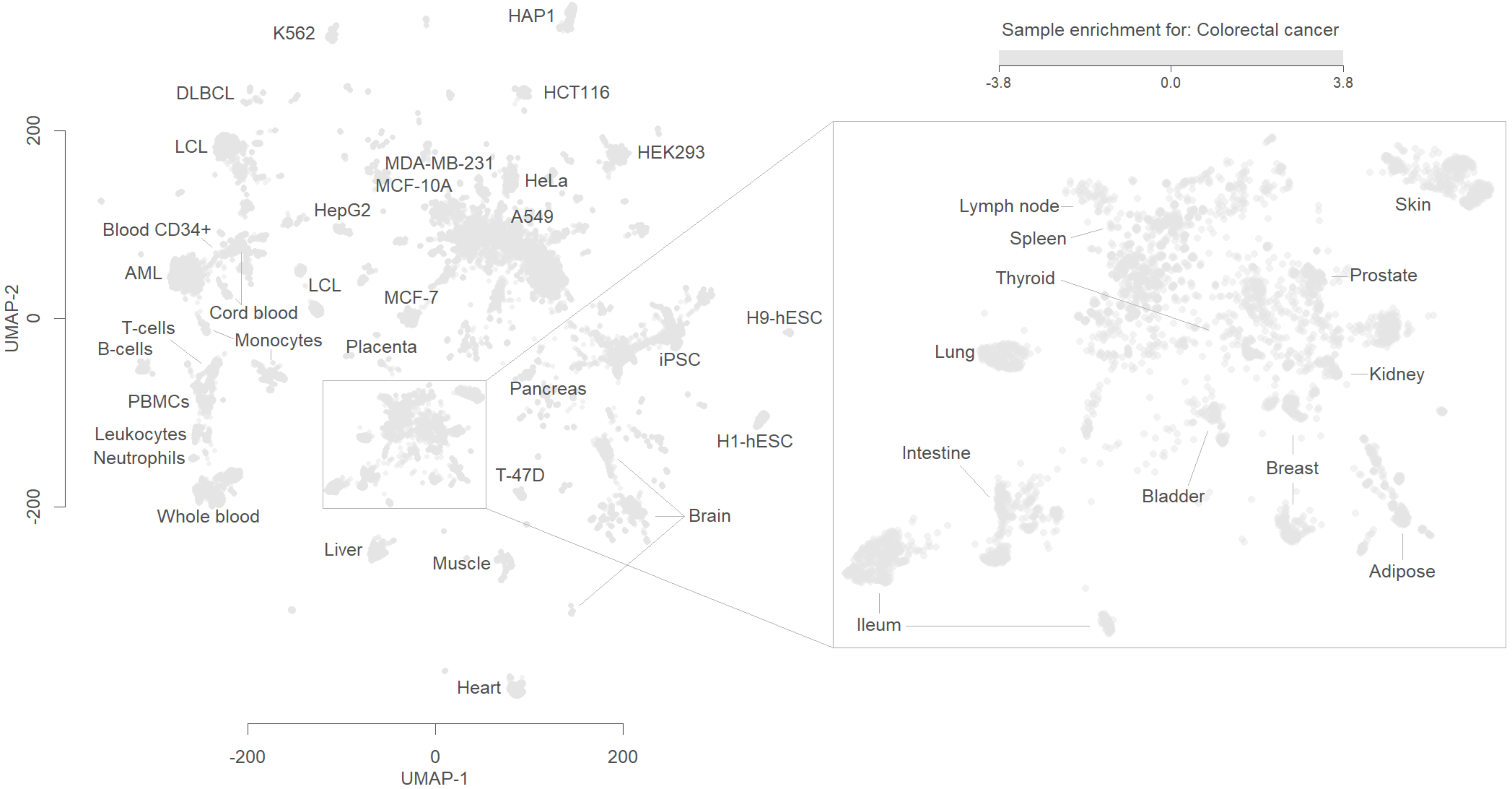

Sample enrichment for: Colorectal cancer
-3.8
0.0
3.8
HAP1
K562
DLBCL
HCT116
LCL
HEK293
MDA-MB-231
MCF-10A
HeLa
HepG2
A549
Blood CD34+
AML
LCL
MCF-7
Cord blood
H9-hESC
T-cells
B-cells
Monocytes
Placenta
iPSC
PBMCs
Pancreas
Leukocytes
Neutrophils
H1-hESC
T-47D
Whole blood
Brain
Liver
Muscle
Heart
UMAP-2
UMAP-1
200
0
-200
-200
0
200
Lymph node
Spleen
Thyroid
Skin
Prostate
Lung
Kidney
Intestine
Breast
Bladder
Adipose
Ileum

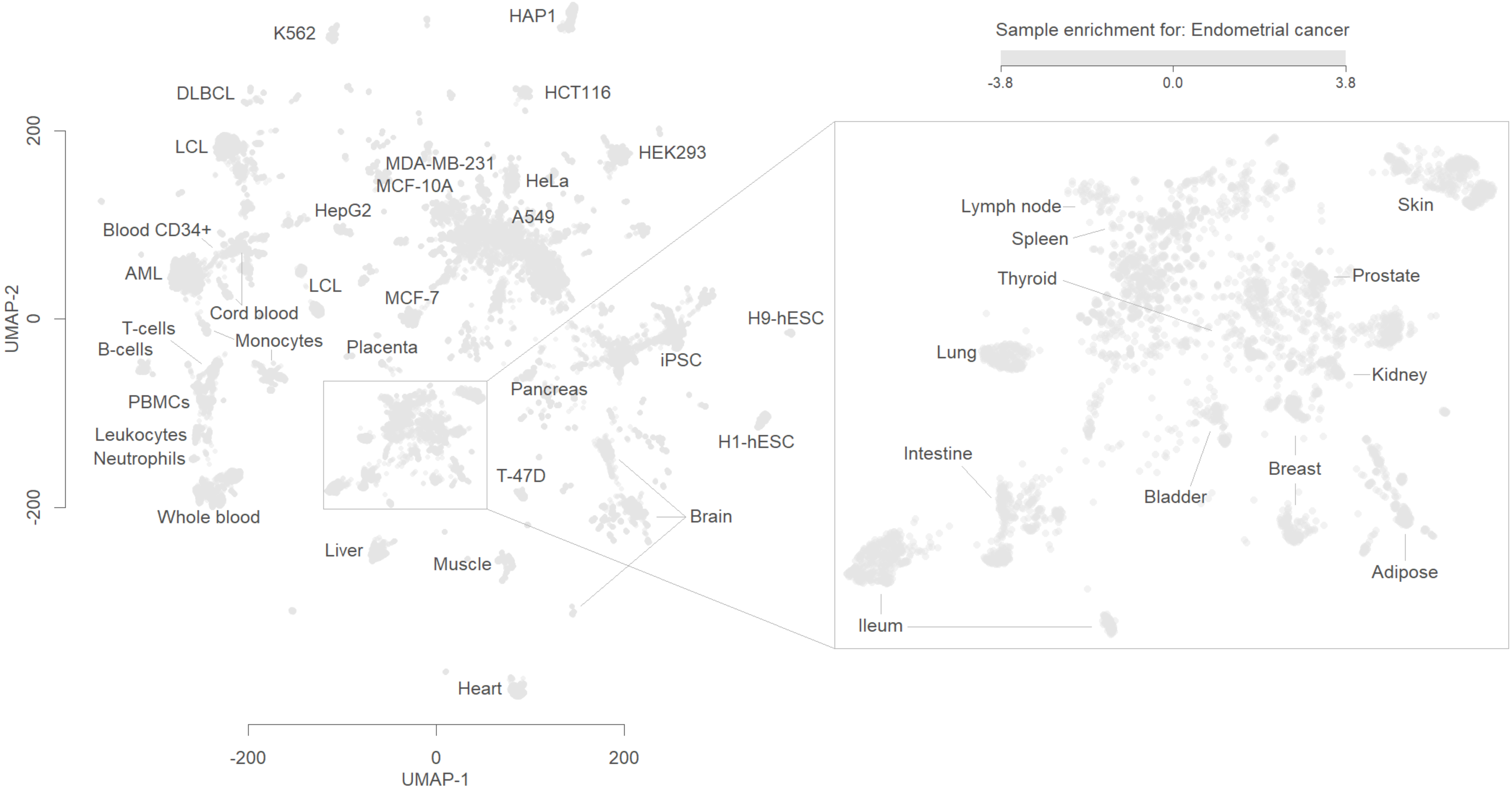
Sample enrichment for: Endometrial cancer
-3.8
0.0
3.8
HAP1
K562
DLBCL
HCT116
LCL
HEK293
MDA-MB-231
MCF-10A
HeLa
HepG2
A549
Blood CD34+
AML
LCL
MCF-7
Cord blood
T-cells
B-cells
Monocytes
Placenta
H9-hESC
iPSC
Pancreas
PBMCs
Leukocytes
Neutrophils
H1-hESC
T-47D
Brain
Whole blood
Liver
Muscle
Heart
Lymph node
Spleen
Thyroid
Skin
Prostate
Lung
Kidney
Intestine
Bladder
Breast
Adipose
Ileum
UMAP-2
200
0
-200
-200
0
200
UMAP-1
